# A unifying functional dichotomy organises breast cancer molecular landscape, resolves PIK3CA ambiguity, and supports tiered tumour classification

**DOI:** 10.64898/2026.02.22.26346715

**Authors:** Amit Gupta, Muthulakshmi Muthuswami

**Affiliations:** Akrivia Biomedics Limited, London, England, UK; Akrivia Biosciences Pvt Ltd, Mumbai, India

## Abstract

Clinical interpretation of breast cancer sequencing is constrained not by a lack of data but by the absence of an organising framework that translates constellations of co-occurring mutations and copy-number alterations into tumour-level biology with prognostic and therapeutic meaning. This challenge is exemplified by PIK3CA, a clinically actionable alteration often treated as a single-label biomarker despite context-dependent associations with outcome. We analysed >5,000 breast tumours across multiple cohorts using integrated multi-omics (somatic mutations, copy-number, transcriptomic, proteomic and phosphoproteomic profiles) and quantified the directionality of downstream molecular consequences of recurrent alterations relative to TP53-associated trends to infer dominant tumour programmes. This revealed a robust functional organisation comprising (i) a canonical proliferative/replicative programme, enriched for cell-cycle, DNA replication and E2F signalling, and encompassing TP53 mutations and most recurrent CNAs, and (ii) a non-canonical signalling/cell-state programme marked by recurrent mutations including PIK3CA, CDH1, GATA3, MAP3K1 and AKT1, with opposing transcriptomic/proteomic directionality, comparatively lower proliferative output and a systematic tendency towards mutual exclusivity with TP53, consistent with alternative evolutionary routes.

To operationalise these findings for clinical use, we developed T-OMICS (<u>T</u>iered <u>OMI</u>CS <u>C</u>lassification <u>S</u>ystem), which layers complementary readouts to deliver a single interpretable tumour profile: Tier 1 provides a continuous genomic-risk backbone via a DNA-anchored prognostic RNA signature capturing canonical proliferative/replicative output; Tier 2 assigns programme identity based on the dominant genomic context; Tier 3 quantifies within-programme activity along a continuum; and Tier 4 overlays non-redundant modifier mutations that refine phenotype, vulnerabilities and resistance liabilities, supported by orthogonal proteomic/phosphoproteomic pathway signals. In ER+/HER2− disease, T-OMICS resolves the prognostic ambiguity of PIK3CA by showing that “PIK3CA-mutant” is not a single biological entity: in a predominant low-genomic-score context, PIK3CA aligns with buffered luminal biology and favourable outcomes, whereas in high-score contexts—conditioned by TP53 background and modifier events—PIK3CA can mark adverse biology with distinct dependencies not captured by proliferation-centric readouts; notably, low-score PIK3CA tumours with CDH1 co-mutation shift to significantly worse outcomes.

Together, these results establish a programme- and state-aware framework that converts sequencing reports into clinically legible tumour biology to support risk calibration, therapeutic prioritisation and evolution-aware sampling decisions from early-stage through metastatic ER+/HER2− breast cancer.

**Lay Summary:** Breast cancer tumours often carry many genetic changes at the same time. While modern sequencing can identify these changes in detail, the results are frequently presented as long lists of mutations and DNA alterations that are difficult to interpret in terms of how a tumour behaves or how it should be treated. A well-known example is the PIK3CA gene: although it can be targeted with specific drugs, studies have reported mixed results on whether PIK3CA mutations are associated with better or worse outcomes, making it challenging to use this information confidently in clinical care.

To address this problem, we analysed genomic (DNA-wide), RNA, and protein data from more than 5,000 breast tumours. We found that many common genomic changes cluster into two main biological “programmes” that reflect distinct ways tumours grow and survive. One programme is driven by rapid cell division and DNA replication and includes TP53 mutations and many common DNA copy-number changes; tumours following this programme tend to be more aggressive. The second programme is less focused on rapid growth and is defined by mutations such as PIK3CA, CDH1, GATA3, MAP3K1, and AKT1, which influence signalling and cell identity rather than directly accelerating proliferation.

These programmes reflect broader tumour behaviours rather than the effects of single genes. Importantly, mutations in the second programme are usually not found alongside TP53 mutations, suggesting that breast cancers can develop through distinct biological routes—with some tumours following an alternative pathway (not overtly proliferation-dependent) that shapes their behaviour and may influence which treatments are most appropriate.

Based on these findings, we developed a practical classification system, T-OMICS, for ER-positive, HER2-negative breast cancer. T-OMICS summarises which biological programme a tumour follows, how active or aggressive it is within that programme, and whether additional mutations are present that may influence treatment response or resistance. Using this framework, we show that PIK3CA mutations most often occur in a biologically buffered context associated with more favourable outcomes, but when they occur in more aggressive tumours—shaped by other key genetic changes—they can signal a higher-risk disease with different treatment needs. These findings indicate that treatment decisions should be based on the tumour’s overall biological pattern, not just the presence of a single mutation. By placing sequencing results in this broader context, T-OMICS supports more accurate risk assessment, better treatment planning, and more informed decisions about when to intensify therapy, from early-stage through advanced breast cancer.

Graphical Summary

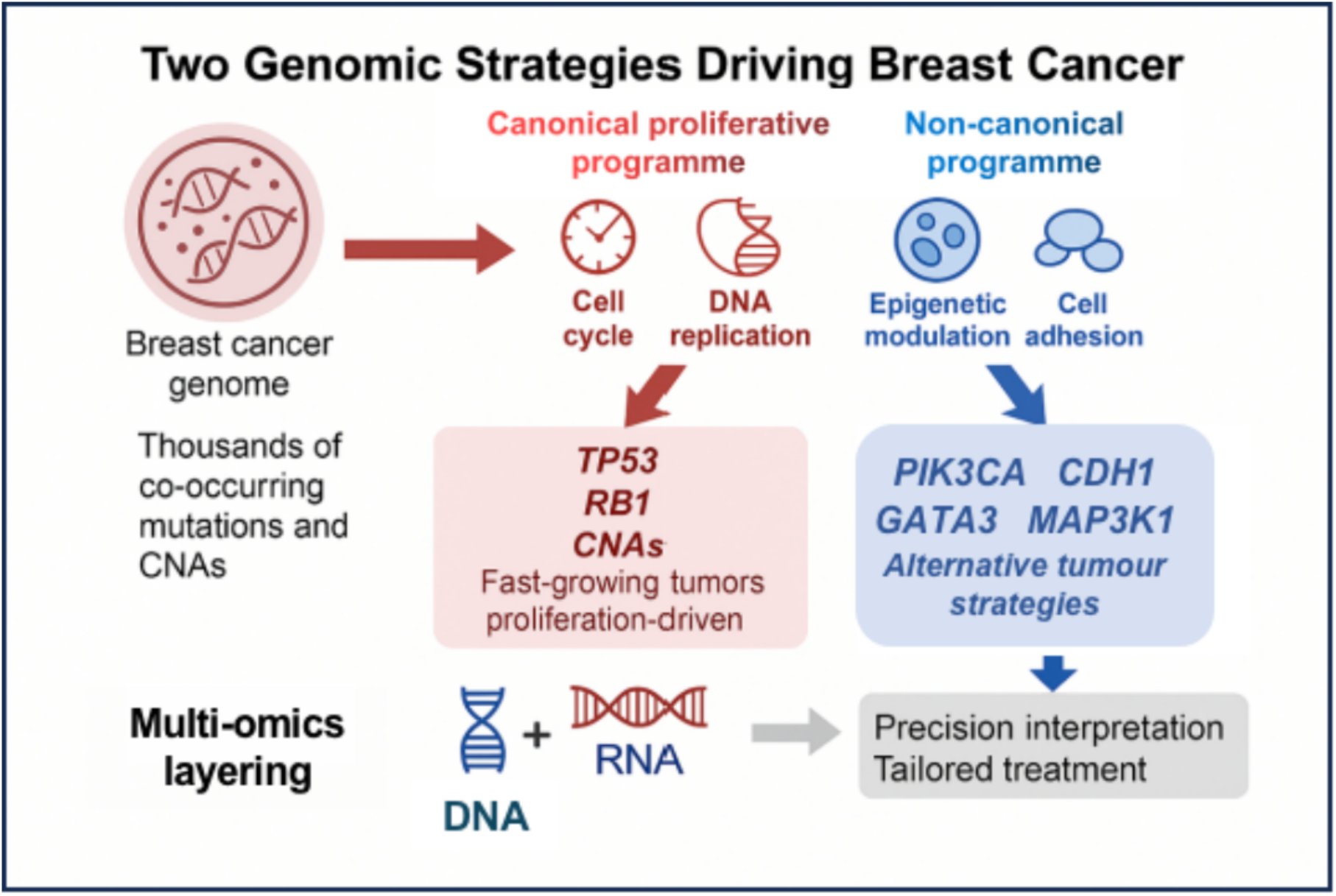

## Introduction

Breast cancer is genomically heterogeneous. Even within clinically defined subtypes, tumours commonly harbour co-occurring somatic mutations and copy number alterations (CNAs) that interact to shape tumour behaviour, therapeutic sensitivity, and clinical outcome [1–3]. Although tumour sequencing is increasingly incorporated into clinical care, translating genomic profiles into clinically usable interpretations remains difficult—particularly when reports enumerate alterations without clarifying the dominant tumour programme they collectively represent or the prognostic context in which mechanistic inferences should be made [4,5]. This challenge is pronounced in oestrogen receptor–positive, HER2-negative (ER+/HER2−) breast cancer, the most prevalent subtype, where recurrent alterations are frequently identified but their prognostic and therapeutic implications are often inconsistent when considered in isolation [6–8]. As a result, clinicians may receive lists of “actionable” or “relevant” genes without a mechanism-first synthesis that explains tumour state, dependencies, resistance liabilities, and rational combination opportunities [4,5,9].

PIK3CA mutations exemplify this interpretability gap. PIK3CA is among the most frequent genomic events in ER+/HER2− disease and is an established predictive biomarker for PI3Kα inhibition in advanced breast cancer [10–12]. Yet, across cohorts and clinical contexts, PIK3CA status has been associated with favourable, neutral, or adverse outcomes, depending on population, intrinsic subtype, exon context, and analytic framing [6,7,13–15]. In practice, PIK3CA is often treated as a single-label biomarker (“PIK3CA-mutant”) without a framework that distinguishes contexts in which the mutation reflects relatively indolent luminal biology from those in which PI3K signalling coexists with adverse biology, resistance liability, or metastatic potential [6,13,16]. Similar context dependence has been suggested for other common ER+/HER2− alterations, including CDH1, GATA3, KMT2C, and MAP3K1, whose contributions may not be reducible to proliferation alone and whose clinical associations can be obscured by population-level averaging [3,17,18]. Collectively, these observations motivate an organising principle that links diverse DNA events and CNA landscapes to tumour-level programmes that are clinically interpretable for risk assessment and treatment planning [4,9].

Existing clinical tools capture important prognostic signals but do not resolve this mechanistic ambiguity. Expression-based assays and intrinsic subtyping frameworks (e.g., multigene recurrence scores and PAM50) quantify proliferative risk and luminal biology, informing adjuvant therapy intensity [19–22], yet they do not explicitly link specific genomic configurations to pathway wiring or resistance routes. Conversely, sequencing-based annotation emphasises canonical oncogenic pathways but often fails to specify how multiple co-occurring events combine to produce the dominant tumour programme, particularly in ER+/HER2− tumours where CNAs are prevalent and genomic instability varies widely [4,23]. Moreover, progression and therapy adaptation in ER+/HER2− disease reflect not only proliferation and genomic instability but also lineage-state regulation, signalling rewiring, microenvironmental interactions, and therapy-selected survival programmes [24–27]. A clinically useful framework should therefore move beyond single-gene labels towards programme-level interpretation while anchoring mechanistic outputs in prognostic context.

To address this need, we developed a discovery-to-translation approach. First, we asked whether apparently heterogeneous genomic alterations can be grouped by shared downstream molecular directionality. Using large breast cancer cohorts, we quantified transcriptomic and proteomic consequences of recurrent alterations relative to TP53 mutation–associated downstream trends [28–30], enabling functional grouping by concordant versus opposing molecular effects. Second, we translated this discovery into tumour-level stratification by deriving complementary RNA- and DNA-based signatures and implementing them in T-OMICS, a tiered tumour classification framework for ER+/HER2− disease. T-OMICS overlays (i) a DNA-anchored RNA signature capturing the cumulative output of a predominantly canonical proliferative/replicative programme with (ii) a complementary DNA mutation signature enriched for mutations mapping to an alternative programme reflecting comparatively non-proliferative mechanisms that are not fully captured by canonical pathway-centric RNA programmes. Proteomic/phosphoproteomic and signalling overlays then connect T-OMICS-defined states to pathway activity and potential therapeutic leverage [31,32]. Conceptually, T-OMICS is hierarchical and modular: it separates core identity from within-subgroup activity/aggressiveness (“state”) and adds modifier events that refine phenotype, vulnerabilities, and resistance liabilities, thereby delivering a mechanistic interpretation explicitly within prognostic strata.

A key clinical premise is that PIK3CA ambiguity is fundamentally context-dependent: the mutation can occur within distinct programmes and states associated with different pathway wiring, evolutionary constraints, and clinical behaviour [6,13,16]. Rather than treating PIK3CA as monolithic, our goal is to define contexts in which it aligns with lower-risk, stress-buffered luminal biology versus those in which it participates in higher-risk biology with distinct dependencies and escape routes—providing a mechanistic basis for previously reported heterogeneous associations [6,7,13–16]. Because clinical utility must extend beyond primary tumours, we also evaluate T-OMICS in paired metastatic samples to distinguish stable truncal identity from state transitions and the acquisition of modifier events, a practical issue in metastatic ER+/HER2− disease where endocrine resistance and lesion-specific evolution can drive heterogeneous vulnerabilities [27,33–35]. Overall, we aim to make sequencing results clinically legible by linking DNA events to tumour programmes and states, interpreted in a prognostic context, to support prognosis, therapeutic targeting, and resistance planning in ER+/HER2− breast cancer [4,5,9].

## Results

### 1. Functional classification reveals two opposing classes of genomic alterations in breast cancer

We analysed more than 3,600 primary breast tumours from TCGA and METABRIC, integrating somatic mutations, CNAs, and transcriptomes (**Figure 1A; Table S1** and **S2**). By comparing the directionality [the fold-change direction] of downstream differential expression across recurrent alterations, we identified a highly robust, previously undescribed dichotomy. TP53 and PIK3CA—the two most recurrent mutations in breast cancer (**Figure S1; Table S3**)—showed diametrically opposing transcriptional consequences, with >90% of shared DEGs showing inverse fold-change directions (genes upregulated in TP53-mutant tumours were downregulated in PIK3CA-mutant tumours and vice versa) (**Figures 1B** and **S2-S4; Table S4**). We found that most recurrent alterations in breast cancer produced downstream molecular effects aligned with either TP53 or PIK3CA mutation states (**Figures 1D** and **S5**). We therefore defined: Group A, alterations with TP53-mutant-like downstream directionality; and Group B, alterations with PIK3CA-mutant-like directionality that exhibit an opposing trend to TP53-mutant tumours (**Figures 1C** and **S5-S7; Table S5**). This functional grouping was consistent across cohorts and robust to subtype stratification and platform-specific batch effects (**Figures S6-S10**; **Table S5**). Notably, the same antagonistic pattern was observed at the proteomic level, reinforcing biological robustness and translational relevance **(Figures 1E** and **S11-S13)**.

**Figure 1.**
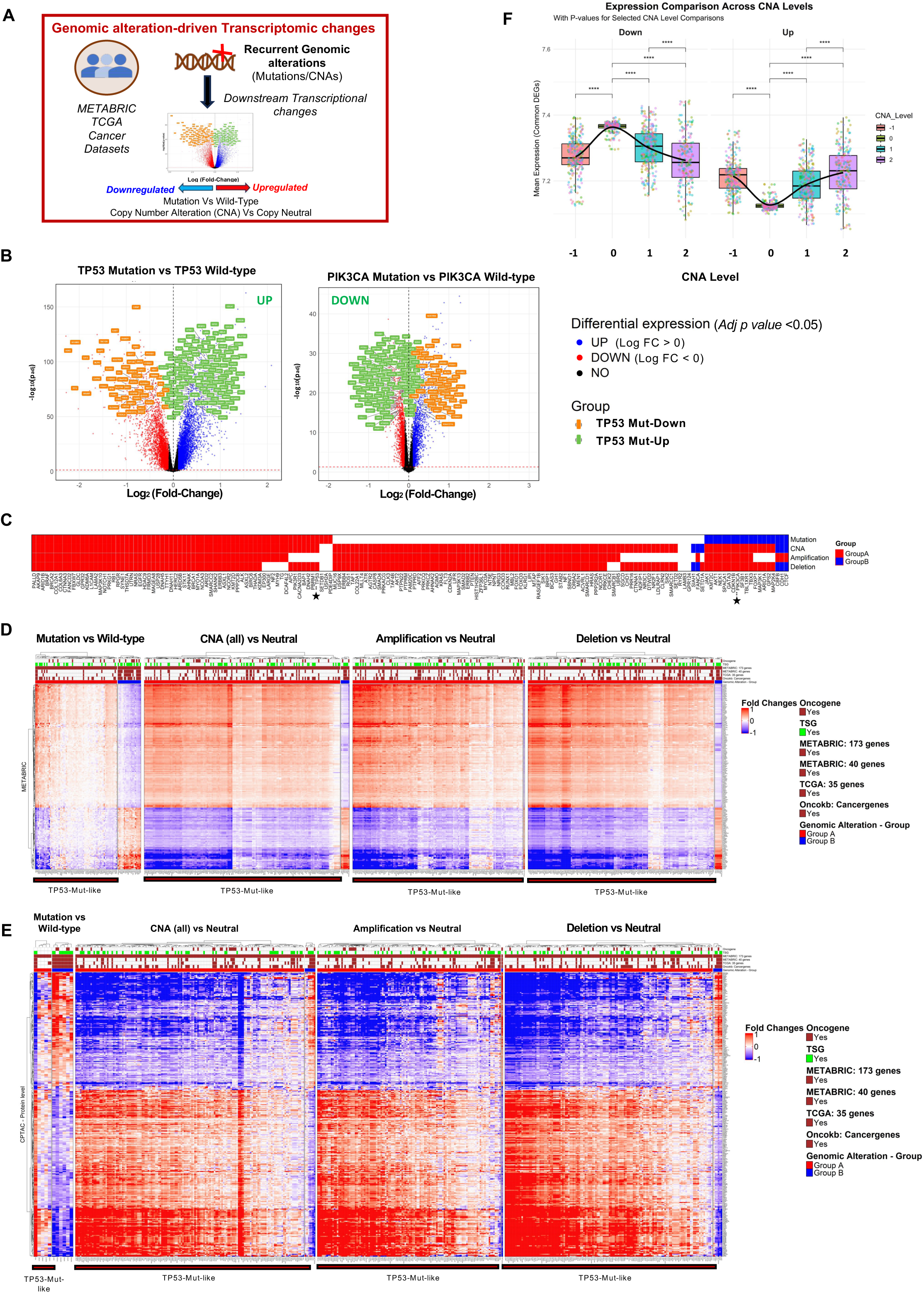
Genomic alteration-driven transcriptional and proteomic programmes reveal two distinct genomic arms in breast cancer. **(A)** Downstream transcriptional effects associated with recurrent genomic alterations (mutations and copy number alterations) were analysed across breast cancer samples from METABRIC, TCGA, and additional cohorts listed in *Supplementary Table 1*. For each recurrent alteration analysed in this study, gene expression differences were quantified by contrasting genomically altered samples (mutant or copy-number altered) with their respective non-altered (wild-type or copy-number neutral) counterparts. **(B)** Volcano plots illustrate significantly altered differentially expressed genes (DEGs) for the two most frequent mutations in breast cancer: TP53-Mutant vs Wild-type (left) and PIK3CA-Mutant vs Wild-type (right). Genes with a significant differential expression (FDR < 0.05) and a log2 fold change greater than 0 are depicted as blue dots, and those with a log2 fold change less than 0 are shown as red dots. Those that fall under the not-significant category are depicted as black dots. Horizontal and vertical dashed lines indicate the thresholds for statistical significance and fold change, respectively. Among the top 500 significantly differentially expressed genes (FDR < 0.05), those overlapping between TP53 and PIK3CA comparisons are labelled, showing reciprocal regulation: genes upregulated in TP53-mutant tumours (gene names highlighted in green) are downregulated in PIK3CA-mutant tumours, while genes downregulated in TP53-mutant tumours (highlighted in orange) are upregulated in PIK3CA-mutant tumours (see *Supplementary Table 4*). **(C)** Functional class assignment of recurrent genomic alterations in breast cancer based on cluster consistency and stability analysis (see *Methods*). Fold-change profiles from Mutation vs Wild-type, CNA vs copy-number Neutral, Amplification vs copy-number Neutral, and Deletion vs copy-number Neutral comparisons were subjected to unsupervised hierarchical clustering, revealing two major transcriptional arms: (1) a TP53-like downstream directionality cluster (*Group A*), and (2) a PIK3CA-like directionality cluster that exhibits opposing signatures to TP53-mutant tumours (*Group B*). Cluster consistency and stability were assessed using both predefined and randomised gene sets across three cohorts (comprising ∼3600 patients’ data; see *Methods*), utilising different gene expression profiling techniques (microarray/RNA-seq), and classifying recurrent mutations into either Group A or Group B. The resulting group assignments are summarised as a heatmap in Figure 1C, where red denotes Group A, blue denotes Group B, and white/blank corresponds to not determined (ND; insufficient evidence for classification) or missing data (NA) (see *Supplementary Table 5*). **(D, E)** Heatmaps show the fold-change patterns of TP53-mutant vs wild-type DEGs/significantly altered proteins across mutation, CNA (amplifications and deletions combined), amplification, and deletion comparisons, using transcriptomic **(D)** and proteomic **(E)** data from the METABRIC and CPTAC datasets, respectively. Each alteration’s downstream impact was assessed via fold-change patterns in gene or protein expression relative to unaltered samples. The heatmap shows fold-change correlations across mutations/CNAs. In all cases, DEGs/altered proteins exhibit a consistent yet opposite fold-change trend between the Group A (TP53-like) and Group B (PIK3CA-like) sets of recurrent alterations. Columns METABRIC_40, Genes_35, METABRIC_173, and Oncokb_Cancergenes denote membership of each gene in previously published or curated gene signatures (see *Supplementary Table 2***). (F)**The boxplot shows the relationship between cancer-associated CNA-driven and TP53 mutation-associated transcriptomic programmes, with CNA dosage showing a monotonic strengthening of the TP53-like transcriptional programme. It depicts the distribution of mean DEG expression (derived from TP53 Mutant vs Wildtype analysis; shown on the y-axis) across the CNA levels (Copy −1, 0, 1, 2; shown on the x-axis) of cancer-associated genes (derived from OncoKb; each dot in the boxplot represents a cancer-associated gene across the various CNA levels), with a loess-smoothed curve overlaid to illustrate overall global trends. Expression of all DEGs identified in the TP53 Mutant vs Wild-type analysis is shown for the METABRIC cohort (n = 12,021 DEGs, based on FDR<0.05 cut-off). UP and DOWN DEGs are shown in two separate panels (left: DEGs downregulated in TP53 mutant tumours, right: DEGs upregulated in TP53 mutant tumours). CNA levels −2 and −1 were merged due to limited −2 samples, and +2 alterations were included only if supported by ≥10 samples (≥0.5% of cohort). Both copy losses and gains of cancer-associated genes produce expression profiles that parallel TP53 mutant–like transcriptional programmes (see also related *Supplementary Figure 4*). TP53-WT: TP53 wild-type; PIK3CA-WT: PIK3CA wild-type.

### 2. Group A is dominated by CNAs and enriched for canonical proliferation and DNA replication programmes

CNAs are prevalent in breast cancer and contribute to progression through genomic instability and dosage effects [36]. Strikingly, most cancer-associated CNAs mapped to Group A, producing TP53-like downstream transcriptional directionality irrespective of CNA polarity (loss versus gain; even when copy loss or gain affects the same gene) and irrespective of whether the affected gene is annotated as an oncogene or tumour suppressor (**Figures 1C, 1D**, **S7-S10, S13,** and **S14**). Across CNA dosage states, a clear monotonic gradient emerged– the TP53-like programme strengthened with increasing CNA magnitude, peaking in deep deletions and high-level amplifications (**Figures 1F, S15,** and **S16**), emphasising the robustness of this association and underscoring its translational relevance. This pattern was reproducible across adjacent CNA states and supported by stepwise regulation of TP53-aligned DEGs. Beyond CNAs, many recurrent mutations (e.g., RB1, SYNE1, DNAH11, AHNAK, BRCA1/2) also mapped to Group A (**Figures 1C, 1D, S5,** and **S6; Table S5**). Gene set enrichment of Group A downstream effects consistently highlighted canonical tumour-promoting programmes, including Cell Cycle, DNA Replication, E2F Targets, and G2/M Checkpoint (FDR <0.05), alongside pathways linked to genomic instability and DNA repair **(Figures 2A-2C**, and **S17-S19)**. Similar CNA-associated TP53-like downstream patterns were also observed across additional cancer types, indicating broad relevance beyond breast cancer (**Figure S20)**.

**Figure 2.**
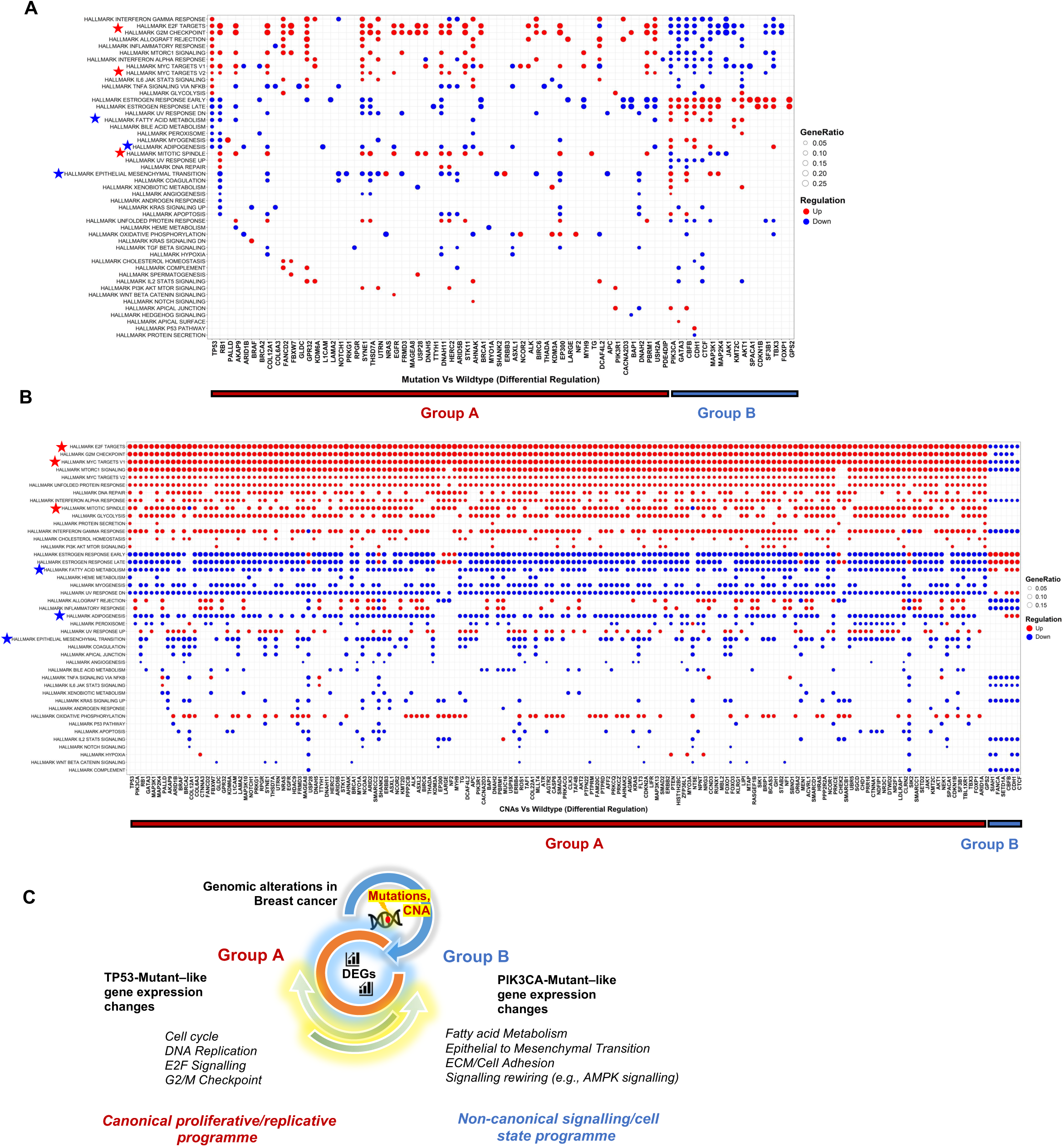
Two dominant functional programmes driven by genomic alterations in breast cancer. **(A)** Hallmark pathway gene set enrichment from differential expression analyses comparing mutant versus wild-type samples for Group A and Group B genes from Figure 1C in the METABRIC cohort, visualised as a dot plot. Each column corresponds to a Mutation vs Wildtype comparison, and each row represents a significantly enriched Hallmark gene set. Red and blue indicate enrichment of upregulated and downregulated Hallmark gene sets, respectively. Two major transcriptional patterns are annotated along the x-axis: Group A (gene alterations showing TP53-like transcriptional directionality and enriched for the canonical proliferative/replicative programme) and Group B (showing PIK3CA-like directionality and an opposing molecular trend to TP53 mutant tumours, influencing non-canonical signalling/cell-state programmes rather than directly accelerating proliferation/DNA replication). **(B)** Hallmark pathway enrichment for copy-number altered (CNA) versus copy-number neutral samples is shown using a similar dot-plot layout, where each column represents a CNA comparison, and each row represents enriched Hallmark gene sets. **(C)**Overall, mutation-associated expression signatures segregate into two distinct transcriptional clusters, whereas CNA-associated signatures predominantly converge on proliferation-linked pathways, aligning primarily with Group A.

### 3. Group B mutations define distinct, non-canonical, comparatively non-proliferative tumour programmes

In contrast, a smaller but recurrent set of mutations—including PIK3CA, GATA3, CDH1, MAP3K1, KMT2C, and AKT1—mapped to Group B and exhibited downstream directionality **opposing** Group A/TP53 **(Figures 1C, 1D, S5,** and **S6; Table S5)**. Group B alterations were not enriched for canonical proliferation/replication pathways and instead implicated alternative biological processes, including metabolic programmes, epigenetic regulation, cell adhesion, and signalling rewiring. Consistent with this, Group B tumours exhibited fewer features of genomic instability than Group A and scored low on standard proliferation-based gene sets (**Figures 2A-2C** and **S17-S19**). Proteomic analyses recapitulated the Group A–Group B antagonism, with Group A alterations enriched for canonical proliferation programmes among the differentially expressed proteins (data not shown), further supporting the mechanistic distinctiveness and translational value of this classification.

### 4. Functional grouping is preserved in metastatic breast cancer and highlights clinically relevant enriched mutations

To assess whether the functional grouping identified in primary tumours remains relevant in advanced disease, we extended the analysis to metastatic breast cancer, which is known to acquire additional or enriched mutations under therapy-induced selection pressures, clonal evolution, and disease progression. TP53 and PIK3CA remained the two most recurrent mutations in metastatic samples **(Figure S21A)** and continued to exhibit strongly opposing downstream transcriptional programmes, consistent with the inverse relationship observed in primary tumours **(Figure 3A)**. Importantly, the overall assignment of recurrent alterations to Group A versus Group B was largely preserved in metastases, supporting the robustness of this functional classification under therapy-driven selection and evolutionary pressure (**Figures 3A, 3B, S21B,** and **S22**).

**Figure 3.**
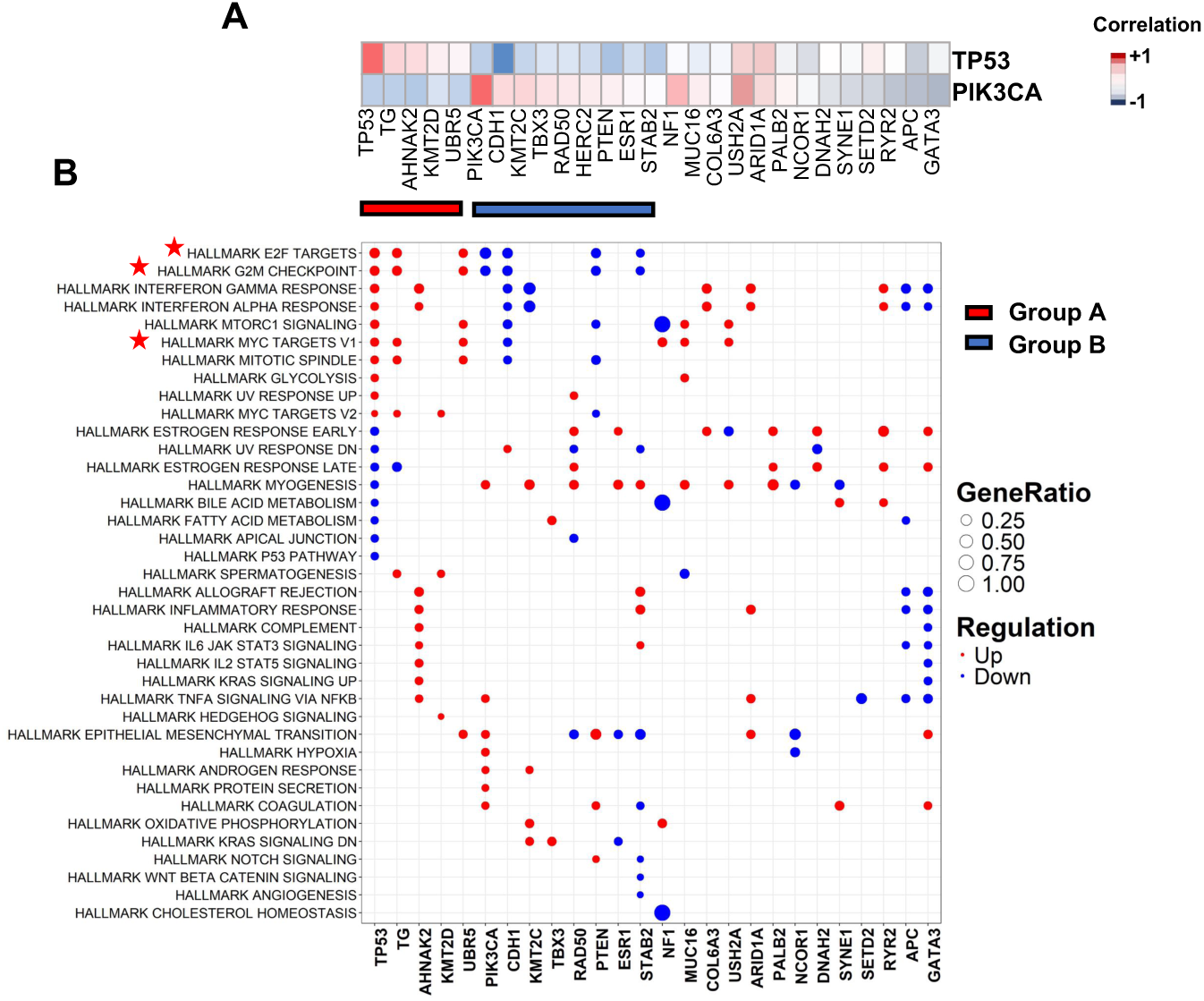
Transcriptional consequences of genomic alterations in metastatic breast cancer. **(A)** Fold-change-based correlation heatmap of recurrent mutations in metastatic breast cancer relative to TP53 and PIK3CA reference profiles. For each recurrent mutation, a vector of log2 fold-changes from mutant versus wild-type comparisons was computed and correlated with TP53-mutant and PIK3CA-mutant reference fold-change vectors. Mutations positively correlated with TP53-mutant tumours and negatively correlated with PIK3CA-associated changes were classified as Group A (TP53-like), whereas those positively correlated with PIK3CA-mutant tumours and negatively correlated with TP53-associated changes were classified as Group B (PIK3CA-like). Two distinct transcriptional patterns observed are annotated below the plot: Group A (TP53-like transcriptional directionality) and Group B (PIK3CA-like directionality and an opposing molecular trend to TP53 mutant tumours). Only genes with ≥3% mutation frequency and those listed as cancer-associated genes (full list in *Supplementary Table 2*) were included. **(B)** Hallmark pathway gene sets enriched in differential gene expression analyses comparing mutant versus wild-type samples in metastatic breast cancer are visualised as a dot plot. Each column represents a mutation-specific comparison that passed the significance threshold (*p < 0.05*), and each row corresponds to a significantly enriched hallmark gene set. The HERC2 mutation-specific comparison did not show enrichment for any hallmark pathways and is therefore not displayed in the dot plot. Red and blue indicate enrichment among upregulated and downregulated gene sets, respectively.

Notably, metastasis-enriched mutations, including ESR1 (a hallmark of endocrine therapy resistance) and PTEN (linked to PI3K pathway deregulation and immune-relevant phenotypes), were predominantly assigned to Group B. This suggests that Group B alterations—representing non-canonical, comparatively non-proliferative tumour-promoting mechanisms—are not only retained during progression but may also be preferentially selected in the metastatic setting, where survival, adaptation, and therapeutic escape can outweigh pure proliferative advantage. These findings support the translational utility of the framework for stratifying vulnerabilities across the disease continuum, from early-stage tumours to advanced/metastatic disease.

### 5. Group A and Group B exhibit structured exclusivity and co-occurrence consistent with alternative evolutionary routes

We observed strong mutual exclusivity between TP53 and most Group B driver mutations, including PIK3CA, GATA3, CDH1, CBFB, CTCF, MAP3K1, TBX3, and SF3B**1** (**Figure 4A; Table S6**), emphasising their crucial role in cancer development. Notably, in metastatic breast cancer, this exclusivity is particularly evident for TP53 with CDH1 and ESR1 **(Figure 4B; Table S7)**. Conversely, Group B alterations frequently co-occurred, particularly with PIK3CA (e.g., co-mutation with MAP3K1, CDH1, CBFB, KMT2C, or SF3B1), and in metastatic disease, PIK3CA and ESR1 frequently co-occur, consistent with cooperative, complementary non-proliferative mechanisms. Together, these patterns support distinct evolutionary strategies underpinning breast cancer progression (e.g., heavily proliferation-dependent vs. alternative mechanisms).

**Figure 4.**
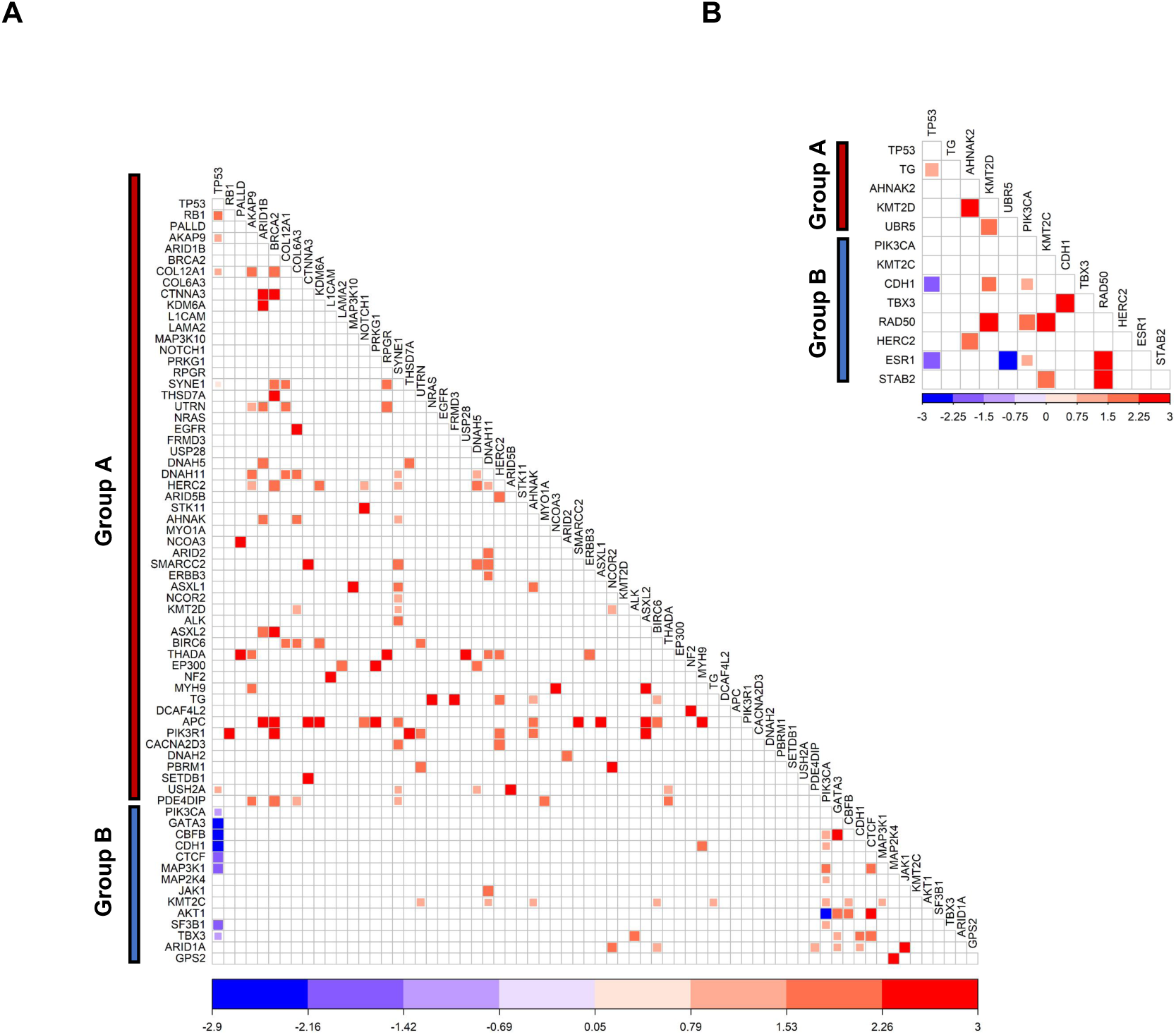
Mutual exclusivity between TP53 and Group B mutations defines distinct oncogenic trajectories. **(A)** Association plots show the log-odds ratios for co-occurrence of genomic alterations across mutation data from 3610 tumour samples (combined METABRIC and TCGA data). Positive log-odds ratios (red) indicate co-occurring alterations, whereas negative log-odds ratios (blue) indicate mutual exclusivity. A pronounced mutual exclusivity is observed between TP53 mutations and Group B alterations. **(B)** Association analysis of metastatic breast cancer data (n = 1,046 samples across five cohorts; *Supplementary Table 1*) demonstrates strong mutual exclusivity between TP53 and Group B mutations–CDH1 and ESR1. Only statistically significant associations (p < 0.05) are shown. Group A and Group B alterations are annotated.

### 6. Development of a DNA-Anchored RNA Signature Capturing Canonical Proliferative/Replicative Output in the Context of a Complementary Non-Canonical Landscape

Building on the Group A/Group B functional framework, we translated alteration-level programmes into tumour-level stratification by deriving a DNA-anchored RNA signature that captures the shared downstream transcriptional state induced across the predominantly canonical proliferative/replicative landscape, while retaining a complementary layer to represent non-canonical mechanisms. We used the two most informative programme anchors—TP53 (Group A; canonical) and PIK3CA (Group B; non-canonical)—to define a transcriptional axis reflecting the cumulative output of the broader genomic landscape rather than any single alteration.

Shared DEGs between TP53 (TP53 mutations linked to poor survival; **Figure S23**) and PIK3CA-mutant tumours showed strongly opposing expression directions (**Figures 1B** and **S2-S4; Table S4**) and were enriched for classical oncogenic programmes, including cell cycle progression and DNA replication (**Figure S24**). This shared DEG set was broadly perturbed across Group A events, including recurrent CNAs and mutations (**Figures 1D, 1F,** and **S6-S10**), supporting the interpretation that it represents a core transcriptional output of a canonical proliferation/genomic instability axis rather than a TP53-specific footprint. Because a subset of tumours is wild-type for both TP53 and PIK3CA, incorporating both wild-type comparators enriched for genes most consistently aligned with the canonical proliferative/replicative axis while filtering context-specific effects. Consistent with this, the shared DEG set showed the largest fold-changes in response to cancer-relevant CNA gains and losses, compared with (i) TP53-exclusive DEGs and (ii) the complete TP53-mutant DEG set (**Figure S15D**), indicating sensitivity to the copy-number–rich, genomically unstable canonical landscape that dominates breast tumour genomes and strengthening its translational utility as a tumour-level readout of cumulative genomic output.

From the shared DEG set, Cox proportional hazards modelling yielded an 11-gene DNA-anchored prognostic RNA signature that generates a continuous composite genomic risk score stratifying early-stage ER+/HER2− tumours into Tier 1 low- and high-risk groups (**Figure S25; Table S8**). Anchored to Group A directionality, the score summarises canonical proliferative/replicative output as a continuous tumour-level measure. Given the clinical use of expression-based risk stratification, we next assessed concordance with the PAM50/Prosigna Risk-of-Recurrence (ROR) framework, in which Luminal A and Normal-like intrinsic subtypes commonly map to low-ROR risk groups [22,37]. This revealed strong agreement between PAM50-based ROR risk groups and our composite-score-defined risk groups, with 84% overall concordance (including 92% within the low-genomic-score group and 76% within the high-score group; the low vs high-score group ratio was 54:46 using our approach and 44:56 using the PAM50-based ROR approach) (**Table S9**).

Notably, this DNA-anchored RNA signature was designed to quantify dominant canonical proliferative/replicative output and is not intended to fully represent the comparatively non-proliferative mechanisms mapped to Group B alterations (e.g., PIK3CA, CDH1, MAP3K1). Accordingly, we interpret the RNA risk score alongside a complementary Group B mutation layer, enabling a programme-aware stratification framework, T-OMICS (<u>T</u>iered <u>OMI</u>CS <u>C</u>lassification <u>S</u>ystem), that captures tumour behaviours, dependencies, and resistance liabilities beyond what proliferation output alone or single-omics would predict (discussed below).

### 7. DNA–RNA integration yields context-aware stratification in ER+/HER2− breast cancer and resolves the prognostic ambiguity of PIK3CA

Integrating the DNA-anchored RNA genomic score with PIK3CA status resolved the long-standing ambiguity in PIK3CA prognostic associations by explicitly linking effects to tumour context. In low-score tumours, PIK3CA mutations aligned with epithelial differentiation and Hippo-pathway activation and were associated with favourable biology (**Figures 5B, S26A,** and **S26B**). Notably, lymph-node involvement—although prognostically adverse in PIK3CA–wild-type disease—had a markedly attenuated impact in PIK3CA-mutant cases and was instead associated with favourable outcomes (p<0.05; **Figure 5C, *left panel***), consistent with relative resistance to progression in this context. In contrast, within high-score tumours, PIK3CA mutations were paradoxically associated with poorer survival despite significantly lower composite genomic scores than PIK3CA–wild-type cases (**Figures 5D, 5E,** and **S26C**), and lymph-node involvement disproportionately worsened outcomes in PIK3CA-mutant disease (**Figure 5C, *right panel***). Together, these results show that PIK3CA effects are determined by tumour programme and activity state, and are not captured by mutation status (**Figure 5A**) or the RNA score alone.

**Figure 5.**
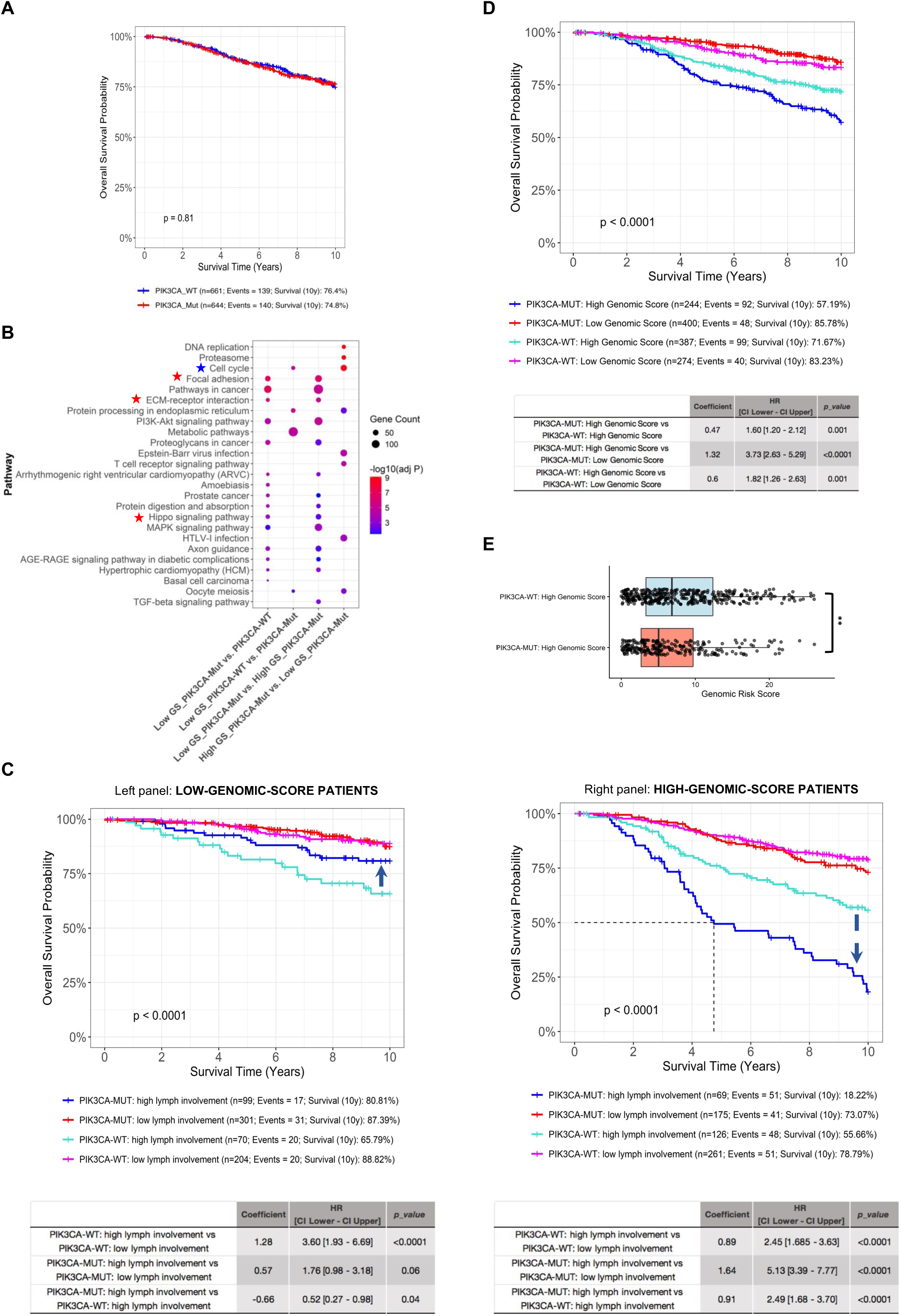
Layered DNA–RNA-based stratification resolves the prognostic ambiguity of PIK3CA in ER+/HER2– breast cancer. **(A)** Kaplan-Meier (KM) survival curve showing the prognostic impact of PIK3CA mutations in ER+/HER2– patients from the METABRIC cohort without molecular subtyping (based on population-level averaging of PIK3CA-mutant vs wild-type patients). Breast cancer-specific overall survival probability is shown on the y-axis. **(B)** KEGG pathway gene set enrichment from differential expression analyses comparing PIK3CA-mutant and wild-type samples across low- and high-genomic-score tumours, visualised as a dotplot. Each column corresponds to a subgroup-wise comparison, and each row represents a significantly enriched KEGG gene set. The analysis uses upregulated DEGs from each subgroup-wise comparison, as indicated on the x-axis. Red and blue asterisks denote key upregulated and downregulated KEGG gene sets in PIK3CA-mutant low-genomic-score tumours, respectively. **(C)**KM survival curves showing the prognostic impact of PIK3CA mutations with lymph-node involvement in low-genomic-score (left panel) and high-genomic-score (right panel) patients. Low lymph-node involvement is defined as up to 1 lymph node involved, whereas high lymph-node involvement is defined as 2 or more lymph nodes involved. A blue dotted arrow denotes the opposite hazard ratio (HR) direction relative to the high-genomic-score PIK3CA wild-type reference group in the two panels. **(D)** KM survival plot showing the overall prognostic impact of PIK3CA mutations in the low- and high-genomic-score groups. Figures C and D show data from the ER+/HER2– METABRIC cohort (n=1305), with 10-year breast cancer-specific overall survival probability shown on the y-axis. Patients with follow-up beyond 10 years were censored. The tables below each survival plot report pairwise comparisons relative to the reference group as indicated, including regression coefficients, HRs with 95% confidence intervals (CIs), and corresponding *p-values*. An HR > 1 indicates increased risk, whereas an HR < 1 indicates reduced risk relative to the reference group. Annotations on the survival plots indicate the total number of patients per group, the total number of observed events, and the survival probability at the specified time. **(E)** Boxplot showing the composite genomic scores across PIK3CA-mutant and wild-type high-genomic-score patients. PIK3CA-mutant tumours have significantly lower genomic scores than PIK3CA wild-type tumours, yet these patients paradoxically have poorer survival. Pairwise comparisons between groups were performed using the Wilcoxon rank-sum test (*p < 0.01*).

To capture TP53–PIK3CA interactions, we co-interpreted RNA score (low vs high) with TP53 and PIK3CA mutation status (TP53 mutations were rare in low-score tumours), defining six Tier 2 prognostic subgroups (**Figures 6A** and **S47**): 1PM (low-score/PIK3CA-mutant), 1PW (low-score/PIK3CA-wild-type), GP2 (high-score/TP53-wild-type and PIK3CA-wild-type), GP3 (high-score/TP53-wild-type and PIK3CA-mutant), GP4 (high-score/TP53-mutant and PIK3CA-wild-type), and GP5 (high-score/TP53-mutant and PIK3CA-mutant). Within high-score tumours, outcomes differed by TP53 status in both PIK3CA-mutant and PIK3CA–wild-type cases (GP3 vs GP5 and GP2 vs GP4, respectively), consistent with distinct co-mutation–defined mechanisms. Despite lower composite genomic scores, GP3 had worse survival than GP2, further underscoring the context-dependent role of PIK3CA in ER+/HER2− disease (**Figures 6B, 6C, S27A** and **S27B**).

**Figure 6.**
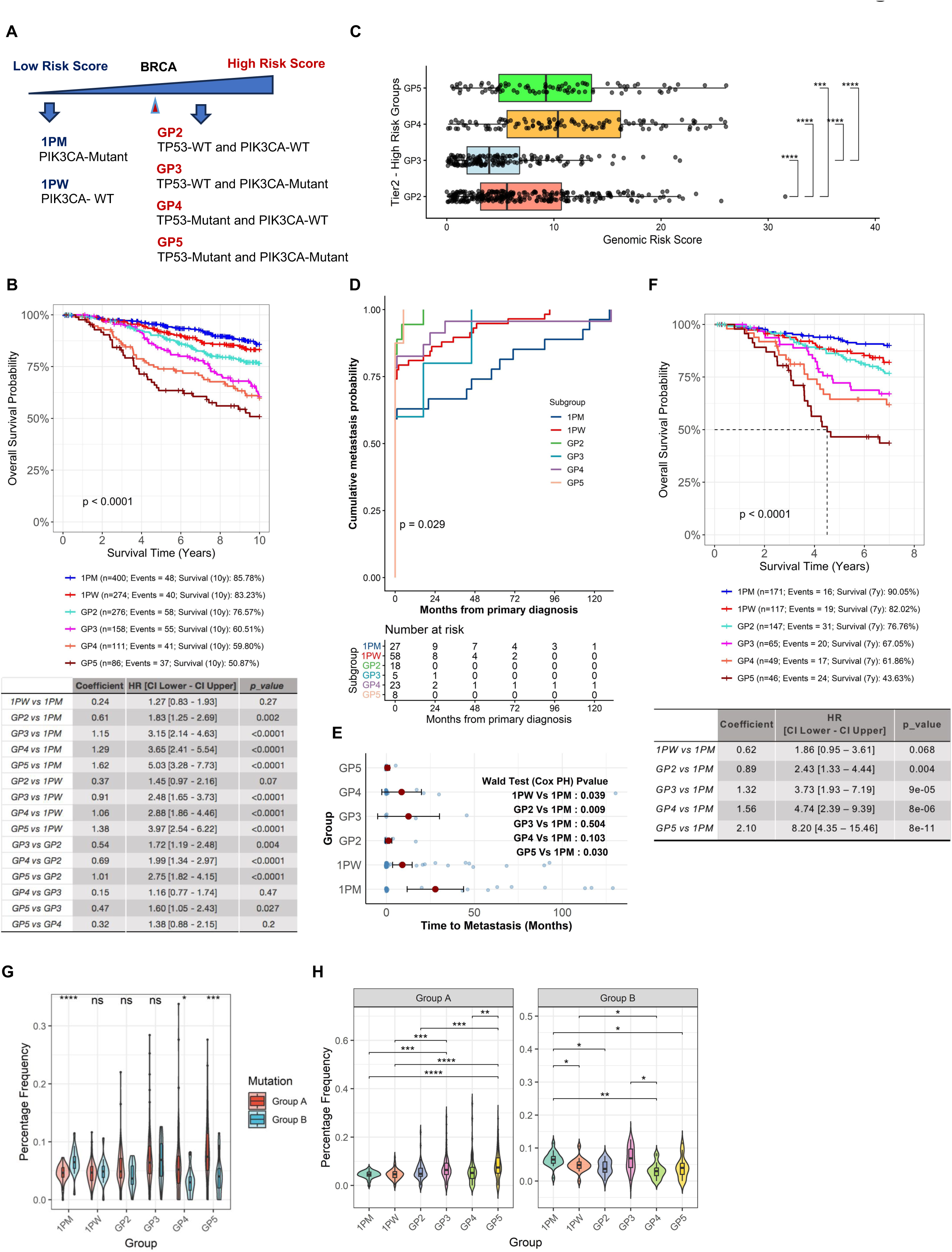
A layered DNA–RNA-based framework yields clinically relevant stratification in ER+/HER2– breast cancer. **(A)** A schematic overview of the Tier 2 stratification framework. Breast cancer samples were stratified into six molecular subgroups by combining PIK3CA and TP53 somatic mutation status (DNA) with Tier 1 RNA signature-based risk groups. **(B)** KM survival plot showing distinct prognostic outcomes associated with the identified Tier 2 subgroups in ER+/HER2– patients from the METABRIC cohort (n=1305). The 10-year breast cancer-specific overall survival probability is shown on the y-axis. Patients with follow-up beyond 10 years were censored. **(C)** Boxplot showing composite genomic scores across the high-genomic-score subgroups. Despite lower genomic scores, GP3 shows worse survival outcomes than GP2, underscoring the context-dependent role of PIK3CA in ER+/HER2– disease. Pairwise comparisons between groups were performed using the Wilcoxon rank-sum test, with statistical significance indicated as follows: *p < 0.05 (*), p < 0.01 (**), p < 0.001 (***), and p < 0.0001 (****)*. **(D)** KM analysis of cumulative metastasis-free survival across Tier 2 subgroups in the Metastatic breast cancer cohort (*Supplementary Table 1)*, demonstrating significant differences in the occurrence of metastatic diagnosis among subgroups (log-rank *p < 0.05*). **(E)** A jittered point-and-interval plot displaying individual metastatic event observations alongside group-level mean estimates and confidence intervals. Cox proportional hazards analysis of time to metastasis across Tier 2 subgroups reveals significant differences between 1PM and other groups, with 1PM showing a longer time to metastasis. **(F)** KM survival plot showing distinct prognostic outcomes associated with the identified Tier 2 subgroups in lymph-node-positive ER+/HER2– patients (with 1 or more lymph-node involvement) from the METABRIC cohort (n=595). The 7-year breast cancer-specific overall survival probability is shown on the y-axis. Patients with follow-up beyond 7 years were censored. In figures B and F, the table below the survival plots report pairwise comparisons relative to the reference group as indicated, including regression coefficients, HRs with 95% confidence intervals (CIs), and corresponding *p-values*. Annotations on the survival plots indicate the total number of patients per group, the total number of observed events, and the survival probability at the specified time. **(G)** Violin plot showing the distribution of average mutation frequencies of Group A and Group B genes across Tier 2 subgroups. Group A mutations are markedly enriched in the TP53-mutant subgroups (GP4 and GP5), whereas Group B mutations are more prevalent in PIK3CA-mutant subgroups, 1PM and GP3. **(H)** Corresponding violin plots with embedded boxplots are shown as separate panels for Group A and Group B. Violin width reflects data density, and boxplots indicate the interquartile range (IQR) with the median shown as the central line. Statistical comparisons were performed within each mutation category, and only statistically significant results are shown in the figure: *p < 0.05 (*), p < 0.01 (**), p < 0.001 (***), or p < 0.0001 (****)*. The y-axes are scaled independently for each panel.

Notably, among patients who ultimately developed metastasis, the time from primary diagnosis to metastatic presentation was significantly longer in 1PM than in 1PW (and in the high-score subgroups) (log-rank *p<0.05*; **Figures 6D** and **6E**), consistent with a favourable biology in low-score PIK3CA-mutant tumours. The six subgroups also remained prognostic in lymph-node–positive disease, with 1PM showing consistently favourable outcomes (**Figures 6F, S27C,** and **S27D**). Genomically, CNA burden and Group A mutation frequencies were substantially lower in low-score tumours (**Figures 6G, 6H,** and **S28-S30**), supporting the view that the RNA score tracks a proliferation/genomic-instability axis. Notably, GP3 showed aggressive outcomes despite a significantly lower CNA burden, indicating adverse biology that is not simply a function of genomic instability. In GP3, Group A and Group B mutations were similarly enriched, whereas Group B mutations were generally enriched in PIK3CA-mutant subgroups (1PM and GP3) and largely excluded from TP53-mutant subgroups (GP4 and GP5), consistent with the alternative evolutionary routes described above.

### 8. Subgroup-Specific Tumor Biology and Targetable Vulnerabilities Revealed by Tiered Multi-Omics Profiling

To define mechanisms underlying each Tier 2 molecular subgroup, we performed Gene Set Variation Analysis (GSVA) across ER+/HER2– tumours using >10,000 MSigDB gene sets, providing a global map of pathway themes across subgroups. Analyses were conducted independently in METABRIC and TCGA (tumour and normal tissues) and validated in an external multi-omics cohort (**Table S1**), showing strong cross-cohort concordance of subgroup pathway signatures (**Figure 7C**). Given this reproducibility, datasets were pooled for pathway meta-analysis (∼2,500 samples) to improve effect-size precision and strengthen subgroup-specific target nominations (**Figures 7A, 7B,** and **S31A-C**).

**Figure 7.**
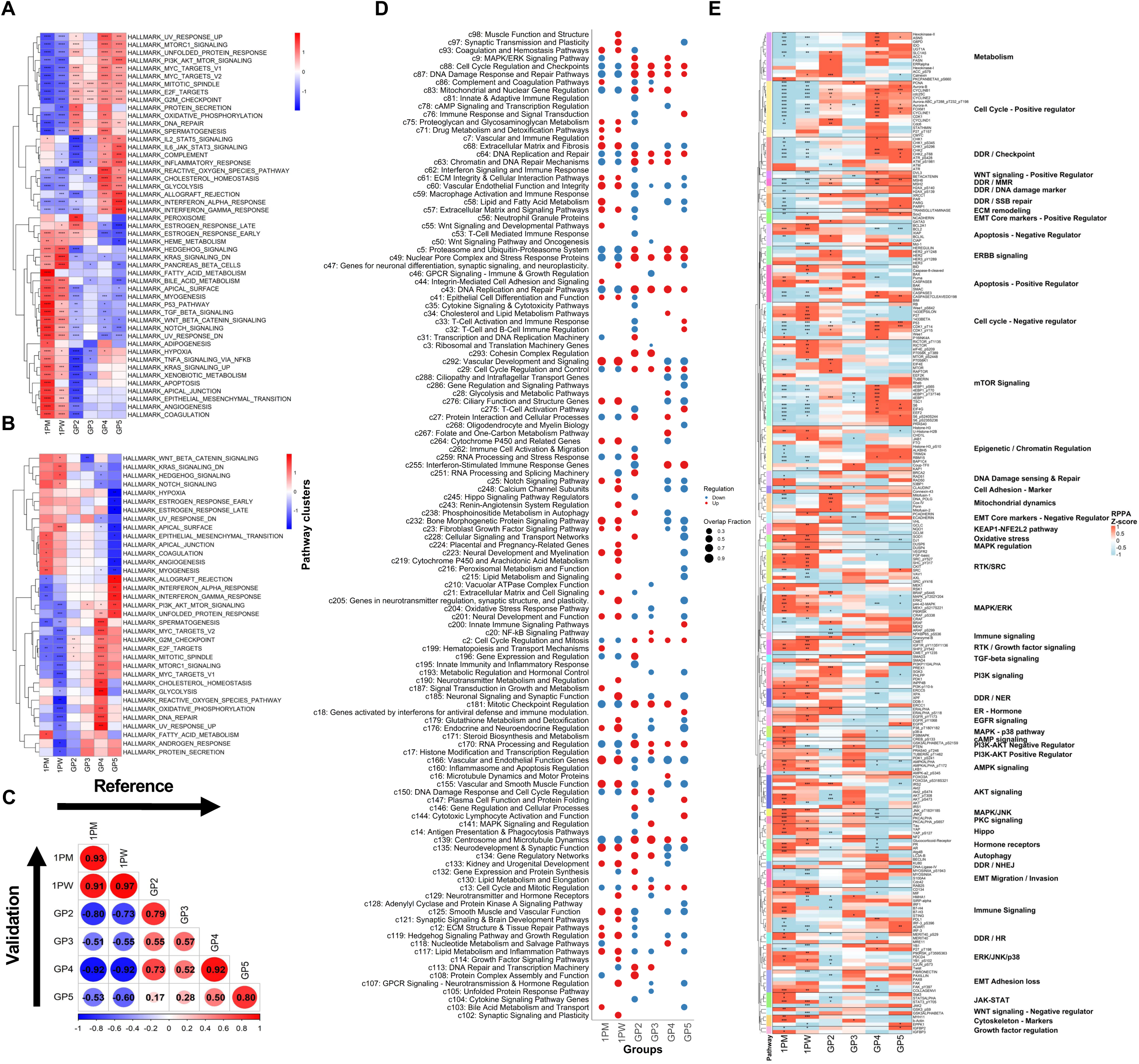
Integrated GSVA pathway and proteomic profiling reveal Tier 2 subgroup-specific tumour biology and targetable vulnerabilities in ER+/HER2– breast cancer. **(A)** Heatmap shows the scaled mean GSVA scores of Hallmark pathways across subgroups in the complete reference ER+/HER2– BRCA dataset. Each column corresponds to a Tier 2 BRCA subgroup, and each row represents a Hallmark pathway. **(B)** Heatmap shows the scaled mean ssGSEA score of Hallmark pathways across subgroups in the Metastatic sample-specific dataset only (n=102). In Figures A and B, statistical significance is annotated as: *adj p < 0.0001 (****), adj p < 0.001 (***), adj p < 0.01 (**), adj p < 0.05 (*)*. **(C)** Correlation plot showing strong concordance between validation and reference datasets across breast cancer subgroups. Pathway activation fold differences were independently calculated for each subgroup relative to all others using an equal-weighted approach in both the reference and validation cohorts (cohorts detailed in *Supplementary Table 1*). Pathways significantly activated in at least one subgroup of the validation dataset (p < 0.05) were selected. Correlations between the fold-difference values of these significant pathways in the validation and reference datasets are displayed. **(D)** Dot plot illustrating pathway clusters enriched within each subgroup. Each column represents a “one-vs-the-rest” comparison (e.g., 1PM vs all other BRCA subgroups). Red dots indicate upregulation and blue dots indicate downregulation. Pathway clusters were derived using a structured, reproducible annotation workflow, followed by enrichment testing using Fisher’s exact test (*see* Methods). **(E)** RPPA-based heatmaps show mean protein/phosphoprotein levels across the Tier 2 BRCA subgroups, with significance markers *p-value < 0.0001 (****), p-value < 0.001 (***), p-value < 0.01 (**), p-value < 0.05 (*),* indicating associations between each protein and the corresponding subgroup compared with the remaining samples. Proteins were annotated using curated pathway mappings, grouped by pathway, and classified by their expected functional effect in breast cancer (positive vs negative regulators); these annotations are displayed alongside the heatmap.

To reduce redundancy across >10,000 GSVA gene sets, we constructed a pathway–pathway similarity metric based on hypergeometric gene overlap and clustered pathways into higher-order biological modules (Methods). Subgroup enrichment was summarised at the module level to generate a global map of themes differentially engaged across six Tier 2 subgroups (**Figure 7D; Tables S10,** and **S11**), including immune homeostasis/recall, epithelial differentiation–adhesion/ECM mechanics, oxidative–lipid metabolism/detoxification, proteostasis/UPR, apoptosis–senescence, chromatin/transcriptional regulation, and chemotaxis/retention signalling.

In parallel, RPPA profiling demonstrated that the six Tier 2 subgroups differ not only at the transcript level but also at the levels of total protein abundance and phospho-signalling (**Figures 7E** and **S32**), supporting that the inferred programmes correspond to distinct functional tumour states. To move beyond a descriptive module map and define the biology that distinguishes each state, we integrated concordant RNA–protein patterns with structured, literature-informed pathway annotations. This analysis showed that Tier 2 subgroups share a common genomic-risk backbone but diverge through subgroup-specific signalling circuitry, metabolic organisation, and immune-control architecture, revealing vulnerabilities not captured by proliferation readouts alone. The defining features, sentinel biomarkers, and therapeutic implications for each subgroup are summarised in **Table 1**, with the core regulatory logic depicted as mechanism-of-action schematics in **Figures S33-S35**. A detailed subgroup compendium—including pathway annotations, marker lists, and the concordant RNA–protein features supporting these assignments—is provided in **Supplementary Appendix A** for deeper interrogation of subgroup wiring and candidate dependencies, with the defining features highlighted below.

**Table 1.** Summary table highlighting Tier 2 subgroup-specific tumour biology revealed by integrated GSVA and proteomic profiling. The defining features, sentinel biomarkers, and therapeutic implications for each subgroup are summarised.

| Subgroup | Biological Identity | Dominant Activated Programmes | Suppressed Context | Key Biomarkers (examples) | Major Therapeutic Liabilities |
| --- | --- | --- | --- | --- | --- |
| **1PM** | Stress-policed FAO–AMPK–Hippo quiescent luminal state | Autophagy, FAO, Hippo restraint, stress MAPK | mTORC1, translation, replication stress | YAP pS127, AMPK pT172, ATG4B/5, PDCD4, B7-H3/H4 | Autophagy/ metabolic stress targeting |
| **1PW** | Neuroendocrine/GPCR–RTK luminal, signalling-on but brake-engaged | ER/RTK–MAPK, feedback phosphatases | Cell-cycle, mTOR output | ERα, IGF1Rβ, DUSP4/6, WEE1 pS642, EGFR pY1173/1068 | Endocrine + MAPK adaptor targeting |
| **GP2** | ER–RTK–metabolic translation-addicted high-risk | mTOR–translation, proteostasis, OXPHOS, ATR–CHK1 | ECM/immune signalling | RAPTOR, EIF4E, FASN, ACC1, SLC1A5, Mitofusin 1/2 | ER+RTK+mTOR, ATR/CHK1, metabolic targeting |
| **GP3** | PI3K–AKT active, mTORC1-restrained, Rho/FAK invasive state | AMPK, autophagy, Rho GTPase, DNA repair | RTK signalling | AKT, AMPKα, STING, Puma, CD4, ATG3, FAK pY397 | PI3K/AKT + autophagy blockade, Rho/FAK |
| **GP4** | TP53-mutant, translation-addicted, replication-stressed | mTOR–eIF4F, DDR checkpoints, glycolysis/PPP | Differentiation, nuclear receptor control | 4EBP1 pS65, S6 pS240/244, CHK1 pS345, ATR pS428, G6PD | ATR/CHK1/ WEE1, translation, redox targeting |
| **GP5** | IFN-wired, checkpoint-primed, redox-compromised | IFN/TLR/NFκB, PI3K survival, DDR | MAPK, detoxification pathways | ADAR1, PD-L1, 4EBP1, CDK1 pT14, ATR pY428, RPA32 pS4/S8, PARP1 | Immunotherapy + DDR + ROS/ferroptosis |

#### 8.1 Low Genomic Score States (1PM and 1PW)

Low-genomic-score groups 1PM and 1PW correspond to restrained luminal states that achieve favourable prognosis, though via different regulatory logics — stress-policed FAO–Hippo quiescence in 1PM versus signalling-receptive, brake-engaged neuroendocrine–RTK wiring in 1PW (**Figure S33C**).

##### 1PM — Stress-policed FAO–Hippo quiescent luminal state (PIK3CA-mutant)

Our analyses highlight 1PM as a catabolic, checkpoint-restrained luminal state in which Hippo–AMPK signalling (evidenced by elevated YAP S127 phosphorylation, enrichment of multiple Hippo/YAP-restraining modules at the transcriptomic level, and AMPK T172 phosphorylation), autophagy, and fatty-acid oxidation (FAO) dominate over proliferative outputs. mTORC1/translation and replication-stress signals are suppressed, whereas stress-MAPK (increased total p38, total JNK2, p38-pT180/Y182, JNK-pT183/Y185) and p53-governed restraint programmes are prominent (**Figures 7A, 7E, S31, S32,** and **S48**). ECM/AXL-linked survival wiring and a “danger-experienced” innate-immune imprint are observed, suggesting durable stress adaptation rather than expansion. This configuration channels oncogenic PI3K signalling into homeostatic buffering, consistent with favourable outcomes, reduced prognostic impact of nodal involvement, and a longer time-to-metastatic presentation. The overall biology suggests metabolic stress/autophagy vulnerabilities as rational therapeutic targets rather than proliferation-centric targeting, despite *PIK3CA* mutation (**Figure S33A; Table 1**).

##### 1PW — Neuroendocrine/RTK–MAPK luminal state with brake-engaged proliferation (PIK3CA-wild-type)

We identified 1PW as a signal-receptive luminal state enriched for neuronal/GPCR programmes and RTK–MAPK signalling, yet constrained by feedback phosphatases (DUSP4/DUSP6) and checkpoint brakes (WEE1 pS642, p27). Upstream signalling is active, yet cell-cycle machinery and mTOR-driven translation remain suppressed, defining a “signalling-on, proliferation-off” configuration. Survival wiring further supports a “buffered, brake-engaged” state (high BCL2, DJ-1, MIF, PEA15, BCL2A1, Bid, 14-3-3 proteins, GCLC, SOD1, LKB1, and AceCS1, with low Bax and MCL1), implying greater anti-apoptotic and redox support (**Figures 7A, 7E, S31,** and **S32**). Compared with 1PM, stress-policing circuitry is less dominant, aligning with earlier metastatic progression despite low genomic risk (**Figures 6D** and **6E**). The endocrine backbone, with MAPK adaptor-targeting upon resistance, provides rational therapeutic logic (**Figure S33B; Table 1**).

#### 8.2 High Genomic Score States (GP2-GP5)

All high-genomic-score tumours share a dominant programme of cell-cycle activation and genome maintenance, with coordinated enrichment of E2F/G2M, DNA replication, ATR/ATM, homologous recombination, Fanconi, and checkpoint pathways, indicating proliferation under chronic replication stress. In parallel, developmental signalling, circadian regulation, stromal programmes, and physiological growth-factor and detoxification pathways are broadly suppressed, consistent with dedifferentiation, metabolic rewiring, and loss of tissue homeostasis as universal high-risk features. Although these shared pathways define the common high-score backbone, each high-genomic-score subgroup exhibits distinct, state-specific biological gains that reveal non-overlapping dependencies and actionable vulnerabilities (discussed below)—providing a rationale for precision therapeutic layering beyond backbone regimens (**Figures S34** and **S35; Table 1**).

##### GP2 — ER–RTK–anabolic/metabolic addiction in a luminal high-genomic-risk state (TP53-PIK3CA double wild-type)

GP2 combines strong ER/nuclear receptor signalling with HER2–IGF RTK input, mTORC1–translation/proteostasis activation, and lipogenic/OXPHOS metabolism (high FASN, ACC1, SLC1A5, Mitofusin-1/2, Cox-IV), in the context of relatively capped canonical AKT phosphorylation. ATR–CHK1–WEE1–skewed replication-stress responses are prominent, whereas ECM and adaptive immune programmes are attenuated, yielding an immune-quiet phenotype. Although mTOR output is high, it is driven by classical RTK→PI3K anabolic signalling, supporting biosynthetic expansion, alongside a disabled AMPK “circuit breaker” arm (lower total AMPK and AMPK pT172 levels). Biology suggests ER plus RTK blockade with mTOR/translation, metabolic, and ATR–CHK1 targeting as rational therapeutic strategies (**Figure S34A; Table 1**).

##### GP3 — PI3K–AMPK–autophagy-buffered, AKT-active but mTORC1-restrained invasive state (PIK3CA-mutant)

GP3 retains proliferative programmes but is distinguished by AKT activation with restrained mTORC1 output, AMPK-centred metabolic control, autophagy dependence, and Rho/FAK-driven cytoskeletal plasticity. Epithelial adhesion is reduced, defining an invasion-oriented phenotype (characterised by low levels of E-cadherin, β-catenin, claudin-7, DDR1, and higher COUP-TFII) (**Figures 7A, 7E, S31,** and **S32**). A PUMA-primed, stress-tolerant DDR network supports survival under genotoxic pressure. PI3K/AKT plus autophagy blockade, DDR exploitation, and Rho/FAK targeting provide a rational therapeutic strategy (**Figure S34B; Table 1**)

##### GP4 — TP53-mutant, stress-coupled translation-addicted state

GP4 is characterised by mTOR–eIF4F translation, intense replication-stress checkpoint signalling (high ATR pS428, CHK1 pS345, CHK2 pT68, RPA32 pS4/S8, CDK1 pT14/pY15, CDC25C, PCNA), glycolysis/PPP flux, and NRF2-driven antioxidant buffering. Here, mTOR/translation supports survival in a TP53-mutant, genome-unstable context rather than in a pure growth-factor-addiction context. Developmental and nuclear receptor circuits are suppressed, resulting in a highly dedifferentiated phenotype. ATR/CHK1/WEE1 and PARP targeting, translation inhibition, and redox/ferroptosis strategies provide a strong therapeutic rationale (**Figure S34C; Table 1**).

##### GP5 — Interferon-wired, checkpoint-primed, redox-vulnerable state (TP53–PIK3CA double mutant)

GP5 shares the DDR backbone but is uniquely enriched for nucleic-acid sensing, type I interferon, TLR/NF-κB, and JAK–STAT signalling, with strong immune-checkpoint signatures (explicit enrichment of several immune-checkpoint/immunotherapy signatures). MAPK tone is low, whereas PI3K-linked survival routes (enrichment of BCR/BCAP/CD19–PI3K, PDGF–PI3K, ERBB–PI3K, CXCR4–PI3K, PD-1/PD-L1–SHP–PI3K modules) and translation nodes remain active. Here, mTOR operates within an immune/inflammatory stress context, not classical RTK addiction. Loss of detoxification and mitochondrial flexibility indicates redox vulnerability. Survival is sustained more by PI3K–JAK–STAT–NF-κB and translation/DDR modules than by a strong MAPK/ERK dependency. Immunotherapy combinations that target DDR and exploit oxidative/ferroptotic stress are rational therapeutic strategies (**Figure S34D; Table 1**).

Together, these data show that genomic risk or mutation status alone does not define tumour biology. Instead, Tier 2 subgroups represent programme-defined tumour states in which proliferation output is decoupled from signalling context, metabolic wiring, and immune engagement. While replication-stress/DDR targeting is a shared vulnerability across high-risk disease, additional liabilities are subgroup-specific: RTK-driven anabolic dependence (GP2), autophagy–cytoskeletal plasticity (GP3), stress-coupled translation/redox buffering (GP4), and interferon-driven immune/redox vulnerabilities (GP5). This framework enables mechanistically grounded precision therapeutic stratification beyond single-gene biomarkers.

### 9. Association of Tier 2 Subgroups with Clinicopathological Characteristics and PAM50 Subtyping

We assessed relationships between the six Tier 2 molecular subgroups and established clinicopathological features, including PAM50 intrinsic subtypes. Tier 2 subgroups differed significantly in the distribution of PAM50 subtypes, Nottingham Prognostic Index (NPI), tumour size, tumour stage, and histological grade (all p < 0.001; **Tables S9** and **S12**), indicating that the DNA–RNA–defined stratification captures heterogeneity across conventional descriptors rather than recapitulating any single clinical variable.

Standardised Pearson residuals (**Figure S36**) highlighted structured enrichment patterns. Basal-like and HER2-enriched features were predominantly enriched in GP4/GP5, whereas Luminal B characteristics were overrepresented in GP2/GP3. Luminal A features were preferentially enriched in the low-score subgroups (1PM/1PW), with differential enrichment between these classes, consistent with refined stratification within genomically low-risk disease.

Clinicopathological distributions showed a concordant subgroup structure. Tumours <2 cm and with low NPI (NPI < 4) were enriched in 1PM, whereas higher-risk NPI (NPI > 4) was most frequent in GP2/GP4/GP5. Histological grade showed a similar pattern: Grade 1 tumours were enriched in 1PM, Grade 2 in 1PW, and Grade 3 in GP5/GP4/GP2. Tumour stage did not show a consistent subgroup-specific enrichment pattern. Notably, unlike other Tier 2 subgroups, GP3 did not exhibit enrichment for NPI, histological grade, tumour size, or tumour stage, suggesting that its biology is not well captured by conventional clinicopathological risk features and may represent a distinct programme state with clinicopathologically heterogeneous presentation (**Figure S36)**. Further, although 1PM and 1PW are both low-score genomic states, 1PM was more consistently associated with favourable clinicopathological features.

Cramér’s V indicated low concordance with tumour size (V = 0.134) and stage (V = 0.112), but moderate associations with PAM50 subtype (V = 0.385), NPI (V = 0.268), and grade (V = 0.291), suggesting partial overlap with intrinsic subtype and aggressiveness while preserving independent stratification beyond conventional clinical markers **(Figure 8)**.

**Figure 8.**
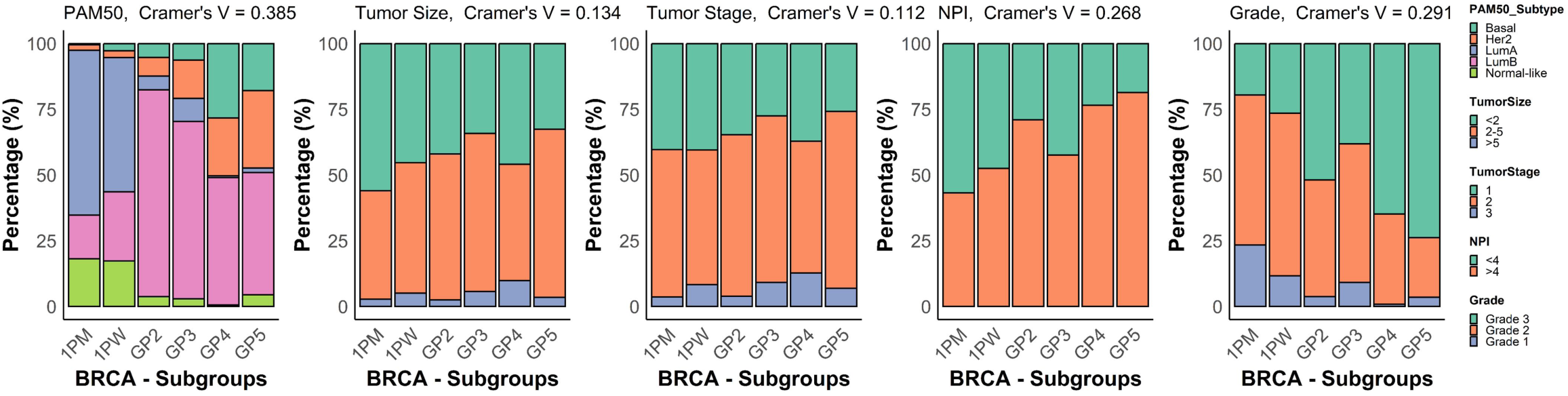
Association of Tier 2 subgroups with clinicopathological characteristics and PAM50 subtyping. The Tier 2 BRCA subgroups were evaluated against established clinicopathological variables, including PAM50, tumour size, tumour stage, Nottingham Prognostic Index (NPI) and histological grade, and are shown as percentage-based stacked histograms. Cramér’s V coefficients, quantifying the strength of association between each clinical variable and Tier 2 BRCA subgroups, are shown alongside each plot.

### 10. Tier 3 Stratification Reveals Within-Subgroup Risk Gradients and Clinically Relevant Intermediate-Risk Subsets

Although Tier 2 stratification into six molecular subgroups (1PM, 1PW, GP2–GP5) provided programme-level resolution, we observed substantial heterogeneity in pathway activity and clinical behaviour within each subgroup (**Figure S38B**). We therefore implemented a Tier 3 refinement by dichotomising tumours within each Tier 2 subgroup using the median DNA-anchored RNA composite genomic score for that subgroup, thereby generating within-subgroup low- and high-activity strata.

This intra-subgroup stratification uncovered clinically relevant intermediate-state subsets and reinforced the gradient nature of tumour biology within each Tier 2 programme. Among tumours assigned to nominally high-genomic-risk Tier 2 classes (GP2–GP5), a subset exhibited a biologically “quieter” high-genomic-risk state (GP2A, GP3A, GP4A, and GP5A), with attenuated activation of canonical cell-cycle/replication programmes (e.g., reduced enrichment of E2F/G2M and DNA replication/DDR gene sets/modules) and concordant dampening of proliferation/replication-stress proteins (e.g., lower Cyclin B1/E1, FOXM1, CDC25C, PCNA, reduced CDK1 pT14/pY15, PARP1, and reduced phosphorylation of checkpoint nodes such as ATR–CHK1/CHK2) (**Figures 9A, 9B, S37A, S37B,** and **S38**). Conversely, within nominally low-genomic-risk Tier 2 groups (1PM and 1PW), a subset showed higher-than-expected activity (1PMB, 1PWB), with increased cell-cycle/DNA replication output accompanied by relaxation of key brakes (p27 and WEE1 pS642 decline). Importantly, Tier 3 progression was not limited to proliferation but also included coherent, subgroup-specific cell-cycle–independent state shifts that tracked with activity. For example, GP2 showed graded increases in lipogenic provisioning (FASN, ACC/ACC1 pS79); GP3 showed progressive strengthening of innate sensing and plasticity wiring (STING, COUP-TFII, N-cadherin) with increased DNA damage tone (H2AX pS139); GP4 showed activity-linked increases in immune-evasion/interaction (PD-L1, ADAR1/SIRPα) alongside reduced energy-sensing tone (total/phospho-AMPK) and ERα; and GP5 showed increased escape/stemness and adaptive wiring (SOX2, IGFBP2/ADAR1) with dampened MAPK output (p44/42 MAPK/MAPK pT202/Y204) and reduced AMPK (**Figure S37B**). Transcriptomic modules similarly exhibited graded non–cell-cycle–centric shifts across subgroups (**Figures 9A, 9B, S37A,** and **S38)**. Together, these patterns support Tier 3 as a biologically grounded, within-subgroup measure of tumour “activity/state” that integrates proliferative output with programme-specific adaptation circuitry.

**Figure 9.**
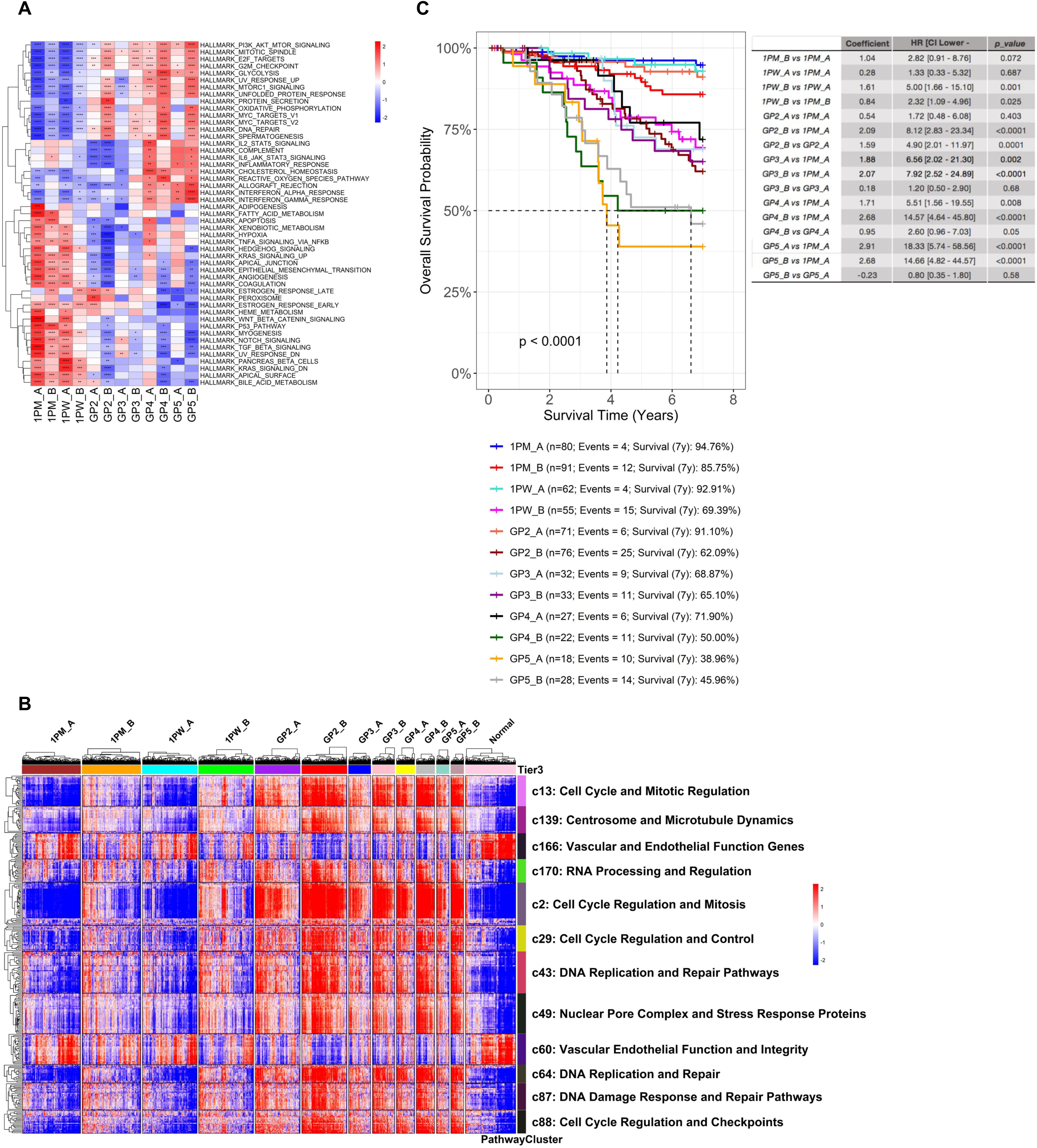
Tier 3 stratification reveals within–Tier 2 risk gradients and improves biological resolution. Tier 3 is defined by dichotomising tumours within each Tier 2 subgroup using the subgroup-specific median genomic risk score (DNA-anchored RNA composite score). **(A)** Heatmap of scaled mean GSVA Hallmark pathway scores across Tier 2 subgroups, further partitioned by Tier 3 (low vs high within-subgroup genomic score). Columns represent Tier 3 strata within each Tier 2 subgroup; rows represent Hallmark pathways. Statistical significance is annotated as: *adj p < 0.0001 (****), adj p < 0.001 (***), adj p < 0.01 (**), adj p < 0.05 (*)*. Suffix A denotes a below-median score; B denotes an above-median score. **(B)** Kaplan–Meier analysis showing improved risk discrimination with Tier 3 stratification among lymph-node–positive ER+/HER2− patients (≥1 positive node) in the METABRIC cohort (n=595). The y-axis shows the 7-year survival probability; patients with follow-up beyond 7 years were censored. The accompanying table reports pairwise comparisons versus the indicated reference group (regression coefficient, HR with 95% CI, and *p*-value). Plot annotations indicate the number of patients per group, the number of events, and the 7-year survival estimate. **(C)** Sample-level heatmaps of GSVA enrichment scores for canonical cell-cycle/replication pathway clusters across Tier 3 strata. These highlight attenuated activation of cell-cycle–associated programmes in a subset of tumours representing a biologically “quieter” state within nominally high-risk Tier 2 classes (GP2A, GP3A, GP4A, GP5A).

Clinically, Tier 3 refinement improved risk discrimination in lymph-node–positive disease. In particular, Tier 3 strata within 1PM separated a consistently favourable subset (1PMA) with >90% 10-year post-diagnosis survival probability from a relatively higher-activity 1PM subset (1PMB), which still showed significantly better outcomes than 1PWB and high-risk subgroups. Together, these results suggest that Tier 3 stratification can identify a sizeable subset of node-positive ER+/HER2− patients (**Figure 9C** and **S39A-S39C**) with favourable biology aligned with the 1PM programme, supporting confident risk calibration and potential de-escalation considerations, while also flagging within-programme higher-activity tumours that may warrant closer monitoring or intensified targeted strategies.

### 11. Tier 4 Mutation Overlay Reveals Subgroup-Specific Roles of Recurrent Drivers and Enables Modular Refinement Beyond Proliferative Risk

We then introduced Tier 4 as a modular mutation overlay to capture recurrent Group B alterations (non-canonical, comparatively non-proliferative) whose contributions to tumour behaviour and prognosis are not redundant with the Tier 1 DNA-anchored RNA score, which summarises the dominant canonical proliferative/replicative output and the associated genomic-risk backbone.

Consistent with prior reports that CDH1 mutations lack clear prognostic value in unstratified invasive breast cancer cohorts [38], CDH1 was not a strong prognostic marker across pan-cohort analyses (**Figure S40A**). However, within defined programme and activity contexts—particularly in low-genomic-score tumours—CDH1 mutations were associated with significantly worse outcomes despite no significant differences in RNA-based genomic scores, revealing score-independent adverse biology that is masked by population-level averaging or genomic-score stratification alone (**Figures 10A** and **S40A-S40D**). MAP3K1 similarly showed strong context dependence. In low-score, node-negative, hormone receptor–positive patients, MAP3K1 mutations were associated with >90% long-term survival even without systemic therapy (**Figure 10B**), raising the possibility of overtreatment in this setting. In node-positive low-score disease, MAP3K1 also marked a relatively favourable subset (**Figure 10C**). We further observed favourable outcomes for GATA3-mutant hormone receptor–positive tumours across low- and high-score disease subgroups (**Figures 10D** and **S41A**), and modest but statistically significant prognostic refinement for KMT2C in low-score hormone receptor–positive tumours (**Figures S41B** and **S41C**).

**Figure 10.**
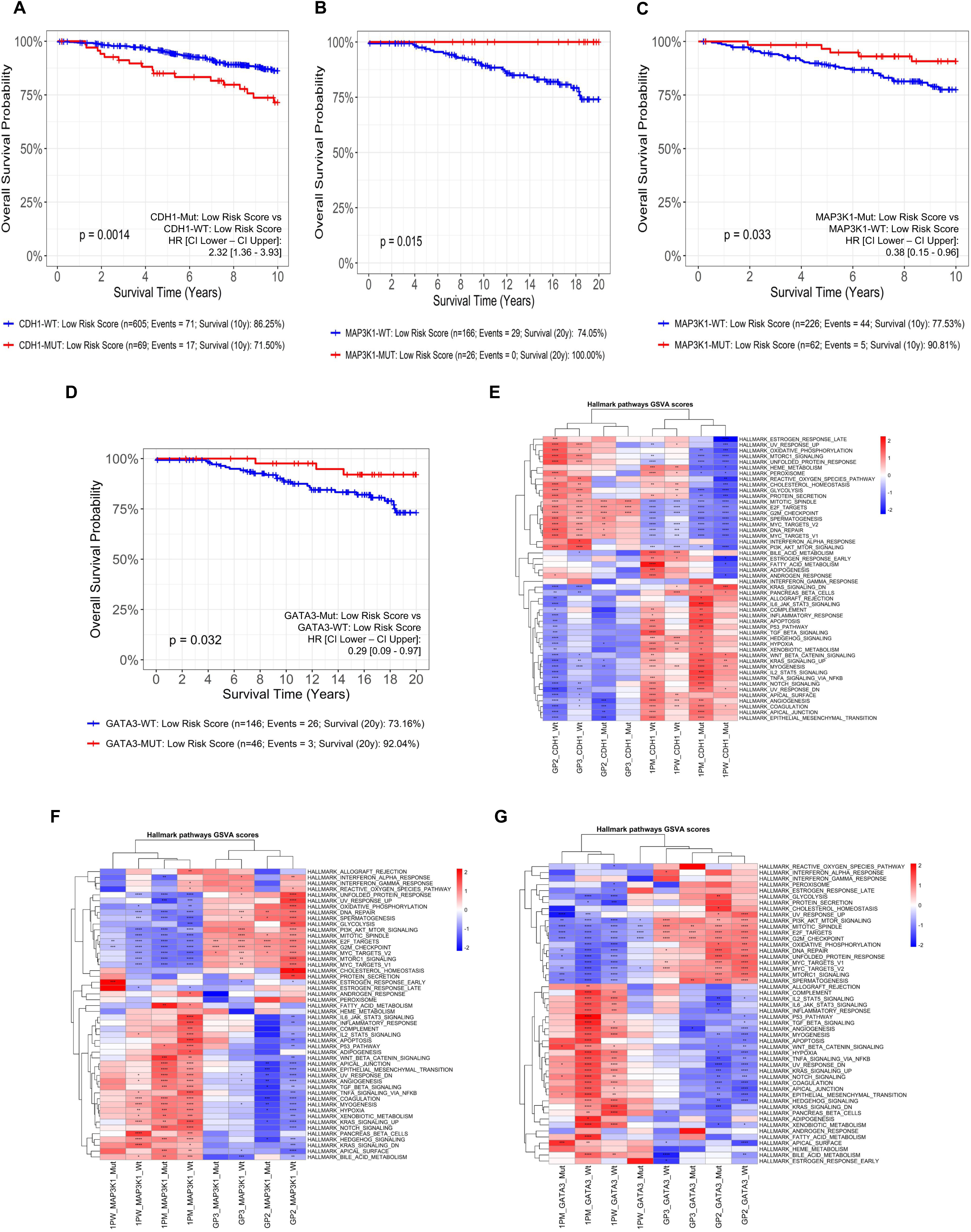
Tier 4 applies complementary, non-redundant mutation overlays to refine prognosis and biology within the T-OMICS framework. **(A–D)** Kaplan–Meier (KM) analyses in ER+/HER2− patients from METABRIC show that Tier 4 modifier mutations, CDH1, MAP3K1, and GATA3, exhibit subgroup-specific prognostic associations. **(A)** CDH1 mutation is associated with a worse outcome in the lymph-node–negative, Tier 1 low-genomic-score subgroup. **(B–C)** MAP3K1 mutation is associated with a favourable outcome in the lymph-node–negative, Tier 1 low-genomic-score subgroup without systemic therapy (B) and in lymph-node–positive patients (C). **(D)** GATA3 mutation is associated with a favourable outcome in the lymph-node–negative, Tier 1 low-genomic-score subgroup without systemic therapy. The y-axis shows breast cancer–specific overall survival probability at 10 or 20 years (as indicated). Patients with follow-up beyond the plotted time horizon were censored. **(E–G)** Heatmaps of scaled mean GSVA Hallmark pathway scores across Tier 2 subgroups, further stratified by Tier 4 mutation overlays: CDH1 (E), MAP3K1 (F), and GATA3 (G). Columns represent Tier 4 mutation status within each Tier 2 subgroup; rows represent Hallmark pathways. Significance is annotated as: *adj p < 0.0001 (****), adj p < 0.001 (***), adj p < 0.01 (**), adj p < 0.05 (*)*. Column-wise hierarchical clustering shows that Tier 4 mutation overlays remain transcriptionally aligned with their parent Tier 2 subgroup, consistent with modifier effects that refine phenotype without redefining core identity.

Together, these findings indicate that selected recurrent non-canonical mutations provide prognostic and biological information independent of the proliferative score, justifying Tier 4 as a non-redundant modifier layer. Several additional mechanistically relevant alterations (e.g., ESR1, AKT1, PTEN) were too infrequent in these cohorts to enable robust subgroup-specific survival testing, but can be incorporated as datasets expand. Finally, tumours harbouring Tier 4 mutations remained transcriptionally aligned with their parent Tier 2 subgroup, indicating preserved core programme identity (**Figures 10E-10G** and **S42A-S42C**). Instead, GSVA and protein/phosphoprotein profiles (**Figures S43A-S43C,** and **S44A-S44C**) suggested subtle shifts in pathway activity, consistent with Tier 4 events functioning as signalling modifiers that tune phenotype, plasticity, and resistance liabilities within a stable programme/state framework.

To extract genotype-driven biology from subgroup composition effects, we performed within-subgroup GSVA, comparing mutant tumours with subgroup-matched wild-type counterparts, and integrated RPPA protein and phospho-protein data to identify conserved mutation-associated programmes that persist across programme contexts.

#### CDH1 Mutation–Associated Programmes (Adhesion–Plasticity and Inflammatory-Lipid Modifier)

Across subgroups, CDH1 mutation was associated with a conserved modifier programme characterised by loss of epithelial cohesion and increased plasticity, supported by reduced junctional/polarity proteins (e.g., E-cadherin/Claudin-7/β-catenin/NF2) (**Figures S43A** and **S44A**). This state was distinguished by inflammatory lipid conditioning with enrichment of arachidonic acid/eicosanoid metabolism modules, while bulk mTORC1–glycolysis outputs were comparatively attenuated. Despite this low-anabolic profile, survival signalling remained engaged via RTK–STAT3 wiring (increased EGFR/HER3/IGF1R phosphorylation with STAT3 pY705) and receptor recycling machinery (RAB11/RAB25), with FOXO-linked stress adaptation, consistent with score-independent adverse biology in defined low-score contexts. These features nominate RTK→STAT3-centred combinations, apoptosis-execution release strategies, and context-appropriate modulation of eicosanoid signalling as rational hypotheses (**Figure S45A**).

#### MAP3K1 Mutation–Associated Programmes (Metabolic-Fitness and Growth-Restraint Modifier)

Across subgroups, MAP3K1 mutation was associated with a metabolically oxidative, growth-restrained modifier programme enriched for mitochondrial fatty-acid oxidation, BCAA catabolism/urea-cycle pathways, and glutathione recycling, consistent with substrate flexibility and redox resilience. Proteomic profiles supported mitochondrial fitness and autophagy maintenance (e.g., MFN2/ATG3) and showed reduced levels of the mitotic and replication-stress machinery (e.g., CDK1/FOXM1/AURORA-A/ATR). Although pERK (MAPK pT202/Y204) was increased, upstream RTK/RAS–MAPK components were comparatively reduced (EGFR/EGFR pY1173, SHP2, NRAS) (**Figures S43B** and **S44B**), with enrichment of MAPK negative-feedback programmes, suggesting tonic ERK output under feedback constraint rather than MAPK addiction; immune cytotoxic features were also attenuated. Clinically, this programme aligns with indolent biology and supports endocrine backbone therapy with careful escalation (CDK4/6 inhibitors as the most aligned option in advanced disease, given the strong cell-cycle–control context), while monitoring PI3K–AKT–mTOR adaptation and considering FAO/redox/autophagy vulnerabilities in endocrine-combination trials (**Figure S45B**).

#### GATA3 Mutation–Associated Programmes (Lineage-Fidelity and “Contained Growth” Modifier)

GATA3 mutation marked a conserved “contained growth” modifier programme characterised by luminal lineage stabilisation, restrained invasive plasticity, Hippo/YAP-associated restraint (NF2-high), and preservation of epithelial organisation. Canonical KRAS/MAPK output and RTK bypass dependence were dampened (reduced MEK phosphorylation and lower EGFR/cMET/IGF1R activation), consistent with dominant endocrine dependence at baseline and favourable clinical behaviour across contexts (**Figures S43C** and **S44C**). At the same time, GATA3-mutant tumours showed persistent buffering via PPP/redox metabolism with mitochondrial/translation support (TFRC↑, TRAP1↑, MTCO1↑; S6↑, EEF2↑) and an elevated apoptotic threshold (MCL1-high/BAK-low), suggesting survival support without major proliferative escalation. This biology supports endocrine-anchored management with biomarker-triggered escalation at resistance, prioritising BCL2-family strategies and, where appropriate, metabolic/redox or mTOR/translation-axis targeting (**Figure S45C**).

Together, these analyses indicate that Tier 4 mutations do not redefine tumour identity but instead act as functional modifiers that tune adhesion, lineage stability, stress signalling, or plasticity within established programme and state frameworks. Unlike the shared proliferative backbone captured by the RNA score, these genotype-linked programmes provide additional resolution in predicting behaviour and therapeutic vulnerability.

### 12. Paired-sample T-OMICS profiling in metastatic disease reveals predominant Tier 2 truncal stability, frequent Tier 3 state transitions, and mixed stability versus acquisition of Tier 4 modifiers

To assess longitudinal applicability in metastatic ER+/HER2− breast cancer, we applied T-OMICS to 50 patients with paired specimens (primary–metastatic, metastatic–metastatic from distinct sites, or temporally separated paired primaries). Across patients, T-OMICS revealed substantial within-patient conservation of core molecular identity, with a quantifiable subset showing evolutionary divergence. Tier 2 subgroup assignment was concordant across all profiled lesions in 38/50 patients (76%), supporting Tier 2 as a largely truncal, patient-specific programme that is typically stable across sites. In contrast, 12/50 (24%) showed Tier 2 discordance, indicating programme-level divergence in a subset (**Figure 11)**. Notably, among these 12 discordant patients, 7 also exhibited a Tier 1 risk-group shift, transitioning between low (1PM/1PW)- and high-genomic-risk (GP2-GP5) classifications across lesions.

**Figure 11.**
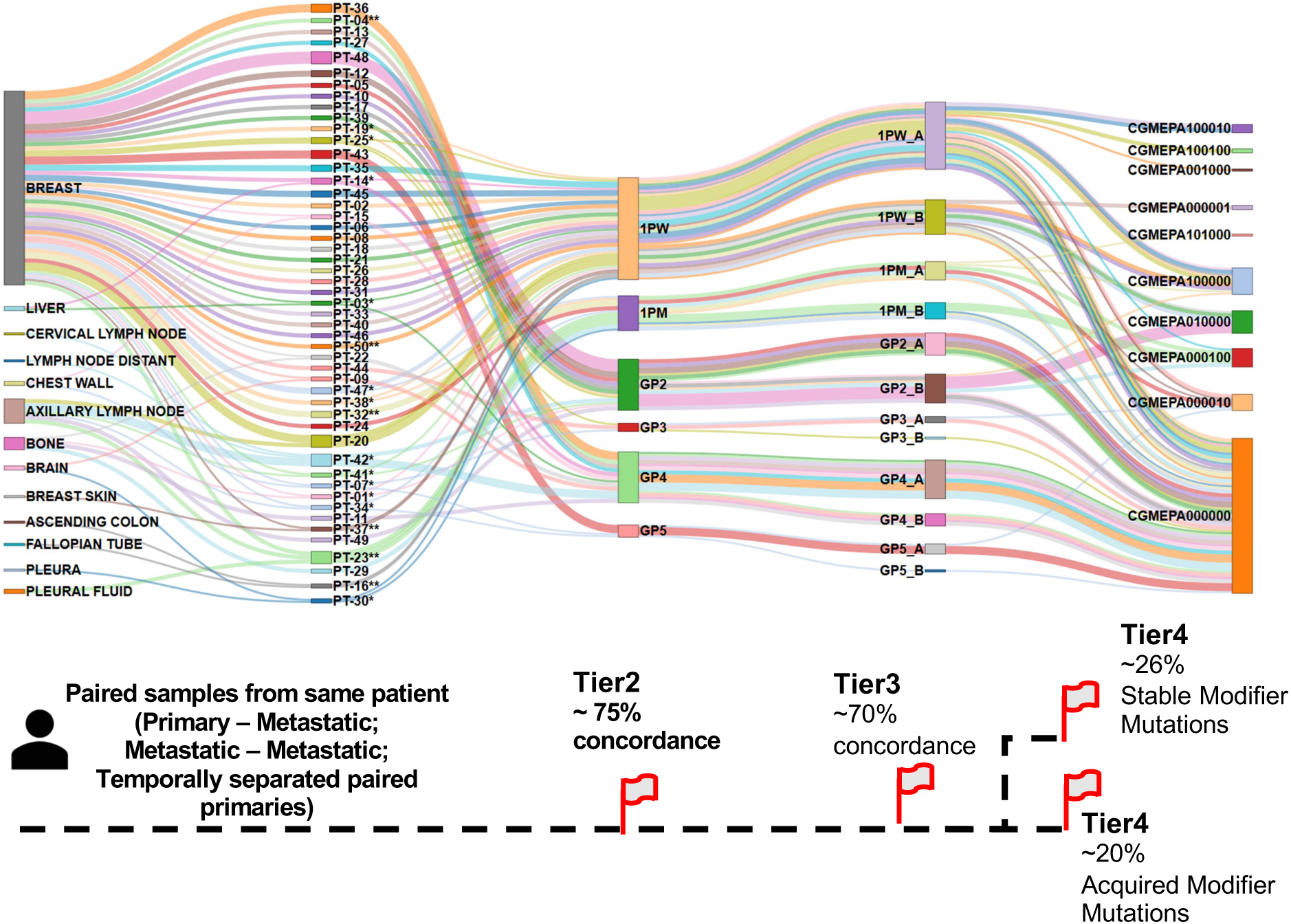
Paired-sample T-OMICS profiling across metastatic sites shows predominantly stable Tier 2 identity with lesion-level state and modifier heterogeneity. The alluvial plot summarises T-OMICS classifications across paired and multi-sample metastatic cases, displaying sample anatomic site (e.g., liver, bone, lymph node), patient identity, and tiered assignments (Tier 2, Tier 3, and Tier 4). Each patient is shown in a distinct colour; ribbons connect samples from the same patient across tiers. Tier 2 denotes the core functional subgroup (“identity”), Tier 3 denotes the within-subgroup activity/aggressiveness state (defined relative to the Tier 2 subgroup–specific median RNA score), and Tier 4 denotes modifier mutation overlays. Most patients show concordant Tier 2 identity across sampled lesions (76%), whereas a subset shows Tier 2 discordance across sites, consistent with inter-lesion programme divergence in some cases. In contrast, Tier 3 state and Tier 4 modifier overlays vary more frequently across lesions, indicating that RNA-defined tumour state and modifier events vary across lesions, including within patients with conserved Tier 2 identity. Patients exhibiting Tier 2 discordance across lesions are marked with an asterisk (*). Among Tier 2–concordant patients, those exhibiting Tier 3 discordance (state variation within a stable Tier 2 identity) are marked with a double asterisk (**).

Tier 3 discordance (within-identity activity/aggressiveness state) was observed in 6/38 Tier 2–concordant patients (16%), consistent with state variation within a stable core programme. Among the twelve Tier 2–discordant patients, lesions frequently differed in both Tier 2 identity and RNA score state; however, these differences reflect programme divergence with accompanying state heterogeneity, rather than within-identity Tier 3 transitions. Overall, Tier 2 captured a largely truncal identity across lesions, whereas the RNA-defined state was more plastic across metastatic sites.

At the Tier 4 mutation overlay level (CDH1, GATA3, MAP3K1, ESR1, PTEN, AKT1), two non-overlapping patterns emerged. First, 13/50 patients (26%) harboured a pre-existing Tier 4 mutation in at least one of these genes, and these alterations were stable across all additional specimens from the same patient (i.e., no further Tier 4 changes were detected with expanded sampling), consistent with truncal modifiers in this subset. Second, 10/50 (20%) showed acquisition or variability of Tier 4 mutations across lesions, with at least one specimen carrying additional Tier 4 events absent in another, suggesting lesion-to-lesion divergence and therapy adaptation in a minority (**Figures 11** and **S46**). Overall, ∼80% (40/50) showed no further acquisition of mutations in this Tier 4 gene set across paired profiling (Tier 4 mutations absent or stable). CDH1 was the most frequently altered Tier 4 gene in both settings (5/13 stable; 5/10 variable), followed by PTEN among stable cases (5/13) and ESR1 among variable/acquired cases (3/10). Tier 4 patterns occurred in both Tier 2–concordant and Tier 2–discordant patients, indicating that modifier stability or acquisition can occur with or without programme-level divergence.

Collectively, these data support T-OMICS as a longitudinal decision framework for metastatic disease, in which Tier 2 typically reflects stable tumour identity, Tier 3 captures more dynamic state transitions, and Tier 4 modifiers are usually stable but can evolve in a subset, thereby identifying patients most likely to benefit from multi-site profiling. Consistent with their design, Tier 4 alterations generally preserve Tier 2 identity while potentially shifting pathway dependencies, resistance liabilities, and context-specific prognosis.

## Discussion

Comprehensive genomic profiling is now routine in breast cancer, yet interpretation has lagged behind measurement because there is no organising framework that links complex constellations of mutations and copy-number alterations to coherent tumour-level biology. Clinicians are often presented with multi-lesion reports without guidance on how alterations collectively define behaviour, prognostic context, or therapeutic priorities. Interpretation therefore defaults to mutation-first matching (matching individual genes to drugs) or indirect inference of “pathway activation” from mutational/CNA proxies—approaches that frequently miss the integrated programme and state of the tumour cell [4,5]. This gap is especially evident in ER+/HER2− disease, where common alterations recur across a broad biological spectrum and show inconsistent prognostic associations when considered in isolation [13,39]. Here, we address this barrier by introducing a programme-oriented framework that translates genomic complexity into clinically legible tumour states.

A central conceptual advance is the identification of a robust functional organisation underlying breast cancer genomic heterogeneity. Across cohorts, recurrent aberrations segregate into two dominant mechanistic classes: a canonical proliferative/replicative programme, exemplified by TP53-linked genomic instability and copy-number–rich landscapes, and a complementary class of non-canonical alterations—including PIK3CA, CDH1, GATA3, MAP3K1, ESR1, and AKT1—that primarily modify signalling and cell state rather than acting as direct proliferative accelerators. This organisation aligns with population-scale observations linking CNA burden and TP53-associated instability to aggressive phenotypes and showing enrichment of luminal/lineage-associated alterations within ER+ disease [1–3], yet prior work remained largely descriptive. We formalise this latent structure by demonstrating that these alteration classes map to opposing programmes defined by downstream molecular directionality and clinical behaviour.

We translated this discovery into T-OMICS, a tiered framework aligned with clinical decision-making needs (**Figure 12**). Tier 1 captures dominant canonical proliferative/replicative output via a DNA-anchored RNA signature that serves as a tumour-level readout of the copy-number–rich proliferative landscape, rather than a surrogate for any single mutation. This provides a mechanistically grounded continuous risk backbone complementary to established expression-based assays and intrinsic subtyping frameworks [22,40], while explicitly integrating genomic configuration. Tier 2 assigns programme identity to six molecular subgroups, separating core tumour system configurations that may share proliferative risk yet diverge through distinct signalling, metabolic, and immune control architectures. Tier 3 quantifies within-programme activity and adaptation, recognising that tumours occupy a continuum rather than binary risk bins. Tier 4 overlays recurrent non-canonical modifiers whose effects are not redundant with proliferative output and which refine phenotype, vulnerabilities, and resistance liabilities.

**Figure 12.**
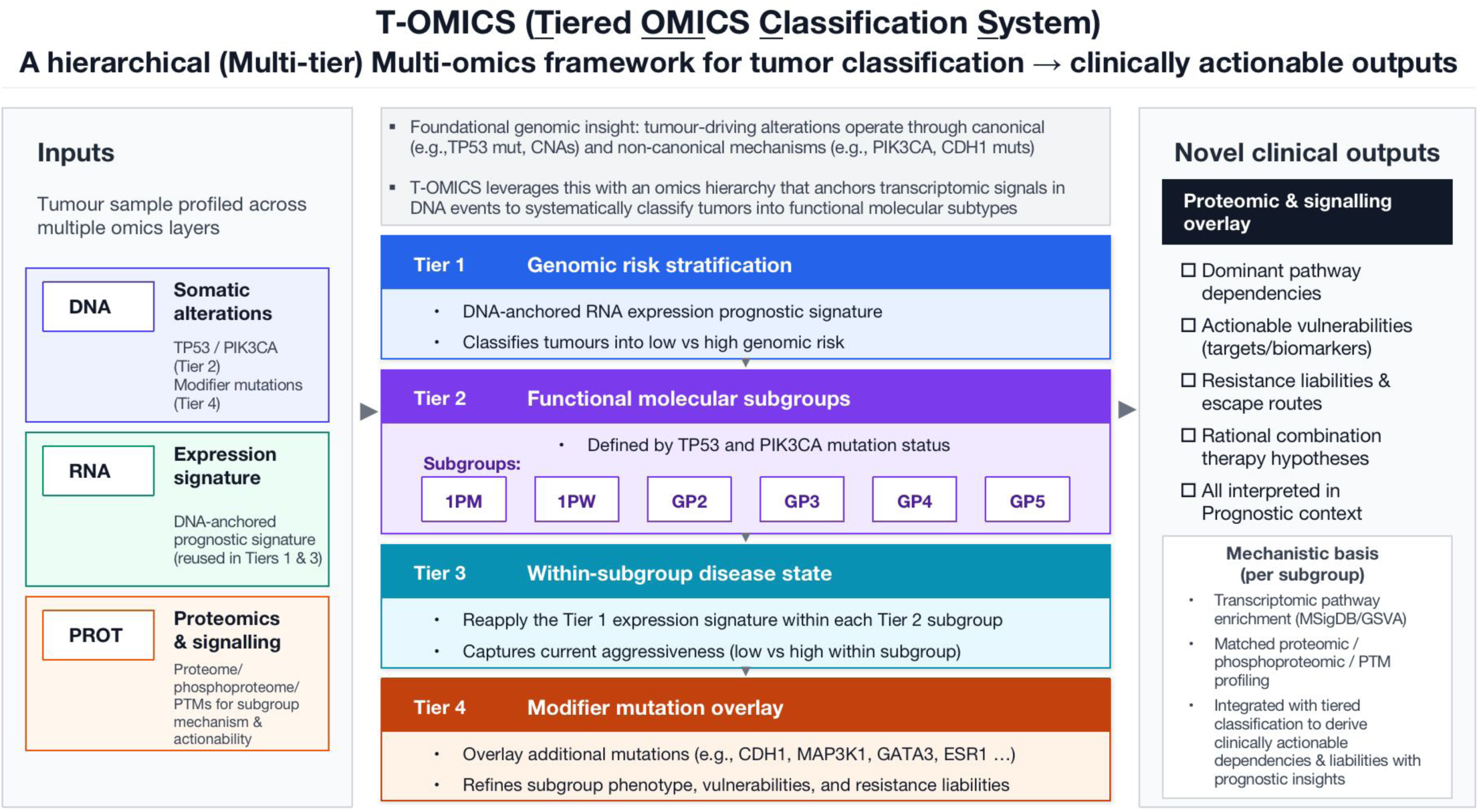
Overview of the T-OMICS tumour classification framework and clinically interpretable outputs. T-OMICS (Tiered OMICS classification System) is a hierarchical multi-omics framework for ER+/HER2− breast cancer that translates tumour genomic complexity into a tiered, clinically interpretable tumour biology profile. For prospective stratification, T-OMICS requires only matched DNA (somatic alterations) and RNA (gene expression) data. Proteomic/phosphoproteomic and signalling measurements are used during framework development to support subgroup mechanism and actionability. They are not required for subgroup assignment in new samples once a reference atlas is established. The framework builds on the genomic discovery that tumour-driving alterations converge on two dominant programmes—canonical proliferative/replicative versus non-canonical, comparatively non-proliferative mechanisms—captured by complementary signatures and operationalised as tiers. Tier 1 applies a DNA-anchored RNA prognostic expression signature to stratify tumours into low- and high-genomic-risk groups. Tier 2 assigns functional molecular subgroups defined by TP53 and PIK3CA mutation status (subgroups 1PM, 1PW, GP2–GP5). Tier 3 applies the Tier 1 expression signature within each Tier 2 subgroup to define a within-subgroup activity/aggressiveness state. Tier 4 overlays modifier mutations (e.g., CDH1, MAP3K1, GATA3, ESR1) to refine phenotype, vulnerabilities, and resistance liabilities without redefining core identity. Proteomic and signalling overlays have been used to link tiered states to pathway activity, supporting clinically oriented outputs, including dominant pathway dependencies, actionable vulnerabilities (targets/biomarkers), resistance liabilities/escape routes, rational combination-therapy hypotheses, and prognostic outlook, reported alongside Tier 1 risk and Tier 3 within-subgroup state.

Although hierarchical, T-OMICS can be delivered as a single integrated report summarising proliferative risk (Tier 1), programme identity (Tier 2), activity/adaptation state (Tier 3), and modifier mutations (Tier 4), enabling clinically interpretable statements about dominant dependencies, expected behaviour, resistance risks, and rational therapeutic strategies anchored to subgroup-associated outcomes. This structure mirrors how breast cancer is already managed—core biology, current clinical behaviour, and actionable features—while grounding each element in tumour-state interpretation rather than an alteration list. This integrated biological–prognostic mapping makes T-OMICS inherently actionable and provides clinicians with a coherent framework for contextualising molecular findings within anticipated clinical trajectories, including in cases where mutation-first approaches offer limited guidance.

PIK3CA exemplifies why programme-aware interpretation is needed. Although PIK3CA is common in ER+/HER2− disease and predictive of response to PI3Kα inhibition in endocrine-resistant advanced settings [10], its prognostic associations have been inconsistent across cohorts and analytic contexts [13,39]. T-OMICS resolves this ambiguity by showing that PIK3CA effects are programme- and state-dependent. In the predominant low-genomic-score state, PIK3CA mutations align with Hippo/AMPK-restrained, epithelial-differentiated, stress-policed biology associated with favourable outcomes and relative resistance to disease acceleration, even with nodal involvement. They also show longer time to metastatic presentation, supporting risk-appropriate management in settings where sequencing is routine, and treatment intensity decisions are frequent [6,13,16]. In contrast, within high-genomic-score disease, PIK3CA marks distinctly adverse biology with dependencies not predicted by proliferation output alone, and these effects differ by TP53 background—emphasising that “PIK3CA-mutant” is not a single biological entity. Mechanistically, TP53-wild-type/PIK3CA-mutant tumours (GP3) exhibit AKT-active but mTORC1-restrained, autophagy-buffered, cytoskeleton/trafficking-driven biology, whereas TP53-mutant/PIK3CA-mutant tumours (GP5) show interferon-wired, checkpoint-primed, redox-compromised states with distinct immune and DDR dependencies and preserved mTOR/translation-node signalling. This framework reconciles conflicting literature by placing PIK3CA within its governing programme architecture.

Importantly, favourable PIK3CA biology in low-score disease is not unconditional. When Tier 4 modifiers are present—most notably a CDH1 co-mutation in the low-score context—the outlook associated with PIK3CA is reversed and becomes significantly worse, demonstrating how modifier events can re-route tumour behaviour within an otherwise favourable programme-state context. Together, these findings support programme- and modifier-aware interpretation of one of the most clinically used biomarkers in ER+/HER2− disease, with direct implications for risk calibration and combination selection in locally advanced and metastatic settings where sequencing is routine.

Beyond PIK3CA, the six Tier 2 subgroups define distinct tumour “failure modes,” reflecting different dominant survival and adaptation systems that sit atop a shared proliferative/replicative backbone. Across subgroups, programme-specific control architectures—such as proteostasis/translation addiction, RTK–ER coupling, innate sensing and inflammatory survival wiring, cytoskeletal/trafficking-driven plasticity, or stress-governed quiescence—provide mechanistic hypotheses for rational therapeutic layering and explain why uniform escalation strategies can underperform in ER+/HER2− disease, where endocrine resistance and adaptive survival programmes drive progression [41,42]. This also clarifies why inferring “pathway activation” from lesions can mislead: a PI3K-pathway alteration does not necessarily indicate that PI3K is the dominant growth engine, but may reflect buffering, lineage maintenance, or immune-interacting survival in a given state.

Tier 3 further shows that aggressiveness evolves along a gradient within each programme. Activity progression was associated not only with increased cell-cycle/replicative markers but also with coordinated, programme-concordant shifts in adaptive circuitry, including metabolic provisioning, inflammatory stress wiring, immune-evasion features, and plasticity-associated signalling in high-score programmes, and with relaxation of checkpoint brakes and partial engagement of replication output in low-score programmes, without wholesale loss of luminal identity. These observations are consistent with broader models in which progression and therapy adaptation reflect dynamic state transitions and stress-response remodelling rather than purely proliferative acceleration [43,44]. Clinically, Tier 3 provides an interpretable within-programme measure of tumour activity that can support rational escalation or restraint within an identity class.

Tier 4 addresses a practical interpretability issue: many frequent canonical alterations converge on proliferative/replicative output already captured by Tier 1, limiting incremental prognostic value downstream. In contrast, recurrent non-canonical alterations such as CDH1, MAP3K1, GATA3, ESR1, and PTEN can refine prognosis and elucidate mechanisms in a context-specific manner, as they modify signalling and cell-state behaviour not reducible to proliferation alone; their clinical impact can be difficult to resolve in unstratified cohorts [45,46]. By treating such events as modifiers that preserve core identity while tuning pathway outputs, vulnerabilities, and resistance liabilities, T-OMICS captures their non-redundant contributions within the programme/state context.

In metastatic disease, a clinically usable framework must accommodate both truncal stability and evolutionary change. Across paired specimens spanning primary–metastatic, metastatic–metastatic, and concurrent primary sampling, Tier 2 programme identity was predominantly conserved, consistent with a largely truncal, patient-specific biology that often persists across anatomical sites and sampling contexts—supporting T-OMICS as a clinically robust stratifier that remains stable despite biopsy-site variation. Tier 2 discordance in a minority of patients (∼25%) indicates greater inter-lesional divergence, consistent with branched evolution and/or contextual selection across metastatic niches and treatment history. Tier 3 state transitions were more frequent, while Tier 4 modifiers were usually absent or stable but occasionally acquired (∼20%), consistent with therapy- and microenvironment-driven adaptation [42,43]. The prominence of CDH1 across both stable and acquired Tier 4 patterns highlights adhesion-axis disruption as a common modifier, whereas PTEN enrichment among stable cases and ESR1 enrichment among acquired/variable cases are consistent with PI3K-pathway perturbation and endocrine-resistance–linked evolution, respectively [25,47]. Collectively, these data support a pragmatic sampling strategy: a representative biopsy can provide a reliable baseline programme context for many patients, with additional sampling reserved for standard clinical scenarios—progression, mixed response, or a new metastatic site—to identify state shifts and emergent modifiers that may refine treatment selection.

This study is observational and requires prospective validation, particularly for predictive utility. Some modifier alterations occur at low frequency, limiting power for outcome analyses; however, the modular design of T-OMICS supports incremental expansion as larger datasets accrue. Future extensions could incorporate quantitative modules for the immune microenvironment, stromal/ECM remodelling, and metastatic competence, particularly given the partial separability of proliferative aggressiveness and metastatic behaviour suggested by our state analyses [42,44]. Prospective studies in early-stage cohorts with long-term follow-up will be important to test whether T-OMICS can refine escalation/de-escalation beyond existing assays and to evaluate predictive performance for targeted combinations earlier in the disease course.

In summary, we propose a programme-aware model for interpreting breast cancer genomes, in which alterations are understood as contributors to tumour-level functional states rather than isolated biomarkers. By integrating proliferative/replicative output, programme identity, activity state, and modifier layers within a prognostic context, T-OMICS translates sequencing results into mechanistically grounded, clinically interpretable guidance. This approach reframes ER+/HER2− disease from a catalogue of lesions into a structured landscape of tumour states, scalable to routine workflows and applicable across early-stage and metastatic settings, providing a foundation for mechanism-based precision oncology.

## Methods

We performed an integrative multi-omics analysis of breast cancer cohorts spanning genomic, transcriptomic, and proteomic profiles. By integrating genomic alterations with transcriptomic expression changes and examining concordant proteomic expression patterns, we explored the molecular landscape of breast cancer across multiple biological layers. The comprehensive computational and statistical methodologies used in this study are detailed in the following sections.

### Integrative Multi-Omics Framework and Analytical Pipeline

The study incorporated multiple breast cancer cohorts, which were analysed either independently or using integrative approaches, as appropriate to the specific analytical objectives. A comprehensive summary of all datasets, including cohort names, data sources, and the total number of samples in each, is provided in **Table S1**. Among these, two major datasets were extensively utilised: the METABRIC (microarray-based) and the TCGA-BRCA (RNA sequencing-based) datasets. The A custom R-based analytical pipeline was developed to systematically evaluate transcriptomic changes associated with recurrent somatic mutations in breast cancer (performs differential expression analysis across the entire transcriptome). Mutation data in Mutation Annotation Format (MAF) were first parsed to extract gene-level mutation status across all samples. For each gene, samples were binarised as *mutated* (1) or *wild type* (0), and this information was compiled into a metadata matrix representing the mutation landscape across the cohort. This matrix served as the foundation for automated generation of contrast designs within the *limma* framework (R/Bioconductor). For each comparison, the system computes log_2_ fold changes, *p*-values, and Benjamini–Hochberg–adjusted *p*-values, enabling a gene-wise estimation of transcriptional impact associated with specific mutations. This approach was applied iteratively across all recurrent genes of interest, encompassing 173 genes previously reported in the METABRIC study [3] (**Table S2**). All downstream analyses and data visualisations were performed using the log_2_ fold change values derived from each recurrent gene’s *mutation versus wild-type* comparison.

A similar analytical framework was used to assess the impact of copy number alterations (CNAs) on gene expression. Gene-level copy number data were retrieved from cBioPortal, where values typically range from −2 to +2. In the analysis, copy number states of −2 and −1 were categorised as deletions, 0 as neutral, and +1 and +2 as amplifications. Copy number data were then integrated with matched gene expression profiles from the same samples, and differential expression analyses were performed using an in-house pipeline adapted for CNA-specific comparisons. For each cancer gene of interest, the corresponding downstream transcriptomic alterations were systematically evaluated across distinct CNA states, including: (i) *Copy Number Altered versus Neutral* [−2, −1, +1, +2 vs 0], (ii) *Amplified versus Neutral* [+1, +2 vs 0], and (iii) *Deleted versus Neutral* [−2, −1 vs 0].

Copy number alteration (CNA) analyses were initially conducted for the 173 genes reported in the METABRIC study [3] and subsequently extended to include the broader cancer-associated gene list presented in **Table S2**. The table lists the analysed genes alongside curated annotations highlighting their functional and clinical relevance in breast cancer. Within this analytical framework, genes with an adjusted *p*-value < 0.05 were considered significantly differentially expressed.

### Gene expression data processing and differential expression analysis

For transcriptome-level analyses, gene expression data were obtained from multiple platforms: log₂ intensity values from the Illumina HT-12 v3 microarray (METABRIC cohort), RNA-Seq V2 RSEM–normalised data from TCGA-BRCA (RNA-Seq), and Agilent microarray expression profiles from TCGA-BRCA (Agilent platform). RSEM values from TCGA-BRCA were log₂(expression+1) transformed to stabilise the mean–variance relationship, thereby ensuring compatibility across datasets. Since the METABRIC and Agilent datasets were already provided as log₂ and log₂ ratio values, respectively, they were used without further transformation.

Differential expression analyses were performed using the in-house analytical pipeline described above. For each gene, the corresponding genomic alteration status (mutated or wild-type; or copy number altered vs neutral) was extracted and encoded as a binary vector, which served as the contrast variable for linear modelling. Within this framework, linear models were fitted independently across the transcriptome (∼18,000 genes) using the *limma* (Linear Models for Microarray Data) package [48], enabling robust estimation of gene-wise expression differences between altered and control groups. The pipeline generates contrast matrices dynamically for each recurrent gene, fits models in bulk mode, and computes log₂ fold-change values, raw p-values, and Benjamini–Hochberg–adjusted p-values for all genes. Empirical Bayes moderation was applied to stabilise variance estimates across genes, improving statistical power and reproducibility. This enables high-throughput, systematic profiling of transcriptional responses to genomic alterations, providing a unified, reproducible framework for mutation- and CNA-driven differential expression analysis across multiple breast cancer cohorts.

### Preprocessing and statistical analysis of proteomic data

Two proteomic datasets utilised in this study include (1) the TCGA Pan-Cancer Atlas (n=104; protein expression quantified by mass spectrometry, comprising 29 PIK3CA mutant and 55 TP53 mutant tumours). (2) Proteomic landscape of breast cancer, CPTAC 2020 (n=122; protein abundance ratios quantified by mass spectrometry, comprising 40 PIK3CA mutants and 50 TP53 mutants). Protein-level expression data were downloaded from cBioPortal [49] and incorporated into the analysis. As the analysis aimed to capture genes with coherent transcriptomic and proteomic expression trends across comparisons, the *limma* framework with empirical Bayes moderation was used to compute moderated *t-statistics* and *adjusted p-values*. An *FDR* threshold of 0.05 was used to define significance.

To evaluate concordance between transcriptomic and proteomic alterations, gene–protein correlation analyses were performed for both proteomic datasets. Genes present in both the mRNA expression and protein abundance matrices were identified and retained for downstream analysis. Log_2_ fold-change (log_2_ FC) values were computed separately for TP53-Mutant vs TP53 wild-type (TP53-WT) and PIK3CA-Mutant vs PIK3CA wild-type (PIK3CA-WT) comparisons, using matched samples for transcriptomic and proteomic data. The resulting log_2_ FC values were then correlated for each gene–protein pair across the CPTAC Breast Cancer and TCGA Pan-Cancer Atlas datasets. The distribution of correlation coefficients was visualised as histograms (**Figure S12**), providing a quantitative overview of the concordance between transcript and protein expression. Furthermore, scatter plots of log_2_FC at the mRNA level against log_2_FC at the protein level were generated for both TP53-Mutant vs TP53-WT and PIK3CA-Mutant vs PIK3CA-WT comparisons.

### Gene Set Categorisation and definition of Group A & Group B genes

An extensive comparative analysis was performed on the DEGs identified in the TP53-Mutant vs TP53-WT and PIK3CA-Mutant vs PIK3CA-WT comparisons. Based on statistical significance and expression directionality, the DEGs were stratified into three categories: *Category 1*: Common genes significantly altered in both comparisons (FDR < 0.05); *Category 2*: Common genes significantly altered in both comparisons (FDR < 0.05) but with opposing fold-change directions between the two contrasts; and *Category 3*: Common genes ranked within the top 500 based on *FDR < 0.05* across both contrasts. These gene categories were independently derived for each breast cancer cohort to ensure robust, reproducible findings. The *Category 3* differential expression analyses for both TP53 and PIK3CA mutation contrasts are provided in **Table S4**. The *Category 3* gene sets, representing the top 500 overlapping DEGs from TP53-Mutant and PIK3CA-Mutant cases, were visualised using volcano plots generated with the *ggplot2* package in R. These plots clearly illustrated the inverse transcriptional trends observed between the two comparisons, with genes upregulated in *TP53*-mutant tumours showing corresponding downregulation in *PIK3CA*-mutant tumours, and vice versa (**Figures 1B, S3A,** and **S3B**).

Using a systematic analytical framework, Group A and Group B alterations were identified that exhibit transcriptomic profiles concordant with the TP53-Mutant and PIK3CA-Mutant phenotypes, respectively. The concordance of these alterations with the corresponding mutation-associated transcriptional changes was further validated across multiple analytical levels. For each BRCA cohort, recurrent gene alterations were included only if they were mutated in at least 0.1% of samples or exhibited copy number alterations in a minimum of 50 samples in at least two of the three transcriptomic datasets analysed in this study. Of the 173 key genes analysed, no somatic mutations in STMN1, KLRG1, and GH1 were detected in the TCGA-BRCA cohort, and no copy number alteration in GPR124 was observed in the METABRIC dataset; therefore, these genes were excluded from downstream analyses.

Cluster stability metrics were computed to quantify the likelihood of each gene aligning with either the TP53-mutant–like (Group A) or PIK3CA-mutant–like (Group B) transcriptional phenotype. The scoring procedure was applied independently across all three categories of gene sets (*Category 1–3*) derived in the study, as well as across the complete fold-change matrix and a subset matrix comprising 60% of genes randomly sampled over 500 iterative runs. For each iteration, hierarchical clustering with k = 2 was performed to generate a dendrogram separating the expression space into two primary clusters based on fold-change correlations, using *cutree* from the *dendextend v1.19.1* package [50]. Consistently, *TP53* and *PIK3CA* were observed on distinct branches, reflecting their characteristic inverse transcriptional trends. Genes that co-clustered with *TP53-Mutation* were assigned to Group A, whereas those co-clustering with *PIK3CA-Mutation* were designated as Group B. Representative dendrograms for each of the contrasts analysed in this study (Mutation vs WT; CNA vs Neutral) are provided in **Figures S6-S10**. Each dendrogram is integrated with an annotation heatmap that captures the recurrent gene’s alteration status, its cumulative frequency across samples, and the corresponding distribution of genomic alterations within ER, PR, and HER2 subtypes.

This procedure was repeated across three transcriptomic datasets—METABRIC, TCGA-BRCA RNA-Seq, and TCGA-BRCA Agilent microarray—under five analytical criteria, and evaluated using multiple genomic alteration contrasts based on fold-change matrices, including Mutation vs Wild-type, CNA vs Neutral, Amplification vs Neutral, and Deletion vs Neutral. A cumulative cluster stability score was computed for each contrast and each gene across all analytical conditions. Genes with a stability score greater than 0.9 were classified as Group A or Group B candidates. For copy number alterations (CNAs), the corresponding TP53 or PIK3CA mutation data were incorporated to guide classification—cluster branches containing TP53 mutations were assigned to Group A, whereas those containing PIK3CA mutations were designated as Group B. The classification of the 173 genes analysed in this study into Group A or Group B across the mutation and CNA analyses is provided in **Supplementary Table 5**.

### Visualisation and functional enrichment of Group A and Group B-associated transcriptomic phenotypes

To visualise transcriptomic directionality across alteration classes, we generated combined heatmaps of log_2_ fold-change values for Category 3 differentially expressed genes (DEGs) across four contrasts for each recurrent alteration: mutation vs wild-type, CNA vs copy-neutral, amplification vs copy-neutral, and deletion vs copy-neutral. Heatmaps were produced in R using *ComplexHeatmap (v2.22.0)* [51], with columns split to denote Group A (TP53-like) and Group B (PIK3CA-like) alterations.

Differential expression analyses were performed using the in-house analytical pipeline described above. For each contrast (mutation vs wild-type; CNA vs copy-neutral), genes with p<0.05 were partitioned into upregulated (log_2_FC>0) and downregulated (log_2_FC<0) sets and analysed separately by gene set enrichment analysis using *clusterProfiler (v4.14.6)* and MSigDB Hallmark gene sets. Up- and down-enrichment results were annotated accordingly and summarised in a single dot plot.

### Mutual Exclusivity and Co-occurrence Analysis

Mutual exclusivity and co-occurrence analyses for the Group A and Group B gene sets were performed using the cBioPortal platform (https://www.cbioportal.org/). For the association analysis, combined breast cancer datasets, as detailed in **Supplementary Table 1**, were used. Somatic mutations and copy number alterations (CNAs) for each gene were analysed separately. The significance of co-occurring or mutually exclusive alterations between gene pairs was assessed using Fisher’s exact test implemented in cBioPortal, with results reported as p-values and odds ratios (**Tables S6** and **S7**). Gene pairs with a *q-value < 0.05* were considered statistically significant. Heatmaps of mutual exclusivity and co-occurrence among gene pairs were generated using the *corrplot (v0.95)* package [52] in R.

### CNA–Transcriptome Coupling Analysis

We systematically evaluated the relationship between cancer-associated CNAs and TP53 mutation-associated transcriptomic programs in multiple tumour cohorts. Analyses were performed separately within the METABRIC and TCGA breast cancer cohorts, both in the full set of tumour samples and in the subset restricted to ER-positive, HER2-negative tumours to ensure clinical homogeneity. In addition, the same framework was applied to TCGA uterine endometrial carcinoma and prostate adenocarcinoma cohorts to assess whether the observed patterns generalised across cancer types. For each DEG set (as shown in the figure), log_2_-normalised expression values were averaged per sample and stratified by CNA states of cancer-associated genes. CNA levels were defined as −2 (deep deletion), −1 (shallow deletion), 0 (diploid/neutral), +1 (gain), and +2 (high-level amplification). To avoid spurious estimates from sparsely populated categories, extreme states were merged with adjacent groups when necessary. In METABRIC breast cancer, −2 and −1 were combined, and +2 was included only if supported by ≥10 samples (≥0.5–0.7% of the cohort). In TCGA breast cancer, −2 and +2 were retained only if represented by ≥5 samples (≥0.5–0.7%). In TCGA uterine endometrial carcinoma, −2 was merged with −1, and +2 was retained if supported by ≥6 samples (≥1% of the cohort). In TCGA prostate adenocarcinoma, both −2 and +2 were merged with their adjacent categories (−1 and +1, respectively) because extreme categories were variably sparse across genes; merging emphasised consistent copy loss versus gain effects and reduced noise.

For visualisation, boxplots were used to depict the distribution of mean DEG expression across CNA levels, with loess-smoothed curves overlaid to highlight global trends. This design enabled us to assess whether copy number losses and gains of cancer-associated genes, regardless of whether the CNA-targeted gene was an oncogene or a tumour suppressor, converged on expression patterns resembling those of TP53-mutant tumours. Statistical differences between CNA levels were evaluated using Student’s t-tests, with significance reported at standard thresholds *(*p < 0.05; **p < 0.01; ***p < 0.001; ****p < 0.0001*). The *ggplot2 (v4.0.0)* package [53] was used to generate the boxplots.

### Identification of Prognostic Gene Signature

To derive a robust prognostic signature, shared DEGs between TP53- and PIK3CA-mutant tumours were identified independently in the METABRIC and TCGA breast cancer cohorts. Genes consistently differentially expressed across both datasets were retained as candidate genes for survival modelling. The prognostic relevance of individual genes was assessed using univariate Cox proportional hazards regression with overall survival as the endpoint in the METABRIC ER+/HER2– cohort. Genes significantly associated with survival (p < 0.05) were entered into multivariable Cox proportional hazards modelling and refined using stepwise variable selection to derive candidate prognostic signatures. Stepwise selection was applied to the univariate-filtered gene set to reduce redundancy and derive a stable set of independently prognostic transcripts. Candidate stepwise models were compared using prespecified performance metrics, including the concordance index (C-index) and time-dependent ROC/AUC. As a sensitivity analysis, penalised Cox regression with LASSO regularisation was also performed on the same candidate gene set to evaluate the robustness of gene selection and prognostic performance. For the final stepwise-derived, parsimonious signature, a patient-level risk score was calculated as a weighted sum of gene expression values using the corresponding model coefficients (risk score = Σβ_i ×_ Expr_i_). Patients were stratified into low- and high-risk groups using a clinically anchored cut-off based on Kaplan–Meier estimates to identify the subgroup with an estimated overall survival probability> 90% at 10 years post-diagnosis. The independent prognostic value of the resulting risk groups (high vs low) was then assessed using multivariable Cox regression adjusting for established clinicopathologic and treatment covariates (age, tumour stage, histological grade, lymph node status, tumour cellularity, inferred menopausal status, and receipt of chemotherapy, endocrine therapy, and radiotherapy), with hazard ratios, 95% confidence intervals, and Wald p-values reported. The prognostic performance of the signature was subsequently externally validated in independent cohorts, using identical risk-score calculation and cut-off definitions.

### Survival Analysis

Overall survival (OS) and progression-free survival (PFS) were evaluated using Kaplan–Meier survival analysis across the identified subgroups in the METABRIC and TCGA cohorts, respectively. Survival curves were generated with time (years) plotted on the x-axis and survival probability on the y-axis. For each subgroup, the total number of patients (n), the number of observed events, and the estimated survival probability at the specified time point (accounting for censoring) were reported in the plot legend. Differences in survival distributions were assessed using the log-rank test.

To quantify survival differences, multivariable Cox proportional hazards regression models were fitted. The regression output included regression coefficients, hazard ratios (HRs) with 95% confidence intervals (CIs), and corresponding p-values. An HR > 1 indicated an increased risk of death, whereas an HR < 1 indicated a reduced risk relative to the reference group. Survival differences between groups were further evaluated using pairwise Wald tests within Cox proportional hazards models across all possible group combinations; only appropriate or statistically significant comparisons were reported. All survival analyses were performed using *survival (v3.8.3)* and *survminer* (v0.5.1) packages of R.

### Pathway activity estimation and statistical analysis (GSVA and pathway heatmaps)

#### (a) Cohorts and expression preprocessing

Pathway activity was quantified using two complementary approaches, gene set variation analysis (GSVA) and single-sample GSEA (ssGSEA), implemented in the *GSVA R package (v2.0.7)* [54]. The combined expression reference dataset, comprising z-score-standardised METABRIC (Illumina microarray) and TCGA (RNA-seq TPM/RSEM) cohorts restricted to ER-positive/HER2-negative samples, was used as input. An independent validation cohort was assembled from four publicly available RNA-seq datasets retrieved from cBioPortal (**Table S1**): CPTAC breast cancer, MBC Project (provisional 2022 release), MBC Project (archival 2017 release), and the SMC (Korean) breast cancer cohort. Expression units were harmonised to log_2_(TPM+1): CPTAC log_2_(FPKM) values were back-transformed, converted from FPKM to TPM, and then log_2_(TPM+1) transformed; MBC 2022 (TPM) and SMC (TPM) were log_2_(TPM+1) transformed; MBC 2017 (FPKM) was converted to TPM and log_2_-transformed, and samples overlapping with the 2022 MBC release were removed to avoid duplication. Within each dataset, expression matrices were collapsed to gene symbols (averaging values corresponding to the same gene symbol), merged using the intersection of common genes, and batch-corrected across cohorts using *ComBat (sva v3.54.0).* Batch correction was assessed by PCA before and after adjustment. For biological comparability, analyses were restricted to ER-positive/HER2-negative tumours. ER and HER2 status were obtained from clinical annotations when available; for SMC, where HER2 status was not reported, ER-positive samples were retained. Following concordant results across reference and validation analyses, cohorts were additionally combined into a complete reference dataset for downstream stratification by T-OMICS categories; an analogous workflow was applied to metastatic subsets within the MBC cohort.

#### (b) Gene sets and GSVA scoring

Gene sets were obtained from MSigDB v2025.1, including Hallmark, KEGG (legacy and Medicus), Reactome, WikiPathways, C4 (cancer gene neighbourhoods), C6 (oncogenic signatures), C7 (immunologic signatures), and C8 (cell type signatures). Collections were merged into a single gene-set library, and GSVA and ssGSEA enrichment scores were computed with parallelisation enabled.

#### (c) Differential pathway activity and visualisation

Differential pathway activity between tumour subtypes was assessed using *limma* with empirical Bayes moderation. For subtype-specific effects, each subtype was compared against the equal-weighted mean of all other tumour subtypes (one-versus-rest contrasts with equal group weighting to reduce sensitivity to unbalanced subtype sizes), excluding normal samples from the tumour-only design matrix. Additional contrasts compared each subtype to normal breast tissue and compared normals versus all tumours (normal vs equal-weighted mean of subtypes). For each contrast, moderated t-statistics, log fold-changes, and Benjamini–Hochberg FDR-adjusted p-values were computed. Subtype-level pathway profiles were summarised as the mean GSVA or ssGSEA score across samples within each subtype. Heatmaps were generated with *pheatmap v1.0.13* [55], displaying both unscaled and row-scaled enrichment scores, with significance overlaid as star annotations *(****FDR<0.0001, ***FDR<0.001, **FDR<0.01, *FDR<0.05*). Hallmark pathways were prioritised in the main figures due to interpretability and robustness.

This multi-layered analytical strategy enabled identification of subtype-specific pathway activity, tumour–normal differences, and consistent signalling patterns across cohorts, and included cross-validation using GSVA and ssGSEA to ensure that results were not dependent on a single enrichment method or contrast specification.

### Pathway cluster identification and cluster-level enrichment analysis

With >10,000 pathways analysed, direct interpretation of individual enrichment results was impractical. To identify biologically coherent pathway groups, we constructed a pathway–pathway similarity matrix based on gene-set overlap. For all MSigDB gene sets, pairwise similarity was computed using the Jaccard index (|A∩B| / |A∪B|), which quantifies the proportion of shared genes between two pathways while normalising for pathway size. The resulting similarity matrix was subjected to average-linkage hierarchical clustering, and clusters were defined using the *dynamic tree cut algorithm* (*dynamicTreeCut v1.63.1*; deepSplit = 3, minClusterSize = 10) [56], enabling adaptive branch pruning based on the dendrogram structure rather than a fixed height threshold. This approach yielded discrete clusters of pathways with substantial gene overlap, representing related biological processes.

For each pathway cluster, the most frequently occurring genes were extracted to define representative markers. These marker lists were used to assign functional labels to clusters via a structured, reproducible annotation workflow. Each cluster was assigned a stable identifier (e.g., “C2: Cell Cycle”) to ensure consistent referencing across analyses.

Cluster-level enrichment analysis was then performed to prioritise biologically relevant pathway modules within each breast cancer subgroup. Differentially activated pathways identified by equal-weighted one-versus-rest contrasts (*FDR < 0.05*; top 1,000 up- or downregulated pathways per subgroup) were tested for overrepresentation within pathway clusters. The background list comprised the union of all clustered pathways. For each set of differentially activated up- or down-regulated pathways, the enriched pathway clusters were identified by computing the number of pathways shared between the query and the pathway clusters; the number of pathways in the query but not in the pathway cluster; the number of pathways in the pathway cluster but not in the query; and the number of pathways in the background set that are neither in the query nor in the cluster. Enrichment was assessed using Fisher’s exact test on 2×2 contingency tables (query pathways vs cluster membership), and p-values were adjusted with the Benjamini–Hochberg procedure to control the false discovery rate. Significantly enriched clusters (*FDR < 0.05*) were visualised as dot plots, providing a structured overview of pathway-level differences among Tier 2 molecular subgroups.

### RPPA protein annotation and functional grouping

To support interpretation of RPPA findings, proteins were curated and annotated using pathway mappings from Konaté et al. [57] and complementary literature-guided annotation that was supported by structured, OpenAI-assisted prompts. Annotated proteins were then assigned to pathway groups and classified by their expected direction of effect (positive vs negative regulators) in breast cancer (**Table S13**).

### PAM50 Subtyping

The PAM50 subtype was derived for all samples using the *genefu (v2.34.0)* R package. It implements the original PAM50 gene expression–based centroid classifier and has been widely validated for breast cancer molecular subtyping [58]. For METABRIC, normalised log_2_-transformed microarray expression data were used, while TCGA RNA-seq data were transformed to log_2_(TPM+1) scale to ensure comparability. Gene identifiers were harmonised to official HGNC symbols, and in cases where multiple probes mapped to the same gene, the mean expression value was retained. The analysis utilised the PAM50 reference centroid model to assign each tumour sample to one of five intrinsic subtypes—Luminal A, Luminal B, HER2-enriched, Basal-like, or Normal-like—based on similarity to predefined gene expression patterns. Subtype probabilities and correlations to PAM50 centroids were extracted to assess classification confidence. Harmonised PAM50 calls were then used for cross-cohort comparisons with T-OMICS strata.

### Association of T-OMICS subgroups with clinicopathological characteristics

Associations between T-OMICS classifications (Tier 1 risk groups and Tier 2 molecular subgroups) and clinicopathological variables, including PAM50 intrinsic subtype, tumour size, tumour stage, Nottingham Prognostic Index (NPI), and histological grade, were evaluated. For each clinicopathological variable, contingency tables were constructed to summarise the frequency of categories across T-OMICS groups. Global associations were tested using Pearson’s χ² test; when expected cell counts were <5 in any Tier 2 subgroup, Fisher’s exact test was used. Analyses were implemented using *gtsummary (v2.4.0)*. To identify subgroup–category combinations driving significant associations, standardised Pearson residuals were computed and visualised as heatmaps (*pheatmap*), with |residual|>3 interpreted as meaningful deviations from expectation (positive = enrichment; negative = depletion). For statistically significant associations, the effect size was quantified using Cramér’s V (*rcompanion v2.5.0*), with higher V values indicating stronger association. Results were additionally summarised with percentage stacked bar plots showing the distribution of clinical categories within each Tier 2 subgroup.

### Time-to-metastasis analysis

Time to metastasis was defined as the interval from primary diagnosis to the first documented metastatic event. As all patients in the analysed cohort developed metastatic disease, all observations were coded as events (event = 1), and no censoring was applied. Differences in time to metastasis between subgroups were assessed using Kaplan–Meier survival analysis implemented in the *survival* (v3.8.3) and *survminer* (v0.5.1) R packages [59]. Log-rank tests were used to assess global differences across subgroups, and the corresponding p-values were shown on Kaplan–Meier plots. Risk tables showing the number of patients at risk at each time point were included. To quantify subgroup effects, Cox proportional hazards models were fitted using the *coxph() function* (*survival v3.8.3* R package) [60], with the subgroup included as a categorical covariate. The statistical significance of subgroup effects was assessed using Wald tests derived from the Cox model, and pairwise comparisons between the reference subgroup (1PM) and other subgroups were reported as Wald p-values.

To visualise sample-level variability, individual time-to-metastasis values were shown as jittered scatter points in *ggplot2*, with subgroup-level means and corresponding confidence intervals overlaid as point estimates with horizontal error bars. Wald test p-values from the Cox model were annotated directly on the plots for clarity.

### Sankey and Alluvial plot visualisation of T-OMICS transitions

Sample-level transitions across tumour tissue location, patient identity, and T-OMICS Tier 2, Tier 3, and Tier 4 subgroup assignments were visualised using the *sankeyNetwork()* function from the *sankeyD3 R package (v0.3.2)* [61]. Each tumour specimen was treated as an individual observation and mapped along ordered axes representing anatomical site and successive T-OMICS tiers. Patient identity was retained as a linking variable to track multiple samples from the same individual across the T-OMICS framework tiers. Divergent flows within a patient indicate potential evolutionary branching during metastatic progression, reflecting tumour adaptation across anatomical sites or disease stages. Conversely, persistent flows across multiple samples from the same patient suggest a conserved biological backbone, representing stable, core molecular programs maintained during disease evolution.

## Data Availability

All data produced in the present work are contained in the manuscript

## Acknowledgments

This work was supported by private angel investor funding. We thank the investigators and patients who contributed data to TCGA, METABRIC, and other public breast cancer cohorts that enabled this study. We thank the developers and maintainers of community bioinformatics resources and software packages that supported this work. We thank Pratik Kothari for support with fundraising and project management, and Nitesh Shriwash for contributions to script and code development.

## Author Contributions

A.G. conceived and designed the study and led the work. A.G. performed data processing and all computational analyses and made the core genomic discovery, revealing a robust functional organisation of genomic alterations that decodes tumour genomic complexity. A.G. developed the functional alteration grouping framework and derived the DNA-anchored RNA signature and complementary DNA mutation tiers. A.G. implemented the T-OMICS tiered classification system and performed proteomic/phosphoproteomic and signalling overlay analyses to support mechanistic interpretation and clinical translation of T-OMICS subgroups. A.G. provided clinical interpretation. M.M. contributed to data visualisation and the development of code and scripts. A.G. and M.M. generated figures and visualisations. A.G. wrote the manuscript with input from all authors. A.G. supervised the project and acquired funding. A.G. is the corresponding author. All authors reviewed and approved the final manuscript.

