## Supplementary Appendix A for "A unifying functional dichotomy organises breast cancer molecular landscape, resolves PIK3CA ambiguity, and supports tiered tumour classification"

A deeper interrogation of Tier 2 subgroup wiring and candidate dependencies is provided below (including defining features, pathway annotations, marker lists, and concordant RNA–protein features):

### Subgroup 1PM (PIK3CA-mutant, low-genomic score): a quasi-quiescent, translation-quiet, epithelial-differentiated, FAO/autophagy-adapted with mesenchymal–innate-evasion wiring

1PM represents a circadian clock-aligned, fatty acid oxidation (FAO)-powered, Hippo-AMPK-restrained "stress-policed, slow-cycling" low-risk ER+/HER2− state that channels MAPK and PI3K signals into stress resilience rather than growth, sustains itself via autophagy and translational control (catabolic, growth-restraining programmes rather than biosynthetic expansion), and exhibits low replication stress with tempered homologous recombination (HR). Overall, 1PM is characterised by high checkpoint- and stress-response-governed signalling and restrained growth (Stress-activated, checkpoint-restrained), despite oncogenic PIK3CA mutations, yielding low proliferative drive and unexpectedly stable outcomes even with nodal involvement (**Figure S33A)**.

#### *1.1 Cell-cycle and Translation Control*

1PM exhibits deep G1 restraint and suppressed cap-dependent translation: very low Cyclin B1/E1/D3, FOXM1, AURORA-A, CDC25C, CDK1-pY15, p90RSK-pT573, p4EBP1 and S6-pS240/244, with elevated cell cycle/proliferation negative regulators PDCD4, PTEN, IGFBP3, and p27 (via dephosphorylation of p27 at T198). ERK/JNK/p38 (total and phosphorylated) and CREB pS133 are high, but their anabolic output is muted. BAK, p16INK4a and PUMA are high, indicating cell death (apoptosis) and senescence brakes (**Figures 7A, 7E, S31,** and **S32**).

#### *1.2 Stress/Energy, Autophagy and Hippo/YAP*

Proteomically, 1PM shows elevated phosphorylation of YAP at S127 (suggesting cytoplasmic sequestration via Hippo-mediated inhibition – corroborated by transcriptomic enrichment of multiple Hippo/YAP-restraining modules) and of AMPK at T172, together with increased p38 and JNK activation (total p38, total JNK2, p38-pT180/Y182, JNK-pT183/Y185), highlighting an active stress-response network. EEF2K, ATG4B, and ATG5 are high, with p62 low, indicating operational autophagy and translational elongation control that favour survival in low-anabolism states. Transcriptomics show selective enrichment of TGFb and senescence/autophagy programmes (WP_CANONICAL_AND_NONCANONICAL_TGFb_SIGNALING, WP_SENESCENCE_AND_AUTOPHAGY_IN_CANCER). Together, this pattern is consistent with active Hippo signalling, energy-stress sensing, and engagement of senescence/autophagy programmes that collectively oppose cell-cycle entry.

#### *1.3 DNA Repair Tone*

RAD51 and PARP1/PARG are low, whereas MERIT40 pS29, RAD50, and RPA32 are high, implying tempered homologous recombination and PARylation flux, with intact BRCA1-A/MRN signalling for DNA-damage sensing rather than high-fidelity repair throughput (a "vigilant but low-throughput" configuration). DNA replication stress sensors are also toned down (ATR pS428, CHK1 pS345, CHK2 pT68, RPA32 pS4/S8, MSH2, MSH6 all low), further supporting a low-replication, low-replication stress state.

#### *1.4 Metabolism*

Pathway-level transcriptomic analyses show enrichment of mitochondrial/peroxisomal fatty-acid β-oxidation modules (including unsaturated/medium-chain fatty acids) in 1PM. Hallmark and curated gene sets related to fatty acid β-oxidation (HALLMARK_FATTY_ACID_METABOLISM, KEGG_FATTY_ACID_METABOLISM, WP_FATTY_ACID_BETAOXIDATION, REACTOME_MITOCHONDRIAL_FATTY_ACID_BETA_OXIDATION) and PPARα/AMPK-coupled metabolic control (WP_PPARALPHA_PATHWAY) are selectively upregulated. SLC1A5 and ASNS are low, consistent with reduced glutamine/asparagine anabolism and greater reliance on lipid oxidation and autophagy-mediated nutrient recycling.

#### *1.5 ECM/Adhesion & Innate-Immune Evasion (1PM-specific)*

Vinculin, PDGFRB, AXL, Connexin-43, PKCα, PKC pan βII pS660, and PI3K-p85 are elevated, indicating strong cell–cell and cell–matrix adhesion, wound-healing/repair, and an organised cytoskeleton, consistent with an epithelial, non-migratory phenotype. Transcriptomics show enrichment of the CXCR4–Gα12/13–RHOA axis, likely reflecting microenvironmental tuning and local tissue residency (a "retention" bias), consistent with low risk/quiescence rather than active invasion (EMT-leaning adhesion). Furthermore, the 1PM subgroup shows a parainflamed, vaccine/infection-like immune transcriptome, marked by recurrent innate danger-sensing and tissue-conditioning modules (multiple TLR/PRR stimulation contexts; IL-1/inflammasome-linked programmes such as NALP12/NLRP1/CLEC7A-inflammasome and IL-1 processing; vaccine/viral-response–like interferon contexts; and injury/repair signatures such as burn/wound-healing). However, alongside this, immune-evasion markers B7-H3/B7-H4 and myeloid checkpoint cues (SIRP-α axis, MIF) are also elevated, suggesting myeloid-biased immune evasion. This indicates 1PM as a "danger-experienced", chronic stress-conditioned epithelial state with an immune-vigilant but functionally tolerised microenvironment, rather than an immune-desert or a fully 'hot' phenotype.

#### *1.6 Lost biology (vs typical ER+ disease)*

In 1PM mTORC1-driven translation/protein-synthesis drive (p-S6, p-4EBP1), de novo lipogenesis (SCD1), glycolysis/PPP (G6PD) and some amino-acid auxotrophies (ASNS) are curtailed; they show relative suppression of canonical growth-factor and nutrient-sensing gene-set modules, including insulin/RTK–RAS–ERK, mTOR and vesicular/secretory trafficking signatures (e.g. KEGG_INSULIN_SIGNALING_PATHWAY, WP_TARGET_OF_RAPAMYCIN_SIGNALING, COPII/COPI) and downregulation of SOS-mediated signalling, NTRK2→FRS2/3→RAS, and FGF/FGFR–RAS–ERK modules, indicating that classic growth-factor pathways are functionally muted at the output level, reducing the drive into translation/cell-cycle entry; glutamine import (SLC1A5) is low along with very low AMPK-a2-pS345 phosphorylation – a marker of AMPK activity suppression under nutrient (amino-acid) abundance. Altogether, these features mark a Hippo-on, AMPK-on, stress-MAPK-on, fuel-efficient, stress-resilient, replication-cautious state that reads PI3K and environmental/immune stress and pushes the cell towards quiescence, repair, and controlled autophagy, not aggressive cycling. This supports a model in which 1PM tumours are not simply "weakly PI3K-driven", but instead maintain an actively enforced, metabolically adapted quiescent state.

### Subgroup 1PW (PIK3CA-wild-type, low-genomic score): Low-proliferative neuroendocrine-like/GPCR–RTK signalling with mTORC2–ERK buffering

The 1PW subgroup (PIK3CA–wild-type) represents a signalling-rich, growth/translation-primed yet brake-engaged neuroendocrine-like low-risk state, distinct from both 1PM and the high-risk groups (**Figure S33B**).

#### *Neuronal/neuroendocrine and hormone-linked identity*

Transcriptomically, 1PW is characterised by neuronal and synaptic programmes absent in 1PM, including *REACTOME_NEURONAL_SYSTEM*, *REACTOME_TRANSMISSION_ACROSS_CHEMICAL_SYNAPSES*, *REACTOME_NEUROTRANSMITTER_RECEPTORS_AND_POSTSYNAPTIC_SIGNAL_TRANSMISSION*, *KEGG_OLFACTORY_TRANSDUCTION*, and *KEGG_LONG_TERM_POTENTIATION*. Enrichment of *WP_GABA_RECEPTOR_SIGNALING*, *WP_BIOGENIC_AMINE_SYNTHESIS*, and sensory modules (e.g. *REACTOME_SENSORY_PROCESSING_OF_SOUND*, *REACTOME_SENSORY_PERCEPTION_OF_TASTE*) suggests neuroendocrine/neuromodulatory wiring with prominent GPCR and ion-channel signalling. Consistent with this, the proteome shows higher levels of ERα, PR, glucocorticoid receptor, cKIT, IGF1Rβ, c-MET, ACVRL1, and YAP, indicating that 1PW remain responsive to growth factors and steroid hormones, but without translating this into high proliferative output.

#### *RTK–MAPK–YAP signalling with restrained mTORC1 and cell cycle*

Proteomically, 1PW shows a robust RTK–MAPK axis, as evidenced by high levels of RTKs and adaptor proteins (including IGFRβ, IRS2, EGFR pY1173/pY1068, c-MET, cKIT, FGF basic, SHP2, SHP2 pY542, INPP4B) and a strong receptor→MAPK tier **(**high MEK1 pS217/S221, p44/42 MAPK/ERK1/2, JNK pT183/Y185, p90RSK total and pT359/S363, YB1 pS102, EphA2 pS897, PKCα pS657, and STAT3 pY705). High P70S6K pT389 and eIF4E pS209 point **to** mTORC1→S6K and cap-dependent translation initiation as a core effector programme in 1PW. They also show high levels of structural/hippo-related proteins (CAVEOLIN-1, YAP, ZEB1).

However, this signalling appears to function as a "buffering/maintenance" circuit rather than a growth engine, as canonical anabolic and proliferative outputs are strongly muted. P70S6K1, S6 (total), S6 pS240/S244, 4EBP1 pS65/pT70, EIF4G, EEF2, and Akt (total) are all significantly lower, and the cell-cycle and replication machinery is consistently depressed (low Cyclin D3, Cyclin E1/E2, Cyclin B1, FOXM1, PLK1, MYT1, RRM2, PCNA, CDC25C, CDK1 pY15), while the negative-feedback and checkpoint-brake proteins are prominent: high DUSP4 and DUSP6 (ERK/JNK phosphatases), p27**,** WEE1 pS642**,** PEA15, and high 14-3-3 ε/β – consistent with maintenance of G1/S and G2/M brakes despite active upstream RTK–MAPK input (**Figures 7A, 7E, S31,** and **S32**). DNA damage and replication stress sensors are also toned down (ATR pS428, CHK1 pS345, CHK2 pT68, RPA32 pS4/S8, MSH2, MSH6 all low), further supporting a low-replication, low-replication-stress state. High total RICTOR with high RICTOR pT1135 is consistent with an mTOR network under feedback regulation (pT1135 is commonly discussed as an inhibitory/feedback phosphorylation on RICTOR). This supports a "feedback-wired" mTORC1↔mTORC2 system—a hallmark of a *growth-competent* rather than *stress-restrained,* low-risk state. Overall, 1PW appears to be a quiescent, **"**signal-receptive but brake-engaged**"** configuration: abundant RTK–ERK–JNK–STAT3 activity and YAP expression, but with cell-cycle and mTORC1-driven translation actively restrained.

#### *Survival, apoptosis, and stress buffering*

Survival wiring further supports a "buffered, brake-engaged" state. High BCL2, DJ-1 (PARK7), MIF, PEA15, BCL2A1, Bid, 14-3-3 proteins, GCLC, SOD1, LKB1, and AceCS1, together with low Bax and MCL1, indicate greater anti-apoptotic and redox support. Caspase-8 and cleaved caspase-8 are elevated, yet caspase-3 and cleaved caspase-7 are low, suggesting that extrinsic death pathways are primed but restrained at the mitochondrial level. Furthermore, stress-adapted chaperone networks and membrane trafficking are evident (e.g. Calnexin, RAB11, Collagen VI, Annexin VII), consistent with controlled secretory and receptor turnover. Metabolically, GCLC/SOD1/DJ-1/LKB1 support redox control and energy-stress sensing, while reductions in PHGDH, IDO, MCT4, GAPDH, and mTORC1 outputs indicate that 1PW is neither strongly Warburg-like nor strongly FAO-addicted, but rather runs on a moderate, antioxidant-protected metabolic programme.

High P-cadherin**,** JAB1, and SMAD4 align with an epithelial-but-adaptable luminal identity that retains organised regulatory architecture (vs high-risk dedifferentiation), while lower p38 MAPK suggests that 1PW is relatively less stress-policed than 1PM, aligning with a more permissive growth state.

#### *Microenvironment and EMT/ECM tone*

Relative suppression of FIBRONECTIN, AXL, SIRPα, MYOSIN IIA, FN14, TFRC, SYK, JAK2, and MCT4 indicates that 1PW is less mesenchymal, less ECM-remodelled, and less myeloid-interactive than more aggressive subgroups. Despite YAP and ZEB1 expression, the absence of strong fibronectin/AXL and low mechanotransductive "scaffold" suggests that YAP is present but not driving a full EMT/invasive programme in this context.

Overall, 1PW appears to be a "signalling-on, proliferation-off" configuration, endocrine-driven with RTK–MAPK plasticity, internal brakes, cell-cycle control, and mTORC1-driven translation actively restrained. Therapies should exploit these liabilities (endocrine±SHP2/MEK/ERK adaptors) upon endocrine resistance.

### Cross low-genomic-risk subgroup comparison (1PM vs 1PW)

Although 1PM and 1PW both fall within low-genomic-risk, largely luminal ER+/HER2− disease, they represent distinct low-risk attractor states with different control systems, biology, and clinical behaviour. The key divergence is that 1PM is a stress-policed, catabolic/quiescent state governed by FAO–AMPK–Hippo circuitry and ECM/immune conditioning, whereas 1PW is a signal-receptive, endocrine/RTK-tuned state with neuroendocrine/GPCR wiring, in which upstream growth-factor signalling is active yet held in check by feedback and cell-cycle brakes (**Figure S33C**).

#### *Deeper suppression of proliferation and translation in 1PM*

Many proliferative and anabolic regulators that are already low in 1PW are further suppressed in 1PM, including Cyclin E1, Cyclin D3, FOXM1, AURORA-A, CDC25C, CDK1 pY15, S6 pS240/S244, p90RSK pT573, and translation/metabolic nodes (GCN5L2, HSP60, ASNS), alongside lower MSH2 and reduced p27 pT198. This deep quiescence is accompanied by strong restraint circuitry in 1PM—high YAP pS127, high AMPK pT172, prominent p38/stress signalling, and enrichment of p53-mediated checkpoint programmes at the transcriptomic level (e.g.,

REACTOME_TP53_REGULATES_TRANSCRIPTION_OF_CASPASE_ACTIVATORS_AND_CASPASES, REACTOME_TP53_REGULATES_TRANSCRIPTION_OF_CELL_DEATH_GENES). In contrast, 1PW is characterised by prominent RTK–MEK–ERK/JNK signalling with feedback and brake engagement (DUSP4/DUSP6, WEE1 pS642, p27) and a comparatively more "growth/translation-primed" configuration, albeit with low proliferative output (**Figures 7A, 7E, S31,** and **S32**).

##### *ECM/AXL/autophagy and infection/vaccine-like imprint in 1PM*

Compared with 1PW, FIBRONECTIN, AXL, SIRPα, and ATG3 are significantly higher in 1PM. This points to a more ECM-anchored, AXL-positive, immune-modulating and autophagy-dependent state, even though net proliferation remains low. Together with infection/vaccine-like gene signatures in 1PM (multiple PBMC/vaccine and pathogen-stimulation modules), this suggests a stronger "danger-experienced" or chronically conditioned epithelium with durable "danger memory", greater reliance on autophagy and ECM/AXL-coupled survival under stress, and a microenvironment in which immune editing and myeloid crosstalk (via SIRPα and ECM) may be more developed than in 1PW.

##### *Clinical interpretation and therapeutic implications*

Both low-risk subgroups share low chromosomal instability and replication stress, with sustained disease control overall, but they reach this state through distinct biology. 1PM reflects high stress-sensing and enforcement (Hippo-on, AMPK-on, stress-MAPKs/inflammatory nodes-on), which channels PI3K/MAPK signals into checkpointing, quiescence/dormancy, and catabolism rather than biosynthetic expansion—consistent with clinical observations that lymph-node involvement has reduced prognostic impact and that time-to-metastatic presentation is longer in 1PM than in 1PW. By contrast, 1PW represents an archetypal luminal, endocrine-dependent "quiet but less policed" state: upstream signalling and survival buffering are more engaged, but there is less evidence of the pronounced dormancy-like stress-governance architecture seen in 1PM, which may permit more expansion-competent disseminated disease once spread occurs.

### Shared Core Biology in High-Genomic-Score Breast Cancer Subgroups
### Shared Core Biology in High-Risk Breast Cancer Subgroups

Across all tumours, high-genomic score status is uniformly associated with a core programme of cell-cycle activation and genome maintenance, together with loss of normal developmental and tissue-specific programmes (e.g. nephrogenesis, vascular/muscle and cardiac conduction pathways). All high-risk groups show coordinated enrichment of *HALLMARK_E2F_TARGETS*, *HALLMARK_G2M_CHECKPOINT*, and multiple REACTOME/KEGG/WP modules for G1–S and G2–M transitions, DNA replication, ATR/ATM signalling, p53 and checkpoint control, Fanconi/HR/BER/MMR repair, and telomere maintenance, indicating replication under chronic DNA damage surveillance as a shared feature of all high-risk groups (**Figure S35A**). In parallel, shared downregulated pathways highlight loss of normal differentiation, circadian control, and stromal/mesenchymal programmes, as well as loss of broad physiological growth-factor and detoxification programmes, including normal RTK–ligand diversity, integrin/ECM homeostasis, and Phase I/II biotransformation pathways (e.g. cytochrome P450, conjugation and antioxidant metabolism modules), consistent with dedifferentiation, metabolic rewiring, and immune suppression across high-risk subgroups (**Figures 7A, 7E, S31,** and **S32**).

Subgroup 3, among other high genomic-score groups, however, exhibited a relatively distinct (relatively silent) transcriptomic and RPPA profile (the odd one out) despite high clinical risk. The other high genomic score groups (all but one) were distinguished by additional, higher-order layers of oncogenic and metabolic rewiring that were not significantly altered in Subgroup 3. First, they showed stronger and broader induction of RTK and mTORC1 signalling modules (including multiple EGFR/HER2, FGFR, MET, FLT3 and fusion-RTK pathways), together with anabolic and nucleotide biosynthesis, and mevalonate/cholesterol biosynthesis. Second, these groups exhibited extensive ubiquitin–proteasome and unfolded protein response activation, consistent with a state of proteostasis addiction superimposed on a high MYC/mTOR-driven biosynthetic load. Third, they showed more profound suppression of developmental signalling axes (Hedgehog, WNT/β-catenin, NOTCH, TGF-β, FOXO and circadian/cardiac conduction pathways) beyond the core developmental/tissue programmes already lost in all high-risk tumours, consistent with a deeply dedifferentiated, lineage-poor, proliferation-focused phenotype (**Figure S35B**).

Functionally, this positions the all-but-one high-risk tumours not only as cell-cycle and DDR-dependent (a feature shared with Subgroup 3), but also as addicted to growth-factor signalling, mTOR-driven anabolic metabolism and proteostasis, and liberated from differentiation and temporal control. Consequently, while CDK and ATR/CHK1/WEE1/PARP-based DDR targeting are rational across all high-risk tumours, the all-but-one groups present additional, subgroup-specific vulnerabilities to RTK and mTOR blockade, proteasome and UPR/chaperone targeting, and metabolic interventions (e.g. nucleotide and mevalonate/cholesterol pathway blockade), in which these pathway dependencies are most pronounced. These features define the all-but-one cohorts as "high-risk plus target-rich", built on the shared replication-stress backbone that characterises high-risk disease overall.

Whereas TP53-mutant subgroups GP4 (TP53-mutant/PIK3CA–WT) and GP5 (TP53-mutant/PIK3CA–mutant) together define a canonical kinase-addicted high-risk state, characterised by strong activation of RTK→PI3K–mTORC1/2 signalling, glycolysis and pentose-phosphate metabolism, and NRF2-driven antioxidant programmes, superimposed on an inflamed, interferon-rich tumour microenvironment with active antigen presentation and T-cell circuitry (**Figure S35C**). What distinguishes them from the other high-risk groups is not a different oncogenic driver but the *depth* of pathway dismantling. They show coordinated suppression of primary cilia and Hedgehog/Notch/BMP/TGF-β developmental signals, broad loss of peroxisomal and mitochondrial fatty-acid oxidation and ketone-body use, and attenuation of LKB1–AMPK energy sensing and endocrine/nuclear receptor pathways (including oestradiol/progesterone and insulin/IGF – HALLMARK_ESTROGEN_RESPONSE_EARLY/LATE, WP_ESTROGEN_RECEPTOR_PATHWAY, nuclear-initiated ER/PR signalling KEGG modules) and REACTOME_SIGNALING_BY_IGF1R, REACTOME_INSULIN_RECEPTOR_SIGNALLING_CASCADE, REACTOME_IRS_MEDIATED_SIGNALLING, WP_INSULIN_SIGNALING_IN_ADIPOCYTES, WP_LEPTIN_AND_ADIPONECTIN). As a result, GP4/GP5 appear as highly dedifferentiated, hormone-independent, "kinase-addicted" tumours that are metabolically locked into glucose/PPP and glutamine use, with limited capacity to switch to oxidative lipid fuels and a heavy reliance on NRF2–GSH buffering for redox control. These features suggest shared vulnerabilities across all-but-one high-risk groups to RTK/PI3K–mTOR and cell-cycle/replication-stress targeting, but highlight GP4/GP5 as ideal candidates for combination strategies that pair RTK/PI3K–mTOR blockade with metabolic stress (glycolysis/glutaminase inhibition or mitochondrial stress), redox perturbation (GSH/ferroptosis-based approaches), and immunotherapy within their highly engaged immune microenvironment.

### Gained (Unique) Biology in Subgroup 2: defines a luminal, RTK–ER–metabolism–driven high-risk programme with buffered AKT output

Among high-risk tumours, GP2 (TP53-PIK3CA double wild-type) is characterised by coordinated activation of luminal RTK and nuclear receptor signalling**,** translation–proteostasis**,** and mitochondrial–metabolic programmes, despite relatively limited canonical AKT phosphorylation. At the signalling level, GP2 shows selective enrichment of oestrogen receptor pathways (*REACTOME_ESR_MEDIATED_SIGNALING*, *WP_ESTROGEN_SIGNALING*, *REACTOME_SIGNALING_BY_NUCLEAR_RECEPTORS*), consistent with high ERα and GATA3 protein expression and a luminal epithelial phenotype (high E-cadherin, Claudin-7, DDR1). This luminal axis is integrated with growth factor signalling via insulin/IGF1R–PI3K pathways (*REACTOME_SIGNALING_BY_INSULIN_RECEPTOR*, *REACTOME_SHC_RELATED_EVENTS_TRIGGERED_BY_IGF1R*, *KEGG_MEDICUS_REFERENCE_IGF2_IGF1R_PI3K_SIGNALING_PATHWAY*) and HER2-centric ERBB networks (*REACTOME_SHC1_EVENTS_IN_ERBB2_SIGNALING*). Consistent with elevated HER2 (total and pY1248), IGFRβ and IRS2, these data support a model of GP2 as a HER2/IGF-addicted, ER-positive high-risk state in which proximal RTKs are strongly engaged, but downstream AKT phosphorylation (pS473/pT308) and canonical NF-κB/STAT3 activation remain comparatively restrained (**Figures 7A, 7E, S31,** and **S32**).

At the effector level, GP2 is characterised by a prominent mTORC1–translation–proteostasis axis**.** Pathways related to mTOR signalling (*REACTOME_MTOR_SIGNALLING*, *WP_TARGET_OF_RAPAMYCIN_SIGNALING*, *KEGG_MEDICUS_REFERENCE_GATOR1_MTORC1_SIGNALING_PATHWAY*), translation and ribosome control (*REACTOME_TRANSLATION*, *WP_TRANSLATION_FACTORS*), and ubiquitin-mediated proteolysis/ER quality control (*KEGG_UBIQUITIN_MEDIATED_PROTEOLYSIS*, *REACTOME_E3_UBIQUITIN_LIGASES_UBIQUITINATE_TARGET_PROTEINS*, *WP_CELLULAR_PROTEOSTASIS*, *REACTOME_ER_QUALITY_CONTROL_COMPARTMENT_ERQC*, *REACTOME_N_GLYCAN_TRIMMING_IN_THE_ER_AND_CALNEXIN_CALRETICULIN_CYCLE*) are selectively up-regulated, consistent with high levels of RAPTOR, P70S6K1, total S6, 4EBP1, EIF4E, CALNEXIN, P62, HSP27, HSP60 and GRP75. Notably, S6 and 4EBP1 phosphorylation are less hyperactivated than in TP53-mutant groups, indicating that GP2 is translation-addicted yet maintains a partially "buffered" mTORC1 output. Complementary enrichment of secretory and vesicular trafficking pathways (*REACTOME_COPII_MEDIATED_VESICLE_TRANSPORT*, *REACTOME_TRANSPORT_TO_THE_GOLGI_AND_SUBSEQUENT_MODIFICATION*, *KEGG_GLYCOSYLPHOSPHATIDYLINOSITOL_GPI_ANCHOR_BIOSYNTHESIS*, *WP_GLYCOSYLPHOSPHATIDYL_INOSITOL_ANCHOR_PATHWAY*) is consistent with a high RTK and adhesion-receptor load at the plasma membrane.

Metabolically, GP2 adopts a lipogenic and OXPHOS-competent state, with up-regulation of fatty-acid and lipid biosynthesis pathways (*REACTOME_FATTY_ACYL_COA_BIOSYNTHESIS*,

*KEGG_BIOSYNTHESIS_OF_UNSATURATED_FATTY_ACIDS*, *WP_LIPID_METABOLISM_PATHWAY*) and mitochondrial oxidative phosphorylation and respiratory chain assembly (*WP_OXIDATIVE_PHOSPHORYLATION*, *WP_ELECTRON_TRANSPORT_CHAIN_OXPHOS_SYSTEM_IN_MITOCHONDRIA*, *REACTOME_COMPLEX_III_ASSEMBLY*, *REACTOME_COMPLEX_IV_ASSEMBLY*, *WP_MITOCHONDRIAL_COMPLEX_IV_ASSEMBLY*). This is mirrored at the protein level by high FASN, ACC1, SLC1A5, Mitofusin-1/2, Cox-IV, TUFM, DNA POLG, GRP75 and MS12, indicating coordinated engagement of anabolic lipid metabolism, amino-acid handling and mitochondrial biogenesis. GP2 has relatively lower total AMPK and AMPK pT172 levels, suggesting a disabled AMPK "circuit breaker**"** arm that would typically throttle these pathways in response to metabolic stress. Integrated with an expanded DNA damage and replication-stress programme (*REACTOME_DNA_DAMAGE_RECOGNITION_IN_GG_NER*, *REACTOME_FORMATION_OF_TC_NER_PRE_INCISION_COMPLEX*, *KEGG_MEDICUS_REFERENCE_DOUBLE_STRAND_BREAK_SIGNALING*, *REACTOME_SENSING_OF_DNA_DOUBLE_STRAND_BREAKS*) and cell-cycle regulators (Cyclins D1/D3/E2, Cdc6, Aurora-A, PLK1, CDK1 pT14/pY15; MSH2/MSH6, PMS2, RRM2), GP2 relies on robust NER/MMR and checkpoint function (high MSH2, MSH6, PMS2, XRCC1, DNA Pol-γ) in the setting of high proliferation and replication stress, with relatively lower ATM indicating an ATR–CHK1–Wee1-skewed dependency (Wee1, CHK1 pS345 elevated**)**.

Conversely, GP2 shows relative suppression of stromal/ECM and cytokine–inflammatory signalling, with down-regulation of ECM and integrin/adhesion programmes **(e.g.,** REACTOME_EXTRACELLULAR_MATRIX_ORGANIZATION**,** REACTOME_COLLAGEN_FORMATION/DEGRADATION**,** REACTOME_INTEGRIN_CELL_SURFACE_INTERACTIONS**,** REACTOME_CELL_EXTRACELLULAR_MATRIX_INTERACTIONS, elastic-fibre/collagen-fibril assembly modules), alongside broad attenuation of cytokine/JAK–STAT modules (e.g**.,** KEGG_CYTOKINE_CYTOKINE_RECEPTOR_INTERACTION**,** KEGG_JAK_STAT_SIGNALING_PATHWAY, interleukin modules including IL-6/IL-4/IL-10/IL-17/IL-23/IL-35 signalling sets, complement/coagulation-related programmes). Importantly, many of the uniquely downregulated gene sets are annotated to adaptive immune lineages and activation states—spanning CD4 activation/differentiation programmes, Treg/Tfh-associated modules, and B-cell state transitions (naïve/GC/plasma/memory)—supporting few effective cytotoxic T, Tfh, Treg, and B-cell programmes and therefore functionally low adaptive immunity in GP2. Consistent with this, the proteomic profile indicates that tumour-intrinsic inflammatory and immune signalling is relatively muted, with lower NFκB p65 pS536**,** STAT3 pY705**,** STAT5α**,** Lyn**,** SIRPα and ARAF pS299**,** accompanied by reduced mechanotransduction/focal-adhesion tone (lower YAP**,** paxillin**,** vinculin) and lower immune co-inhibitory markers (e.g., B7-H4). Together, these data support GP2 as a de-stromatised, immune-quiet, ECM-low, cytokine/JAK–STAT–quiet and adaptively "cold" subgroup with reduced stromal engagement and limited productive immune activation.

Together with maintained E-cadherin/Claudin-7 and high ER/DDR1, this supports a model in which GP2 tumours remain epithelial and cohesion-preserving, and are driven predominantly by luminal RTK–ER signalling, translation–proteostasis, mitochondrial metabolism and ATR-biased DNA damage responses, rather than by EMT, inflammatory or microenvironmental programmes (**Figure S34A**). This integrated wiring diagram directly motivates combined HER2/IGF–ER blockade, mTORC1/translation and proteostasis targeting, OXPHOS/lipogenesis inhibition, and ATR–CHK1–Wee1 axis interference as rational therapeutic strategies for this GP2 high-risk subgroup. Moreover, relatively low AMPK tone alongside high mTOR/translation + lipogenesis/OXPHOS suggests potential vulnerability to AMPK re-activation as a metabolic brake in GP2.

### Gained (Unique) Biology in Subgroup 3 (the Odd-One-Out High-Risk Subgroup): PIK3CA-mutant, AKT-active yet mTORC1-restrained, Rho-driven, autophagy-dependent proliferators with lower EGFR/IGF1R dependence

The PIK3CA-mutant odd-one-out subgroup retained the core proliferative programme observed across high-risk tumours (e.g. HALLMARK_E2F_TARGETS, HALLMARK_G2M_CHECKPOINT, REACTOME_CELL_CYCLE_MITOTIC) but was distinguished by a stress-integrating PI3K–metabolism–autophagy axis. Pathway analysis highlighted enrichment of WP_AMPACTIVATED_PROTEIN_KINASE_SIGNALING, REACTOME_PI_METABOLISM, and REACTOME_PHOSPHOLIPID_METABOLISM, with elevated AMPKα, total and phospho-AKT, TUBERIN pT1462, PTEN and PCNA, low phospho-4EBP1, and increased ATG3 protein levels. Together, these data support a model in which PIK3CA-driven growth is buffered by AMPK-centred metabolic control, PTEN, and pro-survival autophagy, rather than maximal RTK-mTORC1 activation/proteostasis addiction (low EGFR pY1068 and IGF1R_pY1135Y1136) (**Figures 7A, 7E, S31,** and **S32**).

Concurrently, this subgroup exhibited a primed but not disabled p53 network, characterised by elevated PUMA, PTEN, and TP53-regulatory pathways (REACTOME_PI5P_REGULATES_TP53_ACETYLATION, REACTOME_REGULATION_OF_TP53_ACTIVITY_THROUGH_METHYLATION, REACTOME_DEATH_RECEPTOR_SIGNALING), alongside low JAB1, indicating preserved apoptotic competence under genotoxic or replication stress. In parallel, death receptor and NF-κB–linked pathways (TNFR1/ceramide production, TRAF6-mediated NF-κB activation, extrinsic apoptosis regulation) were selectively enriched. Similar to other high-risk clusters, canonical replication-stress checkpoints and repair modules (e.g. REACTOME_ACTIVATION_OF_ATR_IN_RESPONSE_TO_REPLICATION_STRESS, KEGG_HOMOLOGOUS_RECOMBINATION, KEGG_NUCLEOTIDE_EXCISION_REPAIR, REACTOME_DISEASES_OF_BASE_EXCISION_REPAIR, REACTOME_FANCONI_ANEMIA_PATHWAY) were prominently engaged, indicating that these tumours rely on ATR–HR, NER/BER/Fanconi alternative repair pathways, and the p53–PTEN–PUMA checkpoint axis for survival. Furthermore, this subgroup enriched mitochondrial biosynthesis (REACTOME_TRANSCRIPTIONAL_ACTIVATION_OF_MITOCHONDRIAL_BIOGENESIS, REACTOME_MITOCHONDRIAL_BIOGENESIS) and nucleotide/folate/amino acid metabolism programmes (WP_PYRIMIDINE_METABOLISM_AND_RELATED_DISEASES, KEGG_ONE_CARBON_POOL_BY_FOLATE, KEGG_SULFUR_METABOLISM, KEGG_LYSINE_DEGRADATION), indicating that the subgroup is highly invested in genome and mitochondrial maintenance and fine-tuned nucleotide/one-carbon metabolism.

Finally, these tumours exhibited a junction-poor, invasive phenotype characterised by low levels of E-cadherin, β-catenin, claudin-7, DDR1, and higher COUP-TFII and β-actin. They also showed unique up-regulation of multiple Rho-family GTPases and trafficking modules: REACTOME_RHO_GTPASE_CYCLE, REACTOME_RHOB_GTPASE_CYCLE, REACTOME_RHOC_GTPASE_CYCLE, REACTOME_VXPX_CARGO_TARGETING_TO_CILIUM, REACTOME_INTRA_GOLGI_TRAFFIC, REACTOME_INTRA_GOLGI_AND_RETROGRADE_GOLGI_TO_ER_TRAFFIC, and KEGG_MEDICUS_REFERENCE_EARLY_ENDOSOMAL_FUSION. This profile suggests a tumour heavily dependent on cytoskeletal dynamics, polarity, and vesicle trafficking, with a focus on cell shape, migration, invasion, and microenvironmental sensing rather than proliferation (relatively low cell-cycle/proliferation markers compared with other high-genomic-score groups, including total Aurora A and Aurora-ABC pT288 pT232 pT198, but higher FAK pY397). This profile is consistent with a non-canonical EMT-like, COUP-TFII–driven invasive state, in which cell–cell junctions and epithelial polarity are dismantled, but invasion is orchestrated primarily through Rho/FAK/cytoskeletal signalling rather than classical WNT/β-catenin or ECM/RTK-driven invasion. The subgroup also shows distinctive epigenetic plasticity, with enrichment of REACTOME_HDMS_DEMETHYLATE_HISTONES, REACTOME_HATS_ACETYLATE_HISTONES, WP_HISTONE_MODIFICATIONS, and REACTOME_REGULATION_OF_ENDOGENOUS_RETROELEMENTS_BY_THE_HUSH_COMPLEX modules, suggesting that these high-risk tumours rely on dynamic epigenetic rewiring to buffer DNA damage and inflammatory stress and to adjust proliferation and differentiation programmes without engaging the strong RTK/mTOR hyperactivation seen in the other high-risk groups. The proteome profile confirms this odd-one-out group as more immunogenic and STING-active, and autophagy-addicted for stress mitigation (with higher STING, ATG3, CD4, and HMHA-1 protein levels).

Overall, Subgroup 3 comprises AKT-active, mTORC1-restrained, ATR-low, PUMA-primed, COUP-TFII/Rho-driven, STING-high, autophagy-dependent proliferators with reduced epithelial adhesion and weak RTK signalling, and pro-survival death-receptor and NF-κB circuits that allow the tumour to tolerate chronic stress and inflammatory cues, resulting in a replication-stressed but survival-adapted high-risk phenotype.

Therapeutically, these data argue against RTK-centric strategies (low EGFR Y1068, reduced IGF1R phosphorylation) and instead prioritise: (i) PI3Kα/AKT inhibition combined with autophagy blockade (targeting the AMPK–autophagy module), (ii) DNA-damaging or replication-stress–inducing regimens that exploit preserved TP53/PUMA and relatively weak ATR/HR engagement, and (iii) agents targeting adhesion and cytoskeletal signalling (e.g. the Rho/ROCK–FAK–SRC axis) or NF-κB circuits, in combination with immune-modulatory approaches to leverage high STING and $CD4^{+}$ infiltration (**Figure S34B**).

Although both 1PM and the odd-one-out subgroup show Rho GTPase enrichment, 1PM predominantly engages RHO–PAK/ROCK and polarity-related cycles, consistent with mechanical homeostasis in a quiescent, FAO-primed epithelium, whereas the odd-one-out subgroup acquires broad Rho GTPase cycling involving RHOB/RHOC/RHOT1, CIT- and PKN-dependent programmes that couple Rho to cytokinesis, invasion, organelle dynamics and inflammatory/death-receptor signalling, thereby converting a metabolically poised low-risk clone into a replication-stressed, invasive high-risk state.

### Gained Biology in Subgroup 4: a TP53-mutant, translationally addicted, replication-stressed, metabolically rewired, antioxidant-buffered, high-risk subgroup

The GP4 subgroup defines a TP53-mutant, high-risk state in which ERK circuitry remains intact but is not quantitatively dominant. GP4 uniquely enriches ERK-related input modules, including KEGG_MEDICUS_REFERENCE_CXCR_GNB_G_ERK_SIGNALING_PATHWAY, KEGG_MEDICUS_REFERENCE_EGF_EGFR_PLCG_ERK_SIGNALING_PATHWAY, and REACTOME_ONCOGENIC_MAPK_SIGNALING. Increased CRAF, GRB7, PLCγ1, and p90RSK phosphorylation indicate intact GPCR/EGFR–ERK signalling despite lower total ERK, p38α, and JNK protein levels. Nonetheless, the biology is primarily driven by mTOR–translation and replication stress — marked by high levels of 4EBP1 (total and pS65/pT70), S6 (total and pS240/244), eIF4G, MNK1, EEF2, and PAICS, alongside strong DNA damage and checkpoint signalling (ATR pS428, CHK1 pS345, CHK2 pT68, RPA32 pS4/S8, CDK1 pT14/pY15, CDC25C, PCNA), FOXM1, PLK1, Cyclin B1/E1/E2, RRM2, and elevated p53 protein in a TP53-mutant context. This is coupled with a metabolic stress response characterised by upregulation of folate/one-carbon and vitamin/cofactor pathways (REACTOME_METABOLISM_OF_FOLATE_AND_PTERINES, WP_FOLATE_METABOLISM, REACTOME_METABOLISM_OF_VITAMINS_AND_COFACTORS), amino acid and TCA cycle metabolism (REACTOME_METABOLISM_OF_AMINO_ACIDS_AND_DERIVATIVES, REACTOME_CITRIC_ACID_CYCLE_TCA_CYCLE, KEGG_PYRUVATE_METABOLISM), and glutathione/ROS handling and ferroptosis (KEGG_GLUTATHIONE_METABOLISM, WP_NRF2_PATHWAY, WP_FERROPTOSIS, WP_TRYPTOPHAN_CATABOLISM_LEADING_TO_NAD_PRODUCTION), consistent with high G6PD, PHGDH, SLC1A5, HEXOKINASE II, MCT4, ENOLASE1, and GAPDH protein levels. Additionally, increased IDO, PD-L1, and pro-death pathways (WP_APOPTOSIS_MODULATION_AND_SIGNALING, REACTOME_PYROPTOSIS), coupled with reduced BCL2, position GP4 as a hyper-proliferative yet fragile, stress-loaded state, distinct from other high-risk clusters with a potent immunosuppressive tumour environment that helps GP4 evade the immune system (**Figures 7A, 7E, S31,** and **S32**).

Concurrently, GP4 suppresses multiple homeostatic and differentiation circuits, including canonical Notch and TGF-β/Nodal signalling (*KEGG_NOTCH_SIGNALING_PATHWAY*, *WP_NOTCH_SIGNALING_WP61*, *REACTOME_SIGNALING_BY_TGFB_FAMILY_MEMBERS*, *REACTOME_SIGNALING_BY_NODAL*, *REACTOME_NOTCH1_INTRACELLULAR_DOMAIN_REGULATES_TRANSCRIPTION*, *REACTOME_RUNX3_REGULATES_NOTCH_SIGNALING*), endocrine/nuclear receptor modules for lipid and cofactor homeostasis (*REACTOME_NR1H2_NR1H3_REGULATE_GENE_EXPRESSION_TO_LIMIT_CHOLESTEROL_UPTAKE*, *REACTOME_DEFECTS_IN_VITAMIN_AND_COFACTOR_METABOLISM*, *REACTOME_BIOTIN_TRANSPORT_AND_METABOLISM*, *WP_BIOTIN_METABOLISM_INCLUDING_IMDS*), and branched-chain amino-acid/fatty-acid disposal (*REACTOME_BRANCHED_CHAIN_AMINO_ACID_CATABOLISM*, *REACTOME_DISEASES_OF_BRANCHED_CHAIN_AMINO_ACID_CATABOLISM*, *REACTOME_CARNITINE_SHUTTLE*). Down-regulation of

*REACTOME_SIGNALING_BY_FGFR1/3/4*, *REACTOME_SIGNALING_BY_FLT3_FUSION_PROTEINS* and IGF/ACTH modules (*KEGG_MEDICUS_REFERENCE_IGF_IGFR_PI3K_NFKB_SIGNALING_PATHWAY*, *KEGG_MEDICUS_REFERENCE_ACTH_CORTISOL_SIGNALING_PATHWAY* and variants) supports a shift from regulated growth-factor control to autonomous, stress-tolerant/addicted proliferation. These integrated features nominate clear therapeutic vulnerabilities: (i) replication-stress checkpoint dependence, suggesting ATR, CHK1/CHK2 and WEE1 inhibitors, particularly in combination with PARP inhibition and DNA-damaging therapies; (ii) mTOR–4EBP1–S6–eIF4F dependence, highlighting mTORC1/2 or translation-initiation blockade; (iii) metabolic liabilities in serine–one-carbon, glutamine and REDOX/ferroptosis pathways, suggesting redox intervention that overwhelms buffering (targeting PHGDH, SLC1A5, G6PD, glutathione/GPX4); and (iv) immune vulnerabilities via PD-L1 and IDO, supporting the addition of checkpoint blockade and IDO-modulating agents.

Overall, GP4 is best viewed as a TP53-mutant, replication- and translation-addicted, metabolically stressed, high-risk subtype with preserved ERK input wiring but low bulk MAPK levels, whose key liabilities lie in DDR checkpoints, mTOR-dependent translation, and oxidative/ferroptotic metabolism rather than in classic high-ERK signalling (**Figure S34C)**.

### Gained Biology in Subgroup 5: a TP53-PIK3CA double mutant, interferon-wired, checkpoint-primed adverse outcome subgroup with exploitable redox vulnerability

GP5 defines an "innate-immune–rewired" poor-prognostic state that sits on the shared replication-stress and DDR backbone of all high-risk tumours (and shares glycolytic–PPP fueling with GP4), yet has a distinct immune and metabolic tone from other subgroups, including GP4. Transcriptionally, GP5 is dominated by innate immune sensing and inflammatory signalling modules, with strong activation of nucleic acid sensing and type I interferon pathways (e.g. KEGG_RIG_I_LIKE_RECEPTOR_SIGNALING_PATHWAY**,** KEGG_CYTOSOLIC_DNA_SENSING_PATHWAY**,** DDX58/IFIH1-mediated IFNα/β induction**,** WP_TYPE_I_INTERFERON_INDUCTION_AND_SIGNALING_DURING_SARSCOV2_INFECTION**,** WP_MRNA_VACCINE_ACTIVATION_OF_DENDRITIC_CELL_AND_INDUCTION_OF_IFN1), coupled to robust TLR/NOD/inflammasome–NF-κB and JAK–STAT signalling (e.g. WP_TOLLLIKE_RECEPTOR_SIGNALING_RELATED_TO_MYD88**,** WP_NODLIKE_RECEPTOR_NLR_SIGNALING**,** WP_CANONICAL_NFKB_PATHWAY**,** IL2/IL12/IL23–JAK–STAT modules), in line with high EPPK1 protein levels. In parallel, GP5 is enriched for TCR/BCR and co-stimulatory networks and explicit immune-checkpoint/immunotherapy signatures **(e.g.** WP_T_CELL_RECEPTOR_AND_COSTIMULATORY_SIGNALING**,** WP_B_CELL_RECEPTOR_SIGNALING**,** WP_CANCER_IMMUNOTHERAPY_BY_PD1_BLOCKADE**,** WP_CANCER_IMMUNOTHERAPY_BY_CTLA4_BLOCKADE**,** PD-1–PD-L1–SHP–PI3K signalling), marking it as interferon-primed, checkpoint-engaged, chronically stimulated, Th1/antiviral-like microenvironment consistent with an immune-interacting tumour ecosystem. Unlike more classically RTK/MAPK-addicted subgroups, GP5 shows relatively low MAPK (ERK) activity and WEE1 pS642, reduced IRS2, and high IGFBP2, with preserved mTOR/translation-node signalling (4EBP1 and S6 phosphorylation), DDR and cell-cycle activation (CHK1/CHK2, 4EBP1/S6, PCNA, PLK1, FOXM1, PARP1, etc.), indicating attenuated G2/M braking and dampened classical RTK–ERK–IRS2 signalling in favour of alternative survival routes (autocrine, integrin/RTK-coupled pro-survival PI3K/AKT axis). This dovetails with the many GP5-unique PI3K-linked pathways (BCR/BCAP/CD19–PI3K, PDGF–PI3K, ERBB–PI3K, CXCR4–PI3K, PD-1/PD-L1–SHP–PI3K). This suggests that survival is sustained more by PI3K–JAK–STAT–NF-κB and translation/DDR modules than by a strong MAPK/ERK dependency (**Figures 7A, 7E, S31,** and **S32**).

Therapeutically, this biology points to a different “sweet spot” from that of the other all-but-one high-risk groups. As with all high-risk tumours, the replication-stress/DDR and cell-cycle wiring argue for ATR–CHK1/CHK2–CDK1/2 and PARP inhibition as a rational backbone. However, GP5’s unique enrichment of interferon/RIG-I/MDA5/TLR/TNF–NF-κB and PD-1/CTLA-4 pathways suggests immune-checkpoint blockade (anti–PD-1 ± anti–CTLA4) is particularly attractive, ideally combined with agents that further amplify innate sensing (e.g. STING or TLR3/7 agonists, oncolytic or mRNA-like platforms). At the same time, GP5 (in contrast to a “well-buffered” antioxidant programme in GP4) shows selective loss of detoxification and metabolic flexibility pathways (e.g., REACTOME_GLUTATHIONE_CONJUGATION, WP_GLYCINE_METABOLISM_INCLUDING_IMDS, WP_MITOCHONDRIAL_LONG_CHAIN_FATTY_ACID_BETAOXIDATION; with broader lineage/tissue programmes also reduced), indicating a redox-compromised state that likely survives by leaning harder on glycolysis/PPP compensation rather than robust glutathione conjugation. This supports oxidative overload therapeutic approaches—leveraging impaired glutathione/mitochondrial programmes to sensitise to ROS-inducing radiotherapy/chemotherapy and potentially to ferroptosis- or redox-modulating agents. Concomitant loss of neuronal/secretory and ciliary-like programmes (REACTOME_NEUROTRANSMITTER_RELEASE_CYCLE, REACTOME_GLUTAMATE_NEUROTRANSMITTER_RELEASE_CYCLE, WP_SYNAPTIC_VESICLE_PATHWAY, multiple cilium/autophagosome transport and peroxisomal-lipid modules) reinforces a deeply dedifferentiated, non-physiological phenotype in which lineage and microenvironmental homeostasis are sacrificed in favour of proliferation, innate-immune signalling and survival. In contrast, the ERK-low, WEE1-low context implies that classical MEK/ERK or WEE1-centric monotherapies are less likely to be dominant vulnerabilities in GP5 than in other high-risk groups. Altogether, GP5 emerges as a checkpoint-primed, IFN/TLR-driven, redox-compromised high-risk state, biologically and therapeutically distinct from the more MAPK-biased and metabolically anabolic high-risk subgroups (**Figure S34D)**.
