## Supplementary Figures for "A unifying functional dichotomy organises breast cancer molecular landscape, resolves PIK3CA ambiguity, and supports tiered tumour classification"

A

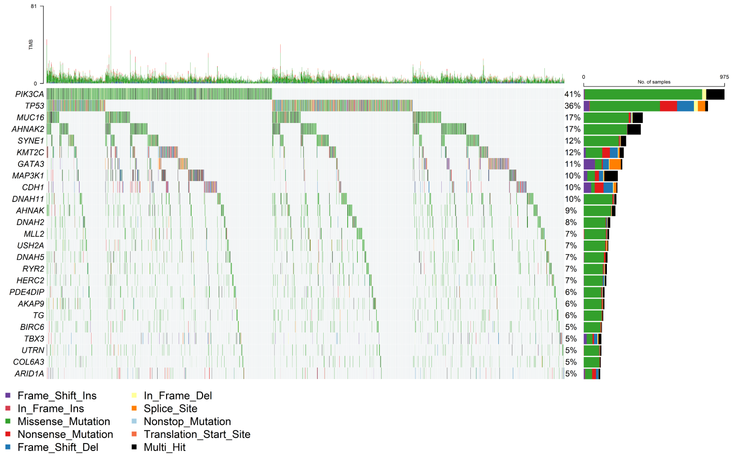


B

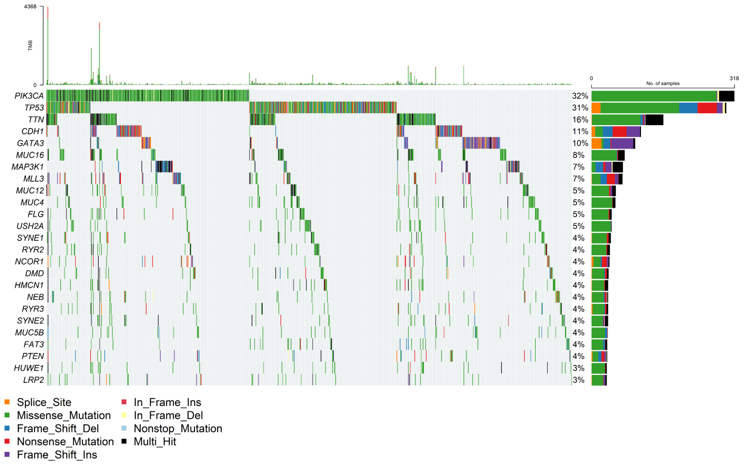


**Supplementary Figure 1 | Landscape of recurrent somatic mutations in breast cancer across METABRIC and TCGA cohorts**. The top 25 most frequently mutated genes in the METABRIC **(A)** and TCGA-BRCA **(B)** cohorts are shown as oncoplots generated using the *maftools* R package. Each column represents a sample, and each row represents a gene. Mutation types are colour-coded, including missense (green), nonsense (red), frameshift insertions (blue), and frameshift deletions (orange), as indicated in the figure legend. The top bar plot displays the tumour mutation burden (TMB) for each sample, while the right-side bar plot indicates the mutation frequency for each gene.

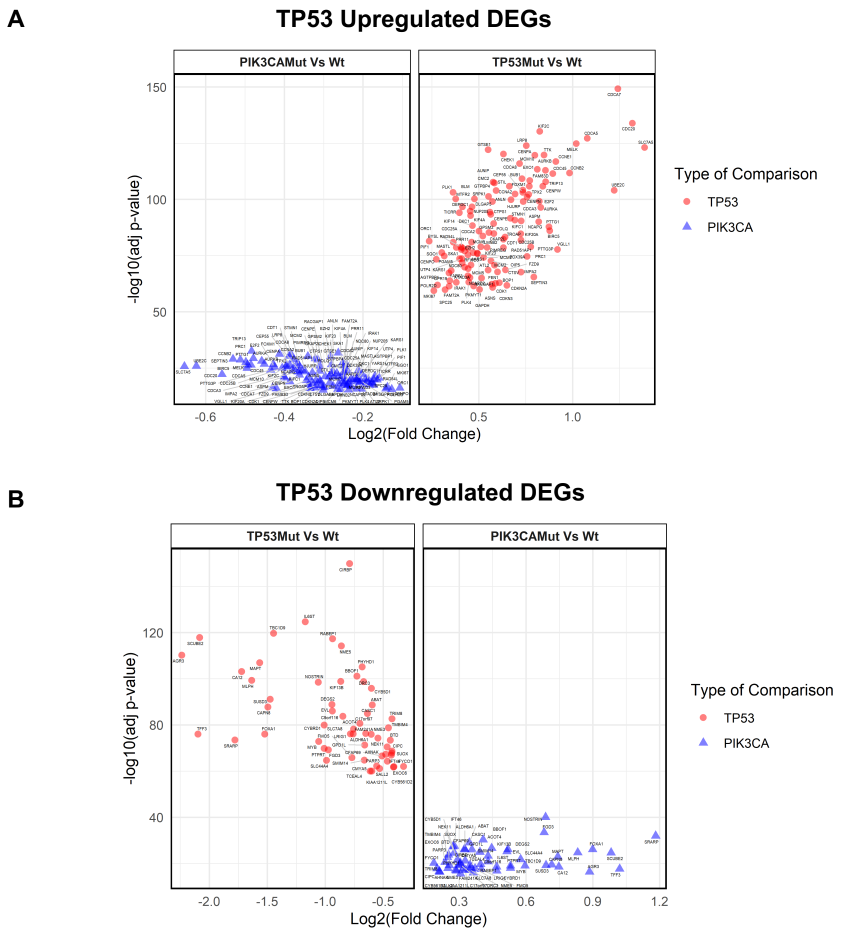


**Supplementary Figure 2 | Overlapping significantly altered genes display opposing transcriptional trends in TP53 and PIK3CA mutant contexts**. Genes overlapping within the top 500 significantly altered category (FDR < 0.05) from the TP53 mutant vs wild-type and PIK3CA mutant vs wild-type comparisons in the METABRIC dataset were selected, and their differential expression patterns are displayed. Each point represents a gene, plotted with $\log_2$(fold change) on the x-axis and $-\log_{10}$(adjusted p-value) on the y-axis. DEGs upregulated in the TP53 mutant vs wild-type comparison show corresponding downregulation in the PIK3CA mutant vs wild-type comparison **(A)**, whereas DEGs downregulated in the TP53 mutant vs wild-type **(B)** exhibit a reciprocal trend of upregulation in the PIK3CA mutant vs wild-type.

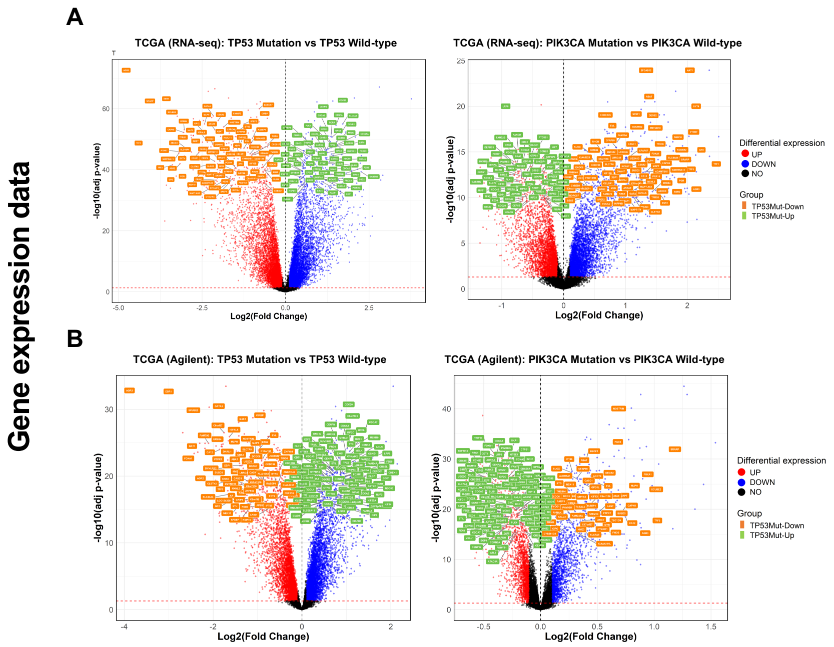


**Supplementary Figure 3 | Opposing expression patterns define TP53- and PIK3CA-driven transcriptional programs across breast cancer cohorts, agnostic of transcriptomic profiling platforms.** Volcano plots generated from **(A)** TCGA-BRCA RNA-seq and **(B)** TCGA-BRCA Agilent microarray dataset illustrate significantly altered differentially expressed genes (DEGs) for TP53-Mutant vs Wild-type (left) and PIK3CA-Mutant vs Wild-type (right). Genes with a significant fold change differential expression (FDR < 0.05) and a $\log_2$ fold change greater than 0 are depicted as blue dots, and those with a log2 fold change less than 0 are shown as red dots. Those that fall under the not-significant category are depicted as black dots. Horizontal and vertical dashed lines indicate the thresholds for statistical significance and fold change, respectively. Among the top 500 significantly differentially expressed genes (FDR < 0.05), those overlapping between TP53 and PIK3CA comparisons are labelled, showing consistent reciprocal regulation across the datasets using different transcriptome profiling platforms: genes upregulated in TP53-mutant tumours (gene names highlighted in green) are downregulated in PIK3CA-mutant tumours. In contrast, genes downregulated in TP53-mutant tumours (highlighted in orange) are upregulated in PIK3CA-mutant tumours (*see Supplementary Table 4*), emphasising the contrasting transcriptional programs driven by these two major mutations.

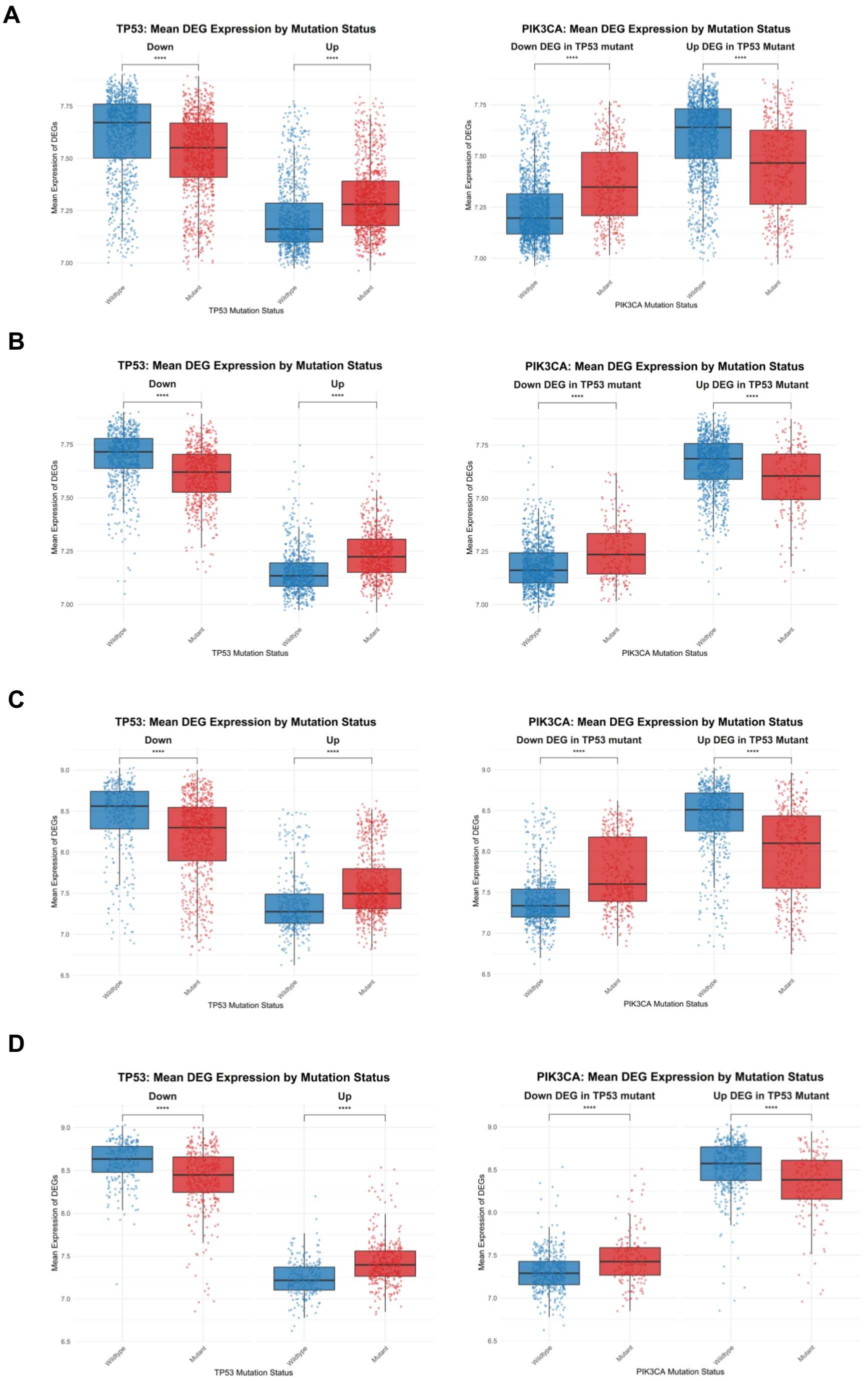


**Supplementary Figure 4 | TP53- and PIK3CA-mutation–associated transcriptional programmes define two distinct genomic arms in breast cancer across cohorts and subtypes.** Boxplots show the distribution of a per-tumour TP53 DEG-set expression score (y-axis; calculated as the mean expression of genes differentially expressed between TP53-mutant and TP53–wild-type tumours) across tumours stratified by TP53 and PIK3CA mutation status (x-axis). Each dot represents one tumour sample. Differentially expressed genes (DEGs; FDR < 0.05) were defined from the TP53-mutant vs TP53–wild-type comparison. Scores are shown for all subtypes **(A,C)** and for the ER+/HER2− subtype **(B,D)** in METABRIC and TCGA-BRCA, respectively. Downregulated and upregulated TP53-associated DEG sets are shown in separate panels (left: genes downregulated in TP53-mutant tumours; right: genes upregulated in TP53-mutant tumours). Statistical significance between wild-type and mutant groups ($p < 0.0001$) is indicated by ***. Across cohorts and within ER+/HER2− disease, PIK3CA-mutant tumours exhibit expression patterns opposite to the TP53-mutant programme.

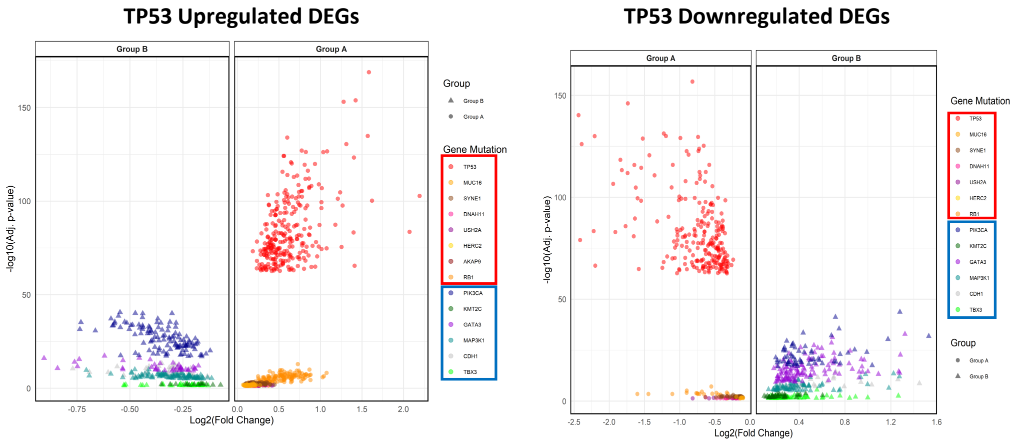


**Supplementary Figure 5 | Downstream transcriptomic effects of recurrent breast cancer alterations align with TP53- or PIK3CA-associated programmes.** The 25 most frequently mutated genes in the METABRIC cohort were identified, and for each gene, differential expression was performed comparing mutant versus wild-type tumours. For each mutation, DEGs were intersected with the TP53-associated reference gene sets, defined as the top 500 significant genes (FDR < 0.05) from the TP53-mutant vs TP53–wild-type analysis, split into TP53-upregulated and TP53-downregulated sets. Scatter plots show the overlapping DEGs, with $\log_2$(fold change) on the x-axis and $-\log_{10}$(adjusted *p*-value**)** on the y-axis. The left panel shows the transcriptomic alignment with the TP53-upregulated set, and the right panel shows alignment with the TP53-downregulated set. Only recurrently mutated genes with non-zero overlap are displayed. Points are shaped according to the Group A/Group B classification (corresponding to TP53-like vs PIK3CA-like alignment), and colours denote the recurrently mutated gene defining each comparison.

### Supplementary Figure 6

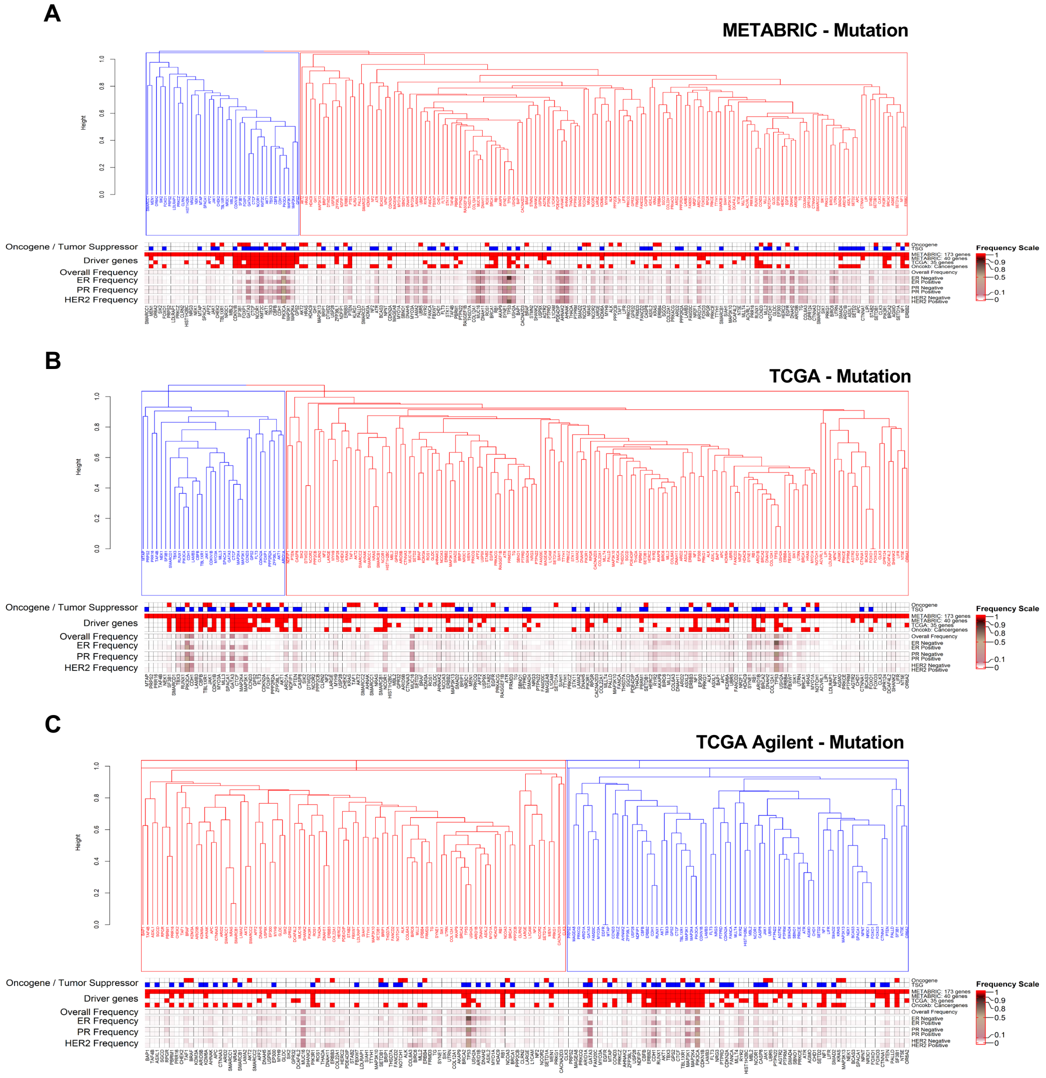


**Supplementary Figure 6 | Dendrograms reveal two divergent transcriptional arms from mutation vs wild-type analyses across breast cancer cohorts.** Dendrograms show hierarchical clustering of $\log_2$ fold-change (FC) profiles derived from differential expression comparisons between each recurrent gene mutation and wild-type (Mut vs Wt) across three independent transcriptomic datasets: **(A)** METABRIC, **(B)** TCGA-BRCA RNA-seq, and, **(C)** TCGA-BRCA Agilent. Each branch represents the gene's Mut vs Wt comparison, and clustering is based on similarity of the corresponding FC profile. The y-axis represents clustering height, reflecting the degree of similarity in transcriptional alteration patterns among recurrent genes. Branch colours denote the two major clusters, primarily associated with TP53 (red) and PIK3CA (blue). Below each dendrogram, a heatmap summarises gene-level annotations, including functional class (oncogene vs tumour suppressor), prior designation as a driver gene in published studies, and alteration frequency within each cohort. Additional panels show overall alteration frequencies and subtype-specific frequencies by receptor status (ER, PR, HER2).

### Supplementary Figure 7

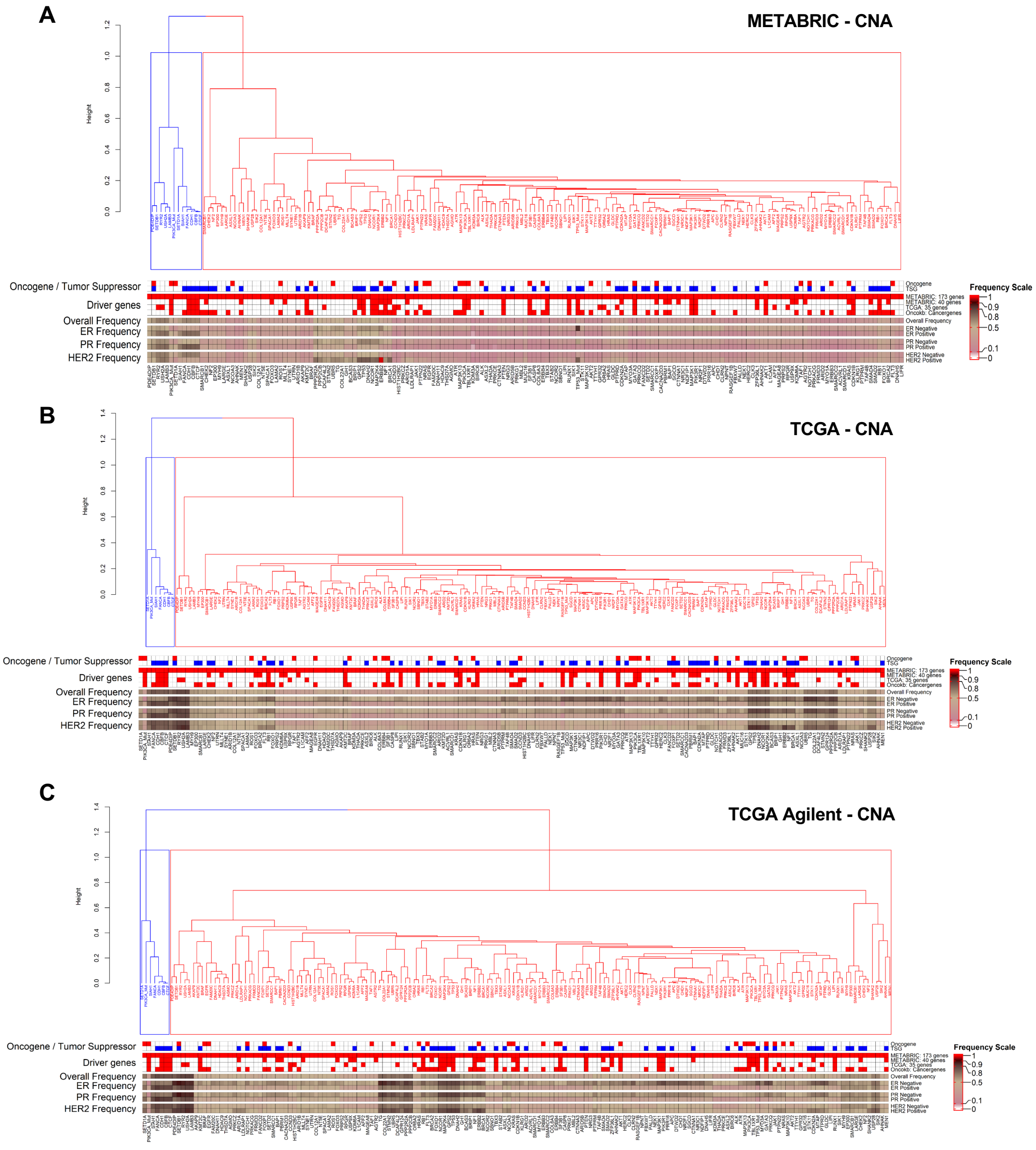


**Supplementary Figure 7 | Dendrograms show that recurrent CNAs exhibit TP53-like downstream transcriptional directionality irrespective of CNA polarity.** Dendrograms show hierarchical clustering of $\log_2$ fold-change (FC) profiles derived from differential expression comparisons between each recurrent gene copy number alteration and copy number-neutral state across three independent transcriptomic datasets: **(A)** METABRIC, **(B)** TCGA-BRCA RNA-seq, and **(C)** TCGA-BRCA Agilent dataset. Each branch represents the gene's CNA versus copy-number-neutral comparison, and clustering is based on the similarity of the corresponding FC profile. FC profiles from TP53-mutant vs TP53–wild-type and PIK3CA-mutant vs PIK3CA–wild-type analyses are included as reference signatures to anchor Group A (TP53-like) and Group B (PIK3CA-like) clusters. The y-axis indicates clustering height, reflecting the degree of similarity in transcriptional alteration patterns among recurrent genes. Branch colours denote the two clusters, primarily associated with TP53 (red) and PIK3CA (blue) mutations. Below each dendrogram, a heatmap summarises gene-level annotations, including functional class (oncogene vs tumour suppressor), prior designation as a driver gene in published studies, and alteration frequency within each cohort. Additional panels show overall alteration frequencies and subtype-specific frequencies by receptor status (ER, PR, HER2).

### Supplementary Figure 8

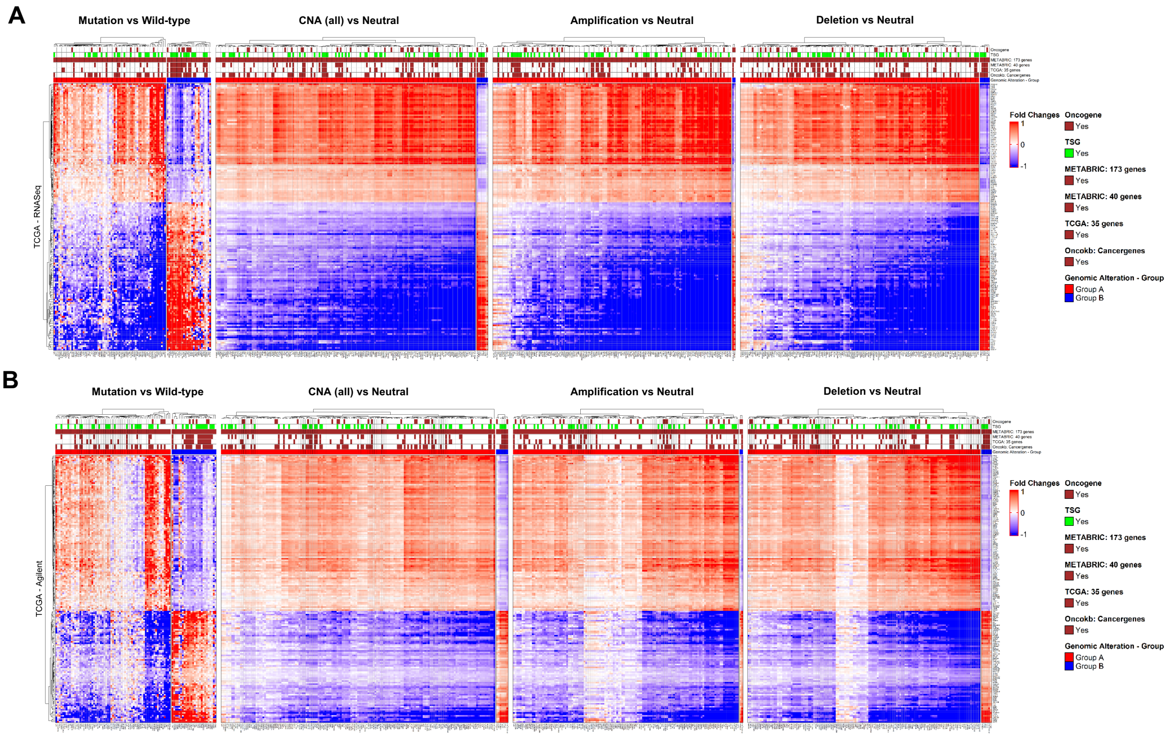


**Supplementary Figure 8 | Heatmaps show contrasting transcriptional programmes between Group A and Group B alterations in the TCGA-BRCA cohort.** Heatmaps depict $\log_2$ fold-change patterns of selected DEGs across recurrent alterations in **(A)** TCGA-BRCA RNA-seq and **(B)** TCGA-BRCA Agilent expression datasets. For each alteration class—mutation**,** CNA (amplifications and deletions combined)**,** amplification, and deletion—downstream transcriptional impact was quantified by comparing genomically altered samples (mutant or copy-number altered) with their corresponding non-altered controls (wild-type or copy-number neutral). The colour scale indicates the $\log_2$ fold-change magnitude (legend). DEGs were defined as the intersection of the top 500 significant genes **(**FDR < 0.05**)** from the TP53-mutant vs TP53–wild-type and PIK3CA-mutant vs PIK3CA–wild-type comparisons. Across alteration types, DEG fold-change patterns are consistent but show opposite directionality between Group A (TP53-like) and Group B (PIK3CA-like) recurrent alterations. Columns METABRIC_40**,** Genes_35**,** METABRIC_173**,** and OncoKB_CancerGenes indicate membership in previously published or curated gene signatures (*Supplementary Table 2*).

### Supplementary Figure 9

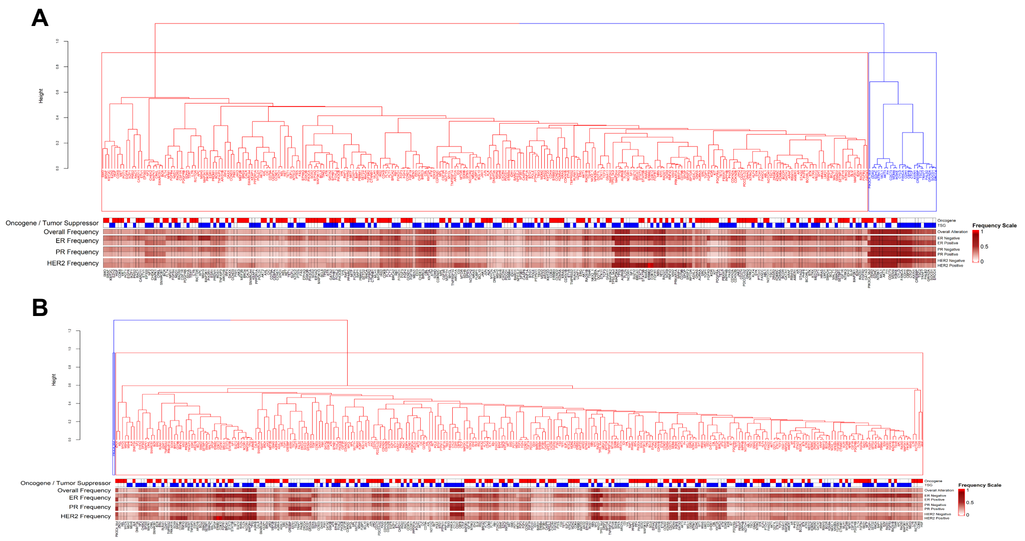


**Supplementary Figure 9 | Dendrograms show that recurrent CNAs exhibit TP53-like downstream transcriptional directionality across breast cancer subtypes.** Dendrograms show hierarchical clustering of $\log_2$ fold-change (FC) profiles derived from differential expression comparisons between each recurrent gene copy number alteration and the copy-number-neutral state across the METABRIC dataset, for all cancer genes listed in Supplementary Table 2, separately for ER-positive **(A)** and ER-negative **(B)** samples. Each branch represents the gene's CNA versus copy-number-neutral comparison, and clustering is based on the similarity of the corresponding FC profile. FC profiles from TP53-mutant vs TP53–wild-type and PIK3CA-mutant vs PIK3CA–wild-type analyses are included as reference signatures to anchor Group A (TP53-like) and Group B (PIK3CA-like) clusters. The y-axis indicates clustering height, reflecting the degree of similarity in transcriptional alteration patterns among recurrent genes. Branch colours denote the two clusters, primarily associated with TP53 (red) and PIK3CA (blue) mutations. Below each dendrogram, a heatmap summarises gene-level annotations, including functional class (oncogene vs tumour suppressor), prior designation as a driver gene in published studies, and alteration frequency within each cohort. Additional panels show overall alteration frequencies and subtype-specific frequencies by receptor status (ER, PR, HER2).

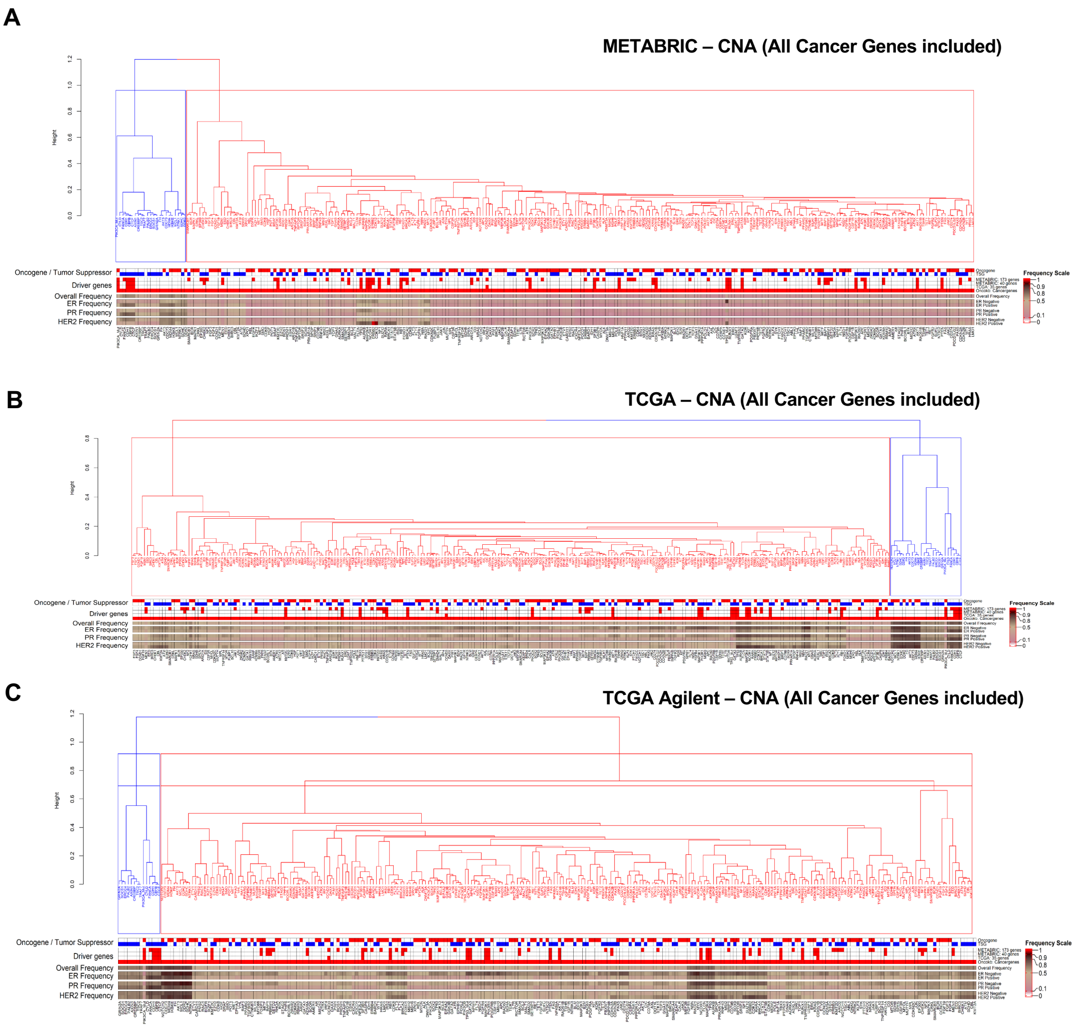


**Supplementary Figure 10 | An extended analysis with all cancer-related genes shows that recurrent CNAs exhibit TP53-like downstream transcriptional directionality irrespective of CNA polarity.** Dendrograms show hierarchical clustering of $\log_2$ fold-change (FC) profiles derived from differential expression comparisons between each recurrent gene copy number alteration and the copy-number-neutral state for all cancer genes listed in Supplementary Table 2 across three independent transcriptomic datasets: **(A)** METABRIC, **(B)** TCGA-BRCA RNA-seq, and **(C)** TCGA-BRCA Agilent dataset. Each branch represents the gene's CNA versus copy-number-neutral comparison, and clustering is based on the similarity of the corresponding FC profile. FC profiles from TP53-mutant vs TP53–wild-type and PIK3CA-mutant vs PIK3CA–wild-type analyses are included as reference signatures to anchor Group A (TP53-like) and Group B (PIK3CA-like) clusters. The y-axis indicates clustering height, reflecting the degree of similarity in transcriptional alteration patterns among recurrent genes. Branch colours denote the two clusters, primarily associated with TP53 (red) and PIK3CA (blue) mutations. Below each dendrogram, a heatmap summarises gene-level annotations, including functional class (oncogene vs tumour suppressor), prior designation as a driver gene in published studies, and alteration frequency within each cohort. Additional panels show overall alteration frequencies and subtype-specific frequencies by receptor status (ER, PR, HER2).

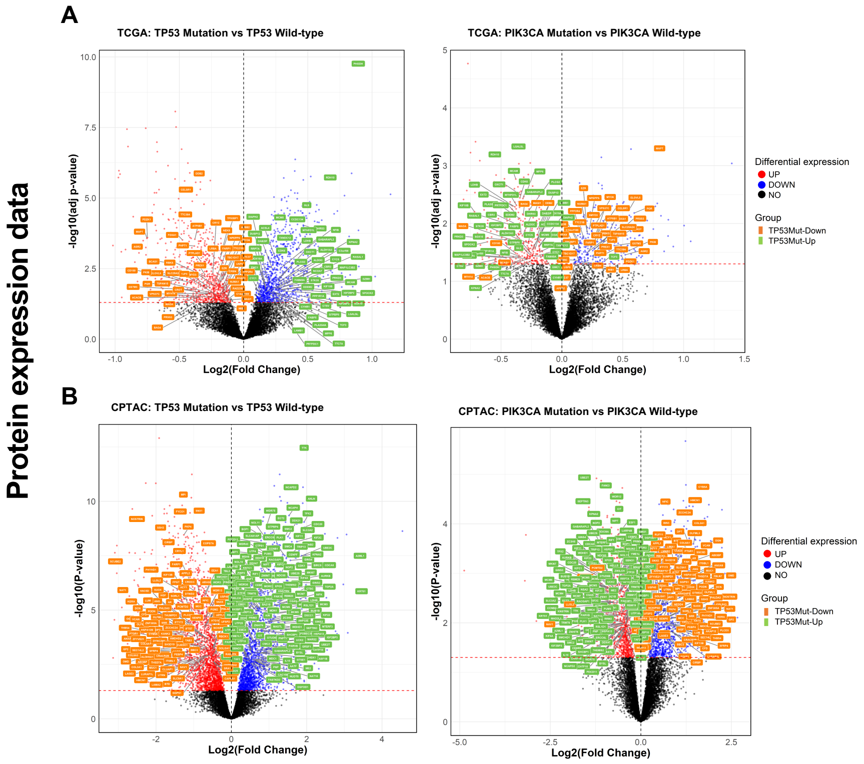


**Supplementary Figure 11 | Antagonistic TP53- and PIK3CA-associated programmes are also observed at the proteomic level across breast cancer cohorts.** Volcano plots show proteins from TP53-Mutant vs Wild-type (left) and PIK3CA-Mutant vs Wild-type (right) comparisons, with $\log_2$ fold change on the x-axis and $-\log_{10}$(adj *P* value) on the y-axis. Each point represents one protein. Proteins significantly upregulated in mutant tumours are shown in blue, downregulated proteins in red, and non-significant proteins in black. Dashed horizontal and vertical lines indicate significance and fold-change thresholds, respectively. Among significantly altered proteins (FDR < 0.05), proteins significant in both TP53-mutant vs TP53–wild-type and PIK3CA-mutant vs PIK3CA–wild-type comparisons are highlighted**:** orange-labelled proteins are those downregulated in TP53-mutant tumours, and green-labelled proteins are those upregulated in TP53-mutant tumours. Plots generated from **(A)** TCGA proteomic data and **(B)** CPTAC breast cancer proteomic data show opposing protein-level changes between TP53- and PIK3CA-mutant tumours, mirroring the transcriptional dichotomy observed in Figures 1C and S3.

### Supplementary Figure 12

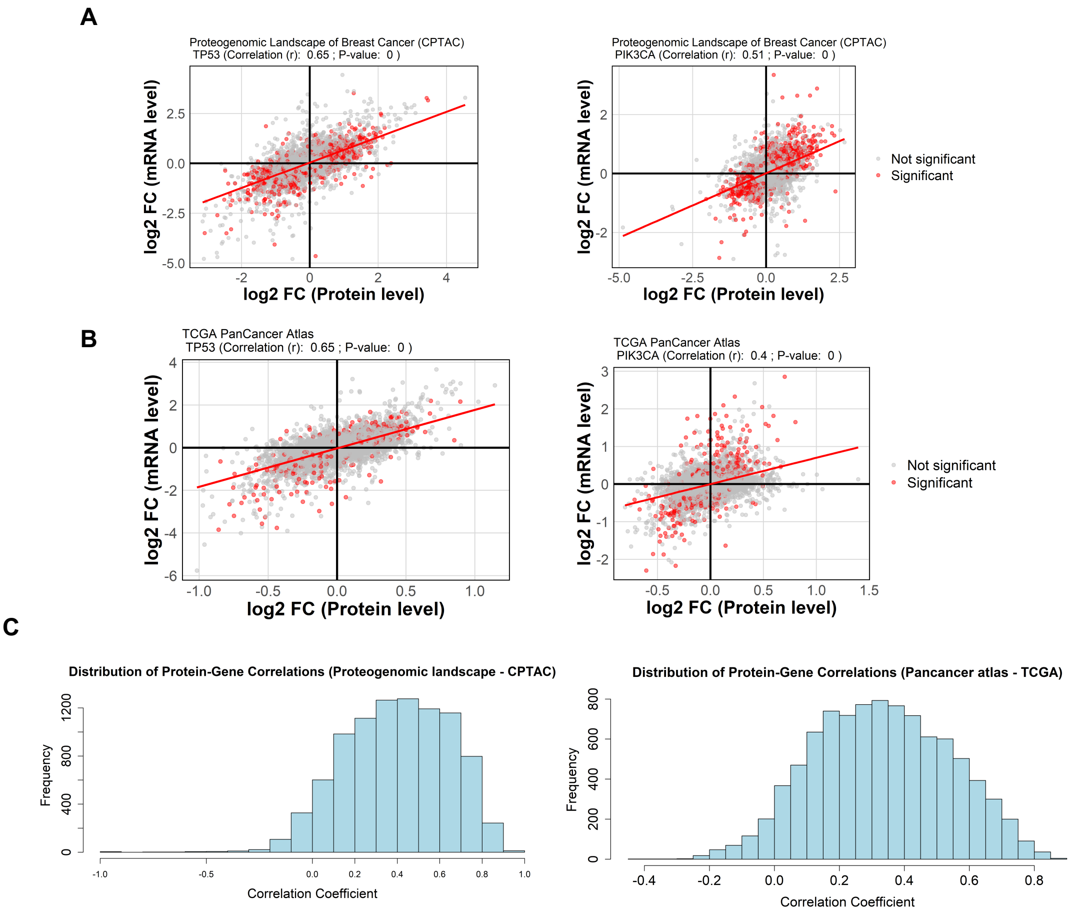


**Supplementary Figure 12 | mRNA–protein concordance of TP53- and PIK3CA-associated expression changes in breast cancer**. **(A–B)** Scatter plots show the relationship between protein-level and mRNA-level expression changes **for** TP53-mutant vs TP53–wild-type and PIK3CA-mutant vs PIK3CA–wild-type comparisons in **(A)** CPTAC breast cancer and **(B)** TCGA PanCancer Atlas (BRCA) datasets. Each point represents a gene, with $\log_2$ fold change (FC) at the protein level on the x-axis and the corresponding $\log_2$ FC at the mRNA level on the y-axis. Overall, mRNA and protein fold changes are positively correlated, suggesting that transcript-level deregulation is reflected at the proteomic level. Genes significantly deregulated ($p < 0.05$) in mutant tumours and significant in both the TP53 and PIK3CA comparisons are highlighted. **(C)** Histograms summarise the distribution of mRNA and protein $\log_2$ FC values, further supporting concordant directional trends across molecular layers. Collectively, these analyses indicate that TP53- and PIK3CA-associated transcriptional changes are mirrored at the protein level across cohorts.

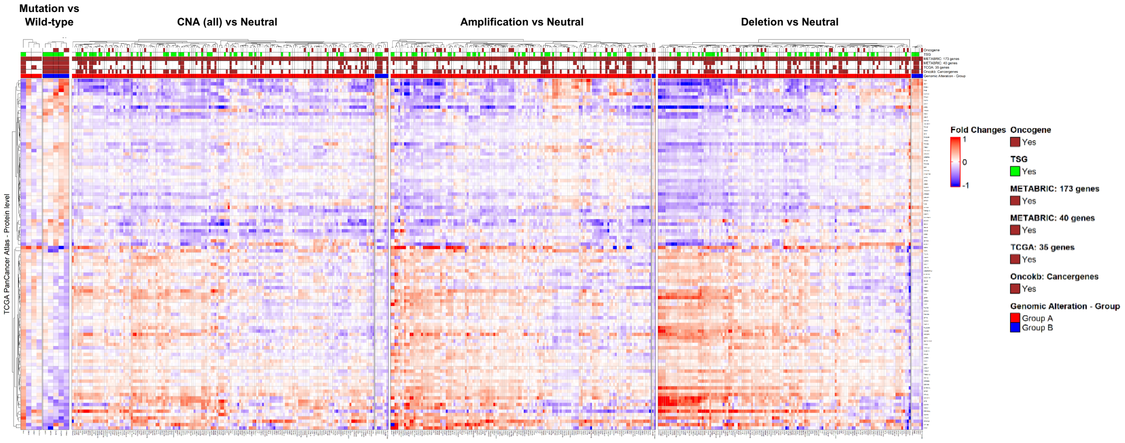


**Supplementary Figure 13 | Heatmaps show contrasting proteomic programmes between Group A and Group B alterations in the TCGA Pan-Cancer Atlas breast cancer cohort.** Heatmaps depict $\log_2$ proteomic fold-change patterns from mass spectrometry–quantified protein abundance across recurrent alterations. For each alteration class—mutation, CNA (amplifications and deletions combined), amplification, and deletion—proteomic impact was quantified by comparing genomically altered samples (mutant or copy-number altered) with their corresponding non-altered controls (wild-type or copy-number neutral). The colour scale indicates the log2 fold-change magnitude (legend). Proteins were defined as the intersection of the top 500 significantly altered proteins **(**$FDR < 0.05$**)** from the TP53-mutant vs TP53–wild-type and PIK3CA-mutant vs PIK3CA–wild-type proteomic comparisons. Across alteration types, fold-change patterns are consistent but show opposite directionality between Group A (TP53-like) and Group B (PIK3CA-like) recurrent alterations. Columns METABRIC_40**,** Genes_35**,** METABRIC_173, and OncoKB_CancerGenes indicate membership in previously published or curated gene signatures (Supplementary Table 2).

### Supplementary Figure 14

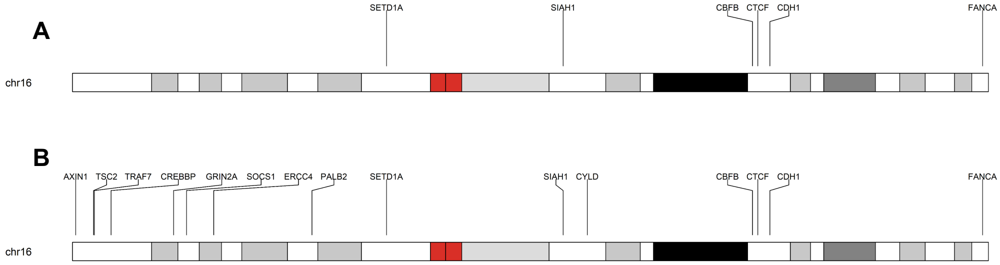


**Supplementary Figure 14 | Enrichment of Group B Copy Number Alteration–Associated Genes on Chromosome 16.** Group B genes identified from the copy number alteration versus neutral analysis show a pronounced enrichment on chromosome 16. Panel **(A)** presents results from the analysis of recurrently altered genes (Figure S7), while panel **(B)** shows the corresponding enrichment of Group B-related genes across chromosome 16 from the complete set of cancer-associated genes analysed (Figure S10).

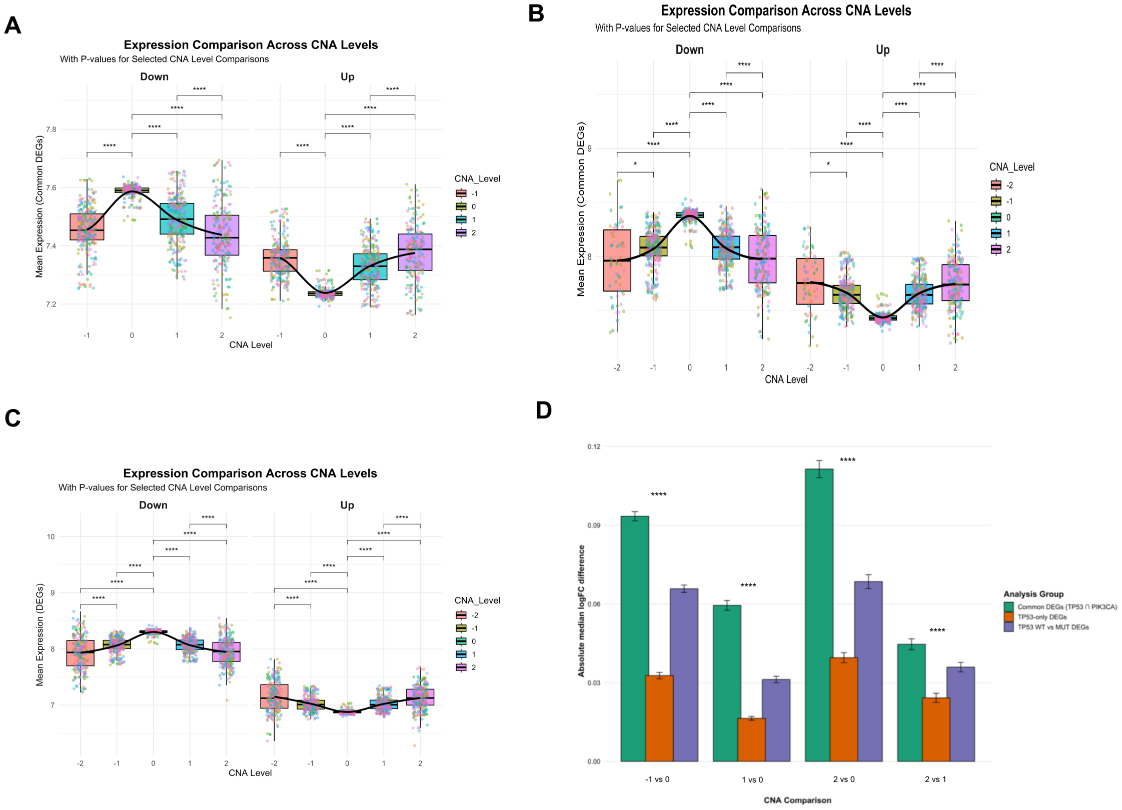


**Supplementary Figure 15 | CNA dosage shows a monotonic strengthening of the TP53-like transcriptional programme.** Mean expression levels of differentially expressed genes (DEGs) shared between TP53- and PIK3CA-mutant vs wild-type comparisons are plotted against corresponding copy number alteration (CNA) levels for the METABRIC **(A)** and TCGA **(B)** cohorts. Expression of all DEGs identified in the TP53-mutant vs TP53 wild-type comparison is shown for the TCGA cohort **(C)**. CNA levels (Copy -1, 0, 1, 2) of cancer-associated genes (derived from OncoKb; each dot in the boxplot represents a cancer-associated gene) are shown on the x-axis, with a loess-smoothed curve overlaid to illustrate overall trends. The y-axis shows $\log_2$-normalized mean DEG expression. UP and DOWN DEGs are shown in two separate panels (left: DEGs downregulated in TP53 mutant tumours, right: DEGs upregulated in TP53 mutant tumours). For METABRIC, CNA levels −2 and −1 were merged due to limited −2 samples, and +2 alterations were included only if supported by ≥10 samples (≥0.5% of cohort). Statistical significance across CNA levels was assessed by Student's t-test; significance thresholds are indicated (*$p < 0.05$; **$p < 0.01$; ***$p < 0.001$; ****$p < 0.0001$). All tumor samples available in each cohort were included in these analyses. Across CNA states, the TP53-like programme increases monotonically with CNA magnitude, peaking in deep losses and high-level gains, indicating that both copy losses and gains of cancer-associated genes can produce TP53-mutant–like transcriptomic patterns (see also Supplementary Figure 4). **(D)** Bar plot showing the absolute median $\log_2$ fold change ($\log_2$FC) for three DEG categories: (1) intersection DEGs shared by TP53 and PIK3CA comparisons, (2) TP53-specific DEGs (not shared with PIK3CA), and (3) all TP53-associated DEGs. Values are shown for CNA subgroup contrasts (−1 vs 0, 1 vs 0, 2 vs 0, and 1 vs 2). Bar heights represent the absolute median $\log_2$FC within each CNA contrast, reflecting the magnitude of transcriptional differences independent of direction

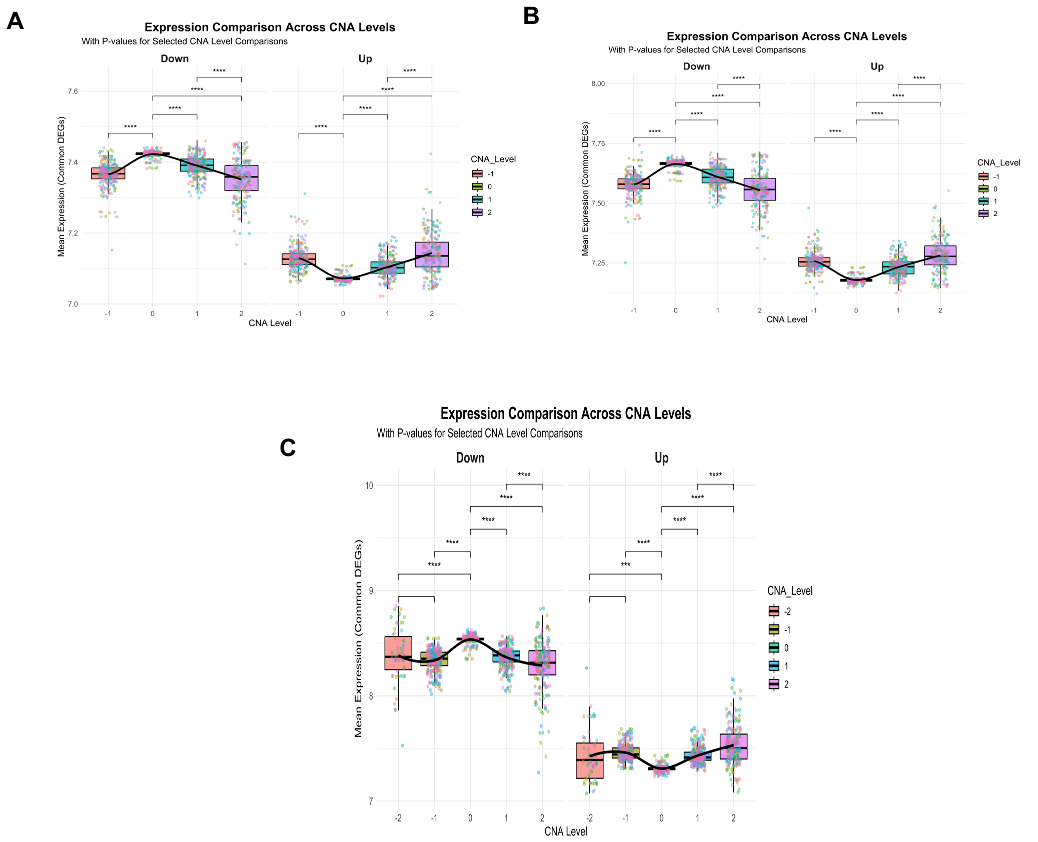


**Supplementary Figure 16 | An extension of Figure S15 showing the reproducibility of CNA dosage exhibiting a monotonic strengthening of the TP53-like transcriptional programme in ER+/HER2– tumours.** Mean expression of all significant DEGs (*FDR < 0.05*) identified in the TP53-mutant vs TP53 wild-type comparison is plotted against CNA levels for cancer-associated genes in the METABRIC ER+/HER2– breast cancer cohort **(A)**. Mean expression of DEGs shared between TP53- and PIK3CA-mutant vs wild-type comparisons are plotted against CNA levels in the METABRIC **(B)** and TCGA **(C)** ER+/HER2– cohorts. CNA levels (Copy -1, 0, 1, 2) of cancer-associated genes (derived from OncoKb; each dot in the boxplot represents a cancer-associated gene) are shown on the x-axis, with a loess-smoothed curve overlaid to illustrate overall trends. The y-axis shows $log_2$-normalized mean DEG expression. UP and DOWN DEGs are shown in two separate panels (left: DEGs downregulated in TP53 mutant tumours, right: DEGs upregulated in TP53 mutant tumours). For METABRIC, CNA levels −2 and −1 were merged due to limited −2 samples, and +2 alterations were included only if supported by ≥10 samples (≥0.5% of cohort). Statistical significance across CNA levels was assessed by Student's t-test; significance thresholds are indicated (*$p < 0.05$; **$p < 0.01$; ***$p < 0.001$; ****$p < 0.0001$). All tumor samples available in each cohort were included in these analyses. Across CNA states, the TP53-like programme increases monotonically with CNA magnitude, peaking in deep losses and high-level gains, indicating that both copy losses and gains of cancer-associated genes can produce TP53-mutant–like transcriptomic patterns in ER+/HER2– breast cancer (see also Supplementary Figure 4).

### Supplementary Figure 17

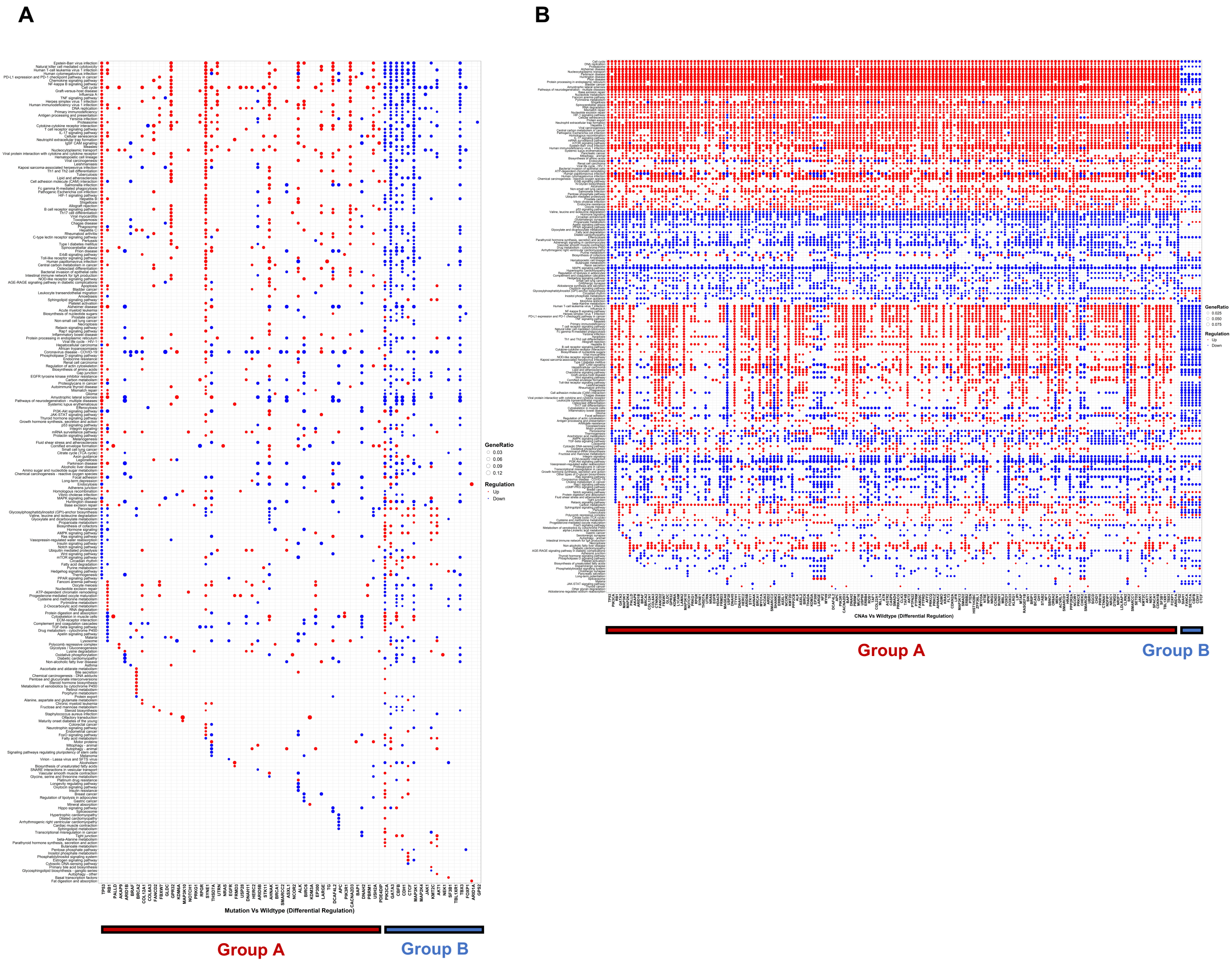


**Supplementary Figure 17 | Summary of KEGG gene sets enrichment in the METABRIC cohort. (A)** KEGG gene set enrichment from differential expression analyses comparing mutant vs wild-type samples for Group A and Group B genes from Figure 1C, visualised as a dot plot. Each column corresponds to a Mutation vs Wild-type comparison, and each row represents a significantly enriched KEGG gene set. Red and blue indicate enrichment of upregulated and downregulated KEGG gene sets, respectively. Two major transcriptional patterns are annotated along the x-axis: Group A (gene alterations showing TP53-like transcriptional directionality and enriched for the canonical proliferative/replicative programme) and Group B (showing PIK3CA-like directionality and an opposing molecular trend to TP53 mutant tumours, influencing non-canonical signalling/cell-state programmes rather than directly accelerating proliferation/DNA replication). **(B)** KEGG pathway enrichment for copy-number altered (CNA) versus copy-number neutral samples is shown using a similar dot-plot layout, where each column represents a CNA comparison, and each row represents enriched KEGG gene sets.

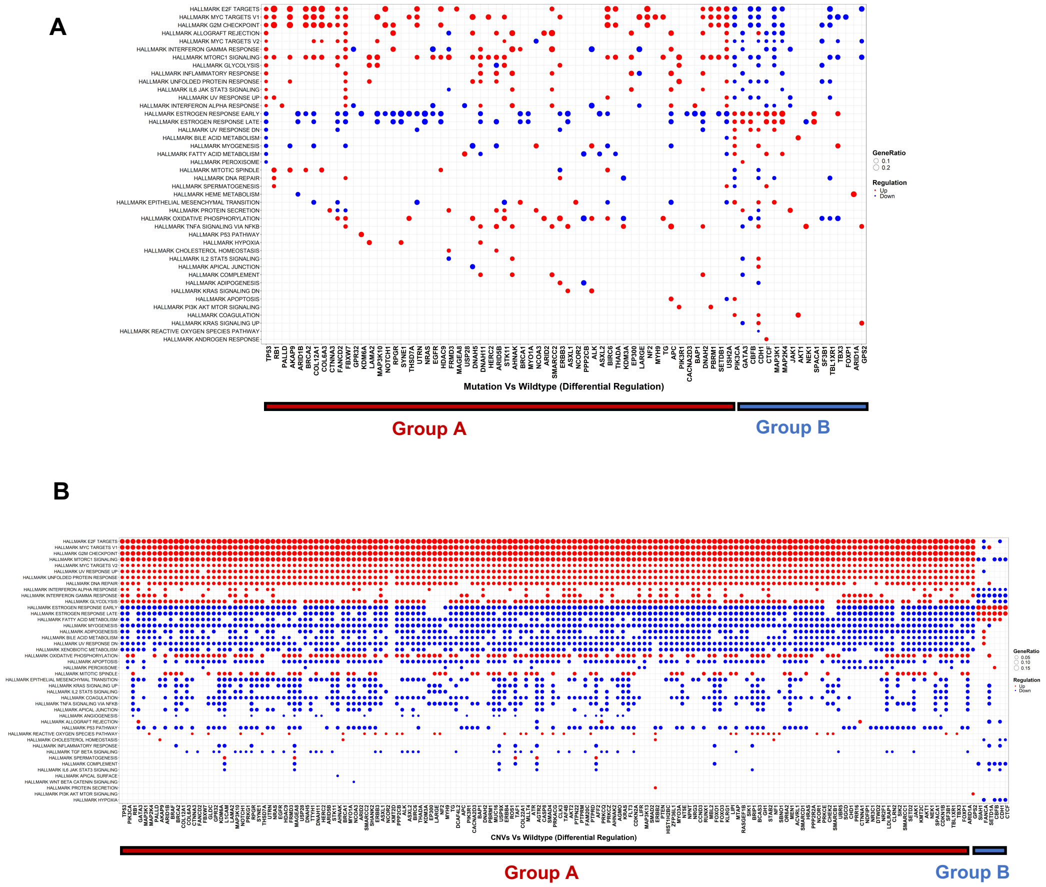


**Supplementary Figure 18 | Summary of Hallmark gene sets enrichment in the TCGA-BRCA cohort. (A)** Hallmark gene set enrichment from differential expression analyses comparing mutant vs wild-type samples for Group A and Group B genes from Figure 1C, visualised as a dot plot. Each column corresponds to a Mutation vs Wild-type comparison, and each row represents a significantly enriched Hallmark gene set. Red and blue indicate enrichment of upregulated and downregulated Hallmark gene sets, respectively. Two major transcriptional patterns are annotated along the x-axis: Group A (gene alterations showing TP53-like transcriptional directionality and enriched for the canonical proliferative/replicative programme) and Group B (showing PIK3CA-like directionality and an opposing molecular trend to TP53 mutant tumours, influencing non-canonical signalling/cell-state programmes rather than directly accelerating proliferation/DNA replication). **(B)** Hallmark pathway enrichment for copy-number altered (CNA) versus copy-number neutral samples is shown using a similar dot-plot layout, where each column represents a CNA comparison, and each row represents enriched Hallmark gene sets.

### Supplementary Figure 19

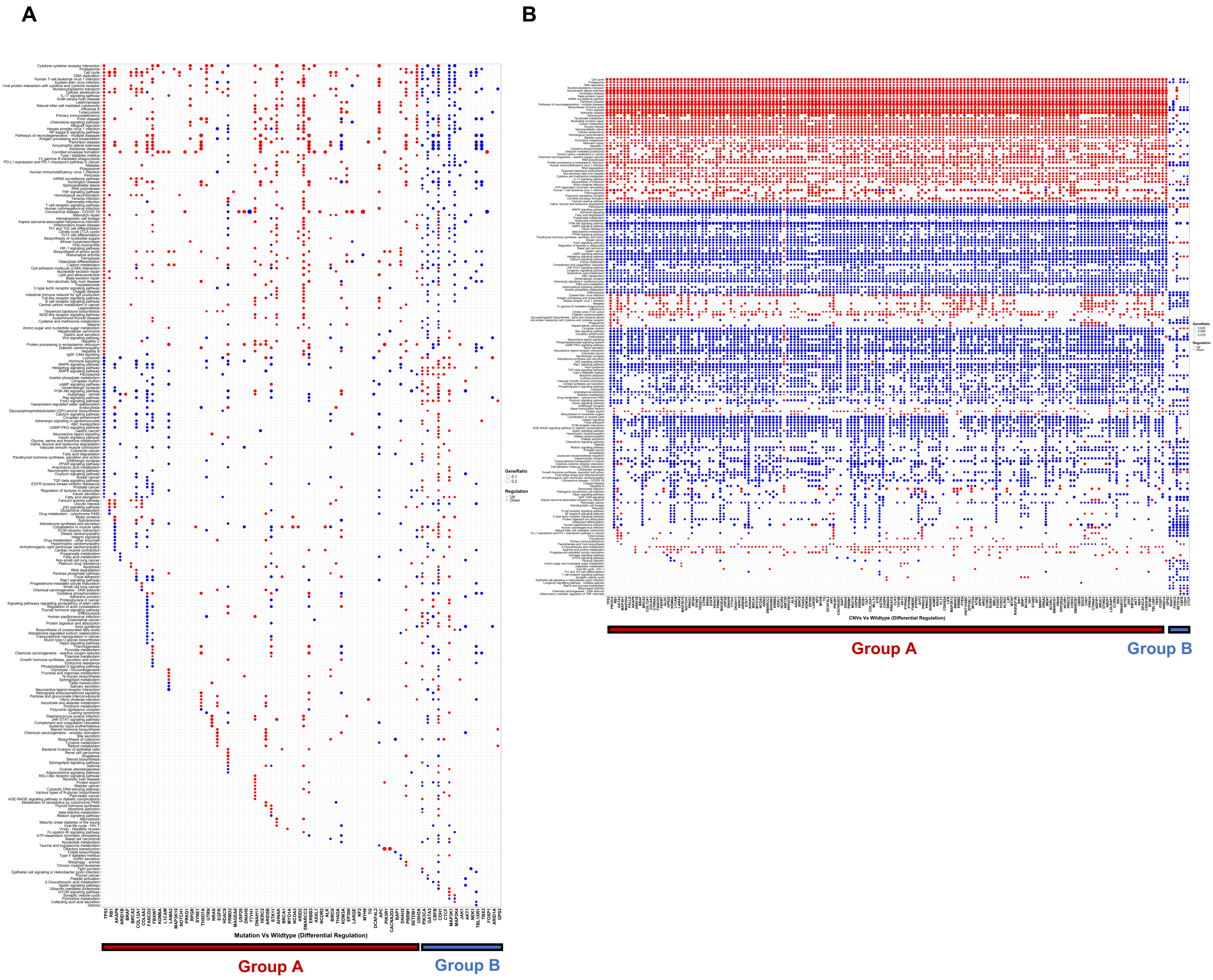


**Supplementary Figure 19 | Summary of KEGG gene sets enrichment in the TCGA-BRCA cohort. (A)** KEGG gene set enrichment from differential expression analyses comparing mutant vs wild-type samples for Group A and Group B genes from Figure 1C, visualised as a dot plot. Each column corresponds to a Mutation vs Wild-type comparison, and each row represents a significantly enriched KEGG gene set. Red and blue indicate enrichment of upregulated and downregulated KEGG gene sets, respectively. Two major transcriptional patterns are annotated along the x-axis: Group A (gene alterations showing TP53-like transcriptional directionality and enriched for the canonical proliferative/replicative programme) and Group B (showing PIK3CA-like directionality and an opposing molecular trend to TP53 mutant tumours, influencing non-canonical signalling/cell-state programmes rather than directly accelerating proliferation/DNA replication). **(B)** KEGG pathway enrichment for copy-number altered (CNA) versus copy-number neutral samples is shown using a similar dot-plot layout, where each column represents a CNA comparison, and each row represents enriched KEGG gene sets.

**A**

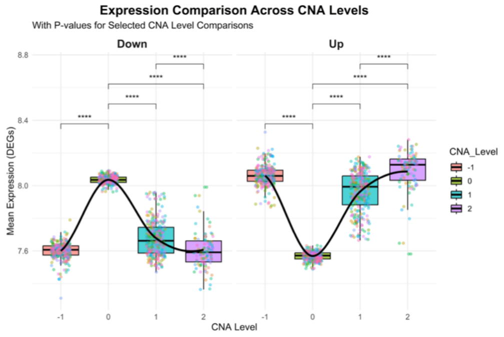


**B**

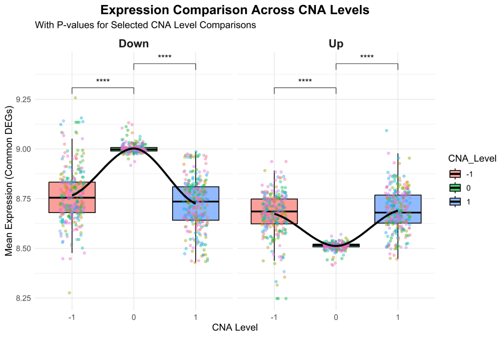


**Supplementary Figure 20 | Cancer-associated CNAs mirror TP53 mutation–like transcriptomic profiles in uterine endometrial and prostate cancer.** Mean expression of all significant DEGs (*FDR < 0.05*) identified in the TP53-mutant vs TP53 wild-type comparison is plotted against CNA levels for cancer-associated genes in the TCGA Uterine Endometrial Cancer **(A)** and TCGA Prostate Adenocarcinoma **(B)** cohorts. CNA levels (Copy -1, 0, 1, 2) of cancer-associated genes (derived from OncoKb; each dot in the boxplot represents a cancer-associated gene) are shown on the x-axis, with a loess-smoothed curve overlaid to illustrate overall trends. The y-axis shows $log_2$-normalised mean DEG expression. UP and DOWN DEGs are shown in two separate panels (left: DEGs downregulated in TP53 mutant tumours, right: DEGs upregulated in TP53 mutant tumours). In the TCGA Uterine cancer cohort, −2 was merged with −1 because most genes had 0 to very few samples at −2, and +2 was retained only for genes supported by ≥6 samples (≥1% of the cohort). For prostate cancer, both −2 and +2 CNA levels were merged with their adjacent categories (−1 and +1, respectively). This was done because extreme CNA categories were variably sparse across genes, and separating them introduced noise that obscured overall transcriptional trends. Since both −1/−2 represent gene loss and +1/+2 represent gene gain along the same biological axis, merging them emphasized robust, global transcriptomic patterns rather than rare, unstable subgroups. Statistical significance across CNA levels was assessed using Student's t-test; thresholds are indicated *(*p < 0.05; **p < 0.01; ***p < 0.001; ****p < 0.0001*).

### Supplementary Figure 21

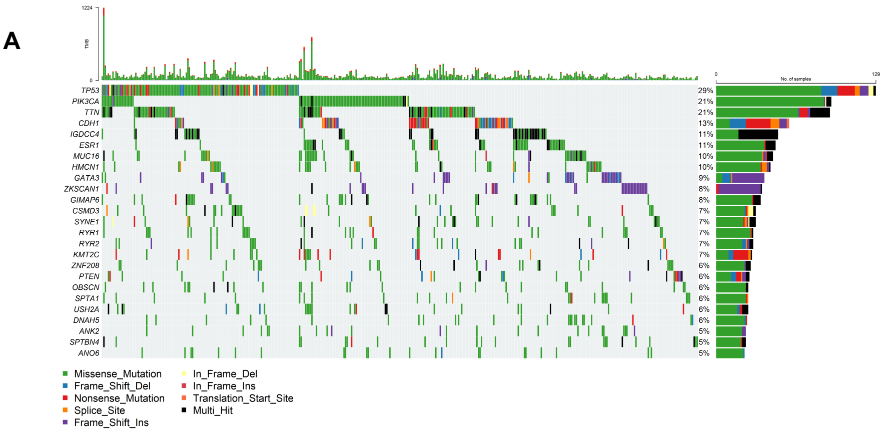


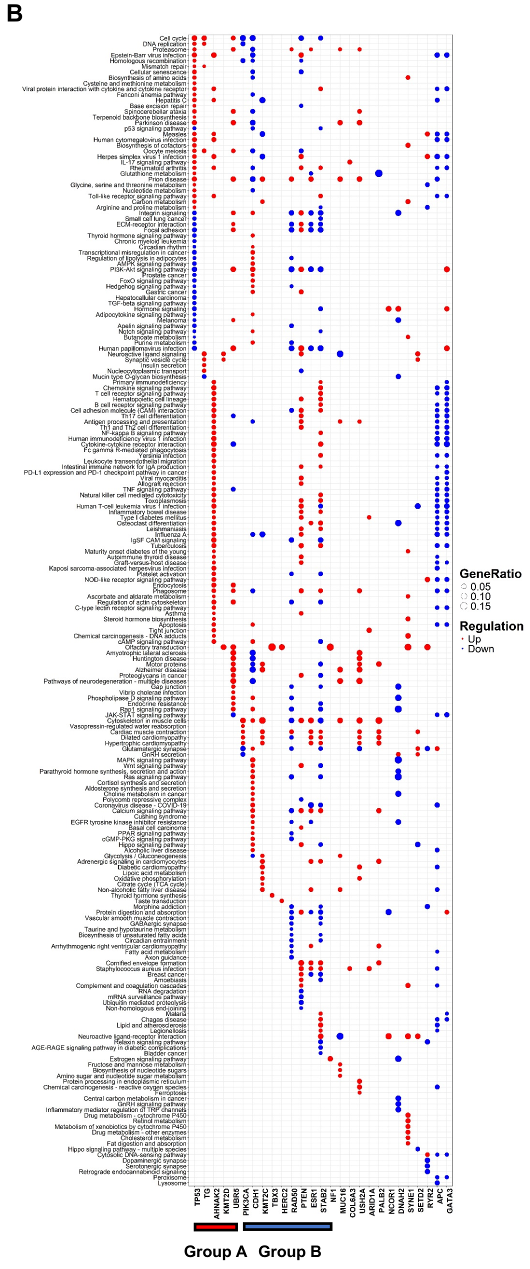


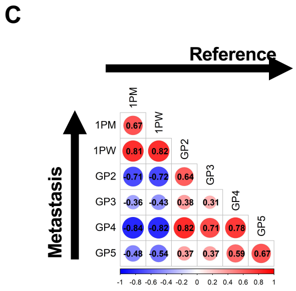


**Supplementary Figure 21 | Genomic alterations in metastatic breast cancer and agreement of pathway-activation signatures with reference dataset. (A)** Oncoplot of recurrent somatic mutations in metastatic breast cancer. The top 25 most frequently mutated genes were identified from the combined MAF files of (i) the Metastatic Breast Cancer Project (Archived, 2020) and (ii) the Metastatic Breast Cancer Project (Provisional, December 2021), and are shown as an oncoplot. **(B)** KEGG pathway gene sets enriched in differential gene expression analyses comparing mutant versus wild-type samples in metastatic breast cancer are visualised as a dot plot. Each column represents a mutation-specific comparison that passed the significance threshold ($p < 0.05$), and each row corresponds to a significantly enriched KEGG gene set. Red and blue indicate enrichment among upregulated and downregulated gene sets, respectively. **(C)** Correlation plot showing concordance between metastatic and reference ER+/HER2– cohorts across Tier 2 breast cancer subgroups. Pathway activation fold differences were independently calculated for each subgroup relative to all others using an equal-weighted approach in both the reference and metastatic cohorts (cohorts detailed in *Supplementary Table 1*). Pathways significantly activated in at least one subgroup of the metastatic dataset ($p < 0.05$) were selected. Correlations between the fold-difference values of these significant pathways in the metastatic and reference datasets are displayed.

A

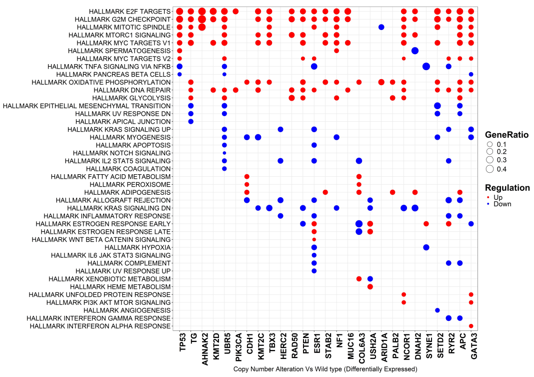


B

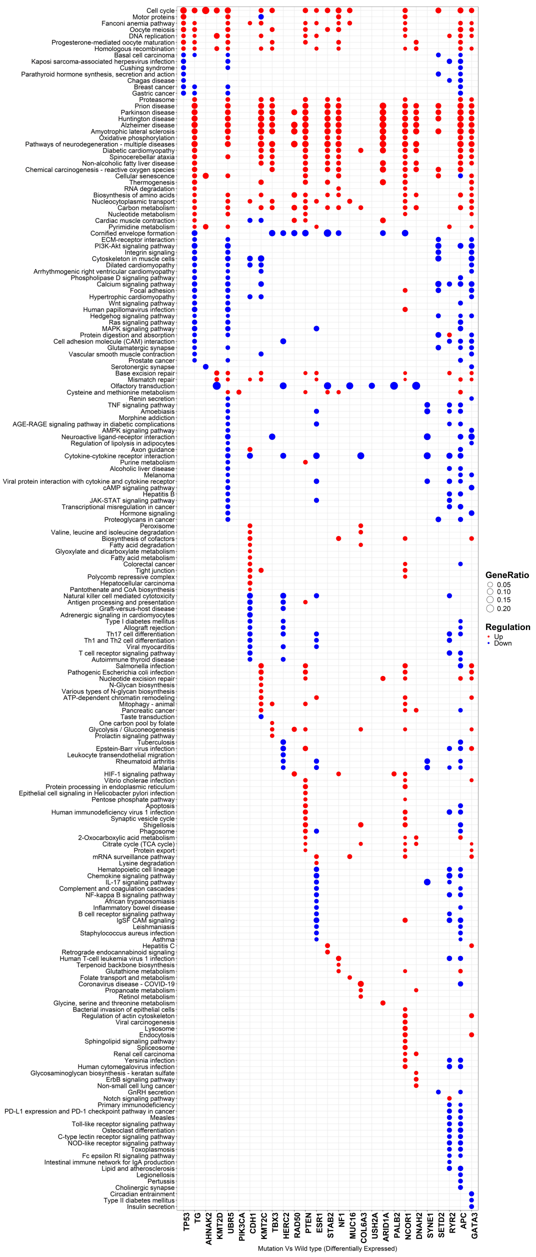


**Supplementary Figure 22 | Summary of Hallmark and KEGG gene set enrichment among copy-number-altered states in metastatic breast cancer. (A)** Hallmark pathway enrichment for copy-number altered (CNA) versus copy-number neutral samples is shown as a dot plot, with each column representing a CNA comparison and each row an enriched Hallmark gene set. **(B)** KEGG pathway enrichment for copy-number altered (CNA) versus copy-number neutral samples is shown as a dot plot, with each column representing a CNA comparison and each row an enriched KEGG gene set. Red and blue indicate enrichment of upregulated and downregulated Hallmark/KEGG gene sets, respectively.

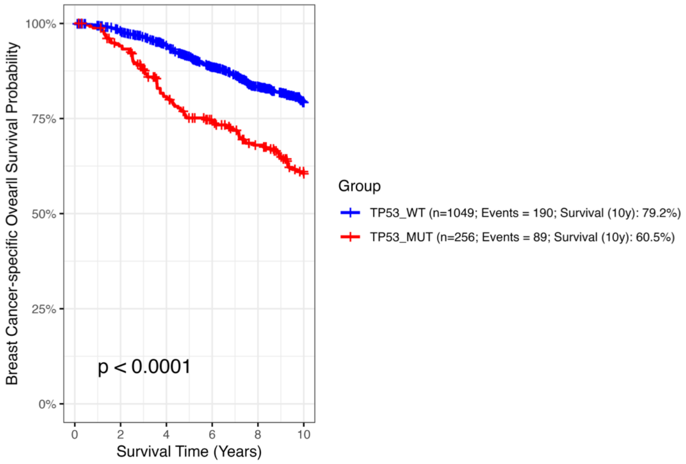


**Supplementary Figure 23 |** Kaplan-Meier (KM) survival curve showing the prognostic impact of TP53 mutations in ER+/HER2– patients from the METABRIC cohort without molecular subtyping (based on population-level averaging of TP53-mutant vs wild-type patients). Breast cancer-specific overall survival probability is shown on the y-axis. The log-rank p-value is indicated on the plot.

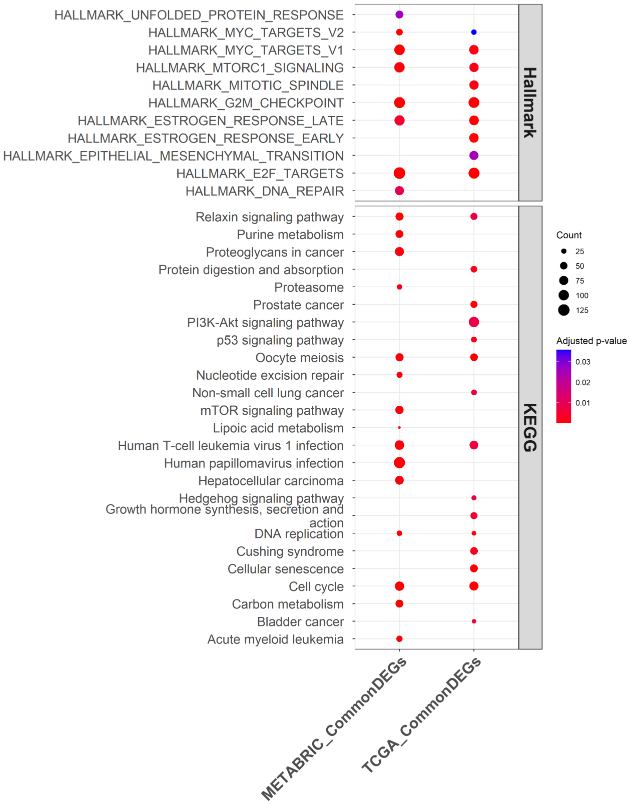


**Supplementary Figure 24 | Shared DEGs between TP53 and PIK3CA-mutant tumours are enriched for classical oncogenic programmes, including cell cycle progression and DNA replication.** Gene sets enriched among the shared DEGs between TP53- and PIK3CA-mutant tumours from the METABRIC and TCGA breast cancer cohorts are shown in a dotplot.

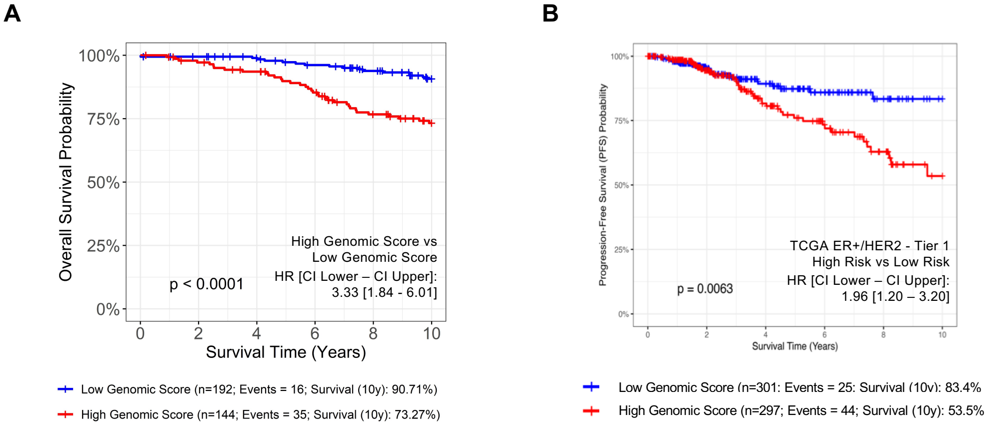


**Supplementary Figure 25 | Stratification of ER+/HER2− tumours into low- and high-genomic risk groups using the Tier 1 composite genomic risk score**. **(A)** Kaplan-Meier (KM) survival curve showing breast cancer-specific overall survival of lymph node-negative ER+/HER2– systemic therapy untreated patients (patients who have not received chemotherapy and endocrine therapy), from the METABRIC cohort. **(B)** Progression-free survival in the ER+/HER2– patients from the TCGA breast cancer cohort is shown. The survival plots display the corresponding hazard ratios (HR) and log-rank p-values within each panel.

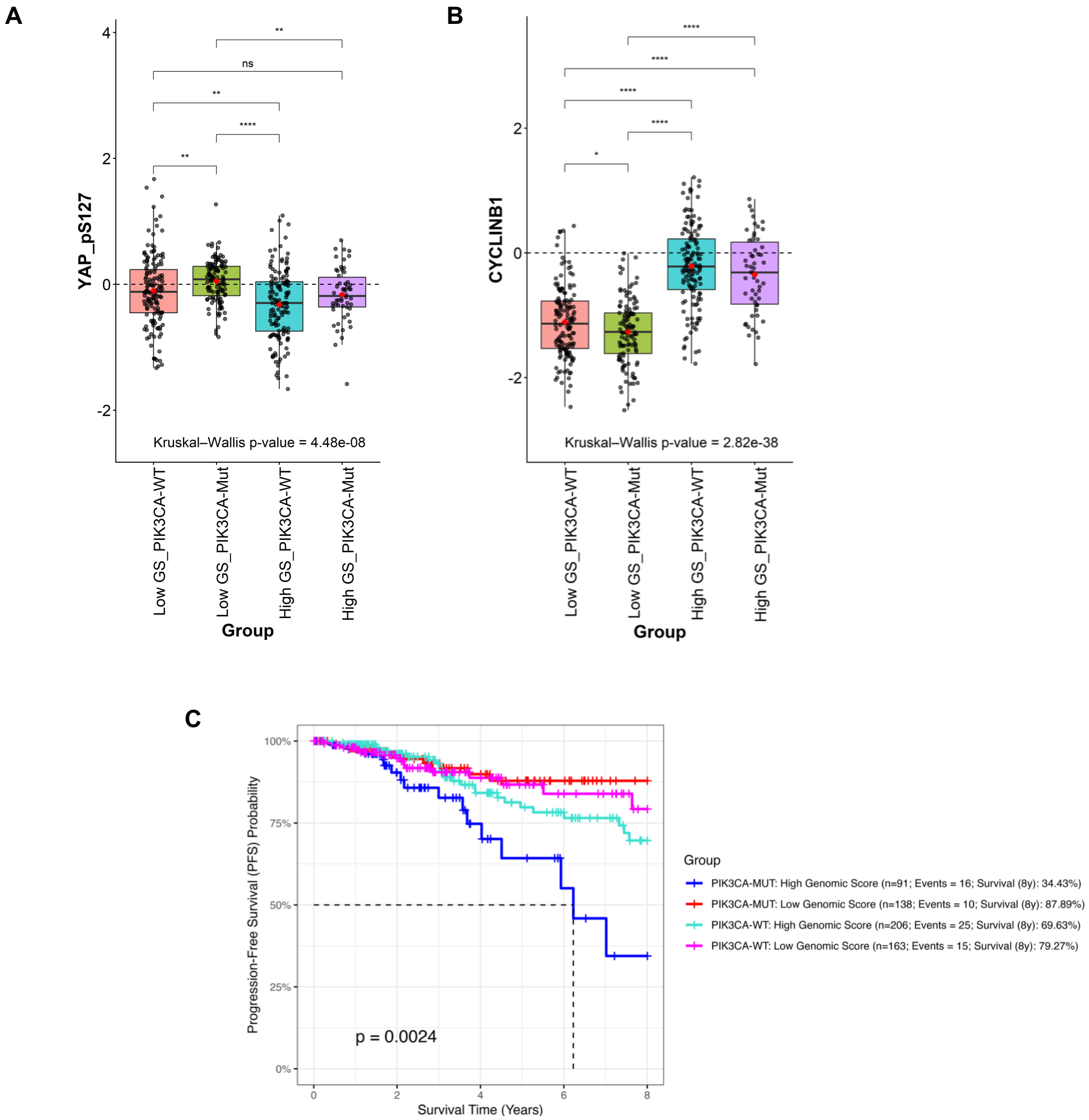


| | Coefficient | HR [CI Lower - CI Upper] | *p_value* |
|---|---|---|---|
| PIK3CA-MUT: High Genomic Score vs PIK3CA-WT: High Genomic Score | 0.88 | 2.41 [1.27 - 4.58] | 0.006 |
| PIK3CA-MUT: High Genomic Score vs PIK3CA-MUT: Low Genomic Score | 1.31 | 3.71 [1.67 - 8.24] | <0.0001 |

**Supplementary Figure 26 | Layered DNA–RNA stratification reveals that PIK3CA-mutant ER+/HER2− breast cancer comprises distinct biological states. (A–B)** Boxplots show RPPA-based protein/phosphoprotein levels of YAP1 pS127 **(A)** and Cyclin B1 **(B)** in ER+/HER2− tumours from the TCGA breast cancer cohort stratified by PIK3CA mutation status and genomic score into four groups: (i) PIK3CA-mutant/Low Genomic Score (Low GS_PIK3CA-Mut), (ii) PIK3CA-mutant/High Genomic Score (High GS_PIK3CA-Mut), (iii) PIK3CA–wild-type/Low Genomic Score (Low GS_PIK3CA-WT), and (iv) PIK3CA–wild-type/High Genomic Score (High GS_PIK3CA-WT). The centre line indicates the median, boxes the interquartile range (IQR), and whiskers 1.5×IQR; individual tumours are shown as jittered points, and the red diamond denotes the group mean. Overall differences were assessed by Kruskal–Wallis test (p-value shown), followed by pairwise Wilcoxon rank-sum tests with significance annotated as *p < 0.05 (*), <0.01 (**), <0.001 (***), and <0.0001 (****)*. A dashed horizontal line marks y = 0. **(C)** Kaplan–Meier plot showing progression-free survival (PFS) of PIK3CA-mutant ER+/HER2− patients in the TCGA cohort stratified by low versus high genomic score. The accompanying table reports pairwise comparisons relative to the indicated reference group, including regression coefficients, hazard ratios (HRs) with 95% confidence intervals (CIs), and p-values. HR > 1 indicates increased risk and HR < 1 indicates reduced risk relative to the reference group. Plot annotations indicate the number of patients per group, the number of events, and the survival probability at the specified time point.

**A**

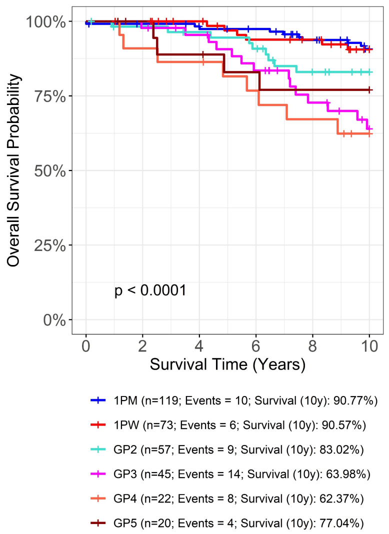


| | Coefficient | HR [CI Lower - CI Upper] | p_value |
|---|---|---|---|
| *1PW vs 1PM* | 0.04 | 1.04 [0.38 - 2.86] | 0.93868 |
| *GP2 vs 1PM* | 0.72 | 2.06 [0.84 - 5.06] | 0.11655 |
| *GP3 vs 1PM* | 1.49 | 4.43 [1.97 - 9.99] | 0.00033 |
| *GP4 vs 1PM* | 1.68 | 5.39 [2.12 - 13.65] | 0.00039 |
| *GP5 vs 1PM* | 1.09 | 2.98 [0.94 - 9.51] | 0.06474 |

**C**

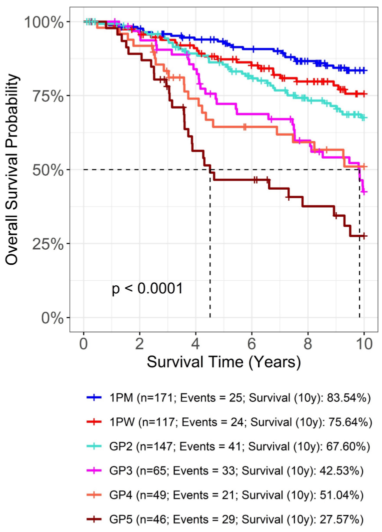


| | Coefficient | HR [CI Lower - CI Upper] | p_value |
|---|---|---|---|
| *1PW vs 1PM* | 0.46 | 1.58 [0.90 - 2.76] | 0.111 |
| *GP2 vs 1PM* | 0.76 | 2.14 [1.30 - 3.52] | 0.003 |
| *GP3 vs 1PM* | 1.43 | 4.19 [2.49 - 7.06] | 7e-08 |
| *GP4 vs 1PM* | 1.38 | 3.96 [2.22 - 7.08] | 3e-06 |
| *GP5 vs 1PM* | 2.00 | 7.36 [4.30 - 12.61] | 3e-13 |

**B**

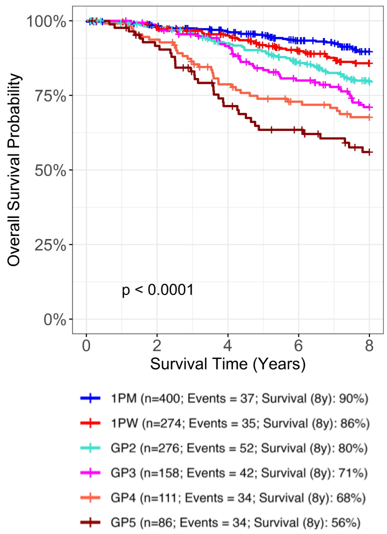


**D**

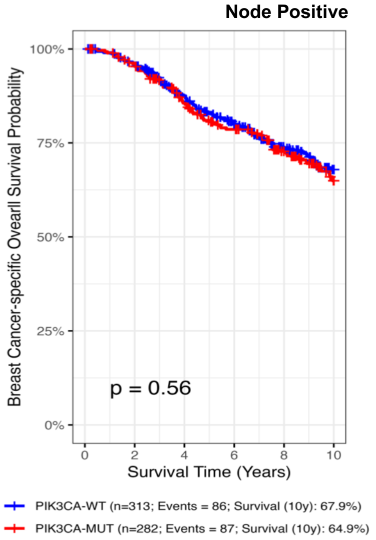


**Supplementary Figure 27 | Tier 2 stratification identifies prognostic subgroups in ER+/HER2– breast cancer. (A-C)** KM survival plot shows breast cancer-specific overall survival of identified Tier 2 subgroups from the METABRIC cohort in **(A)** lymph node-negative ER+/HER2– systemic therapy untreated patients (who have not received chemotherapy and endocrine therapy) at 10 years, **(B)** all ER+/HER2– patients at 8 years, **(C)** lymph-node positive patients at 10 years post-diagnosis from the METABRIC cohort. **(D)** KM survival curve showing the prognostic impact of PIK3CA mutations in lymph-node-positive ER+/HER2– patients, without molecular subtyping (based on population-level averaging of PIK3CA-mutant vs wild-type patients). In all KM plots, breast cancer-specific overall survival probability is shown on the y-axis. For each subgroup, the total number of patients (n), the number of observed events, and the estimated survival probability at the specified time point (accounting for censoring) are reported in the plot legend. The table (below panels A and C) reports pairwise comparisons relative to the reference group as indicated, including regression coefficients, HRs with 95% confidence intervals (CIs), and corresponding *p-values*. The log-rank p-value derived from cox proportional hazards analysis is shown within the plot.

#### Supplementary Figure 28

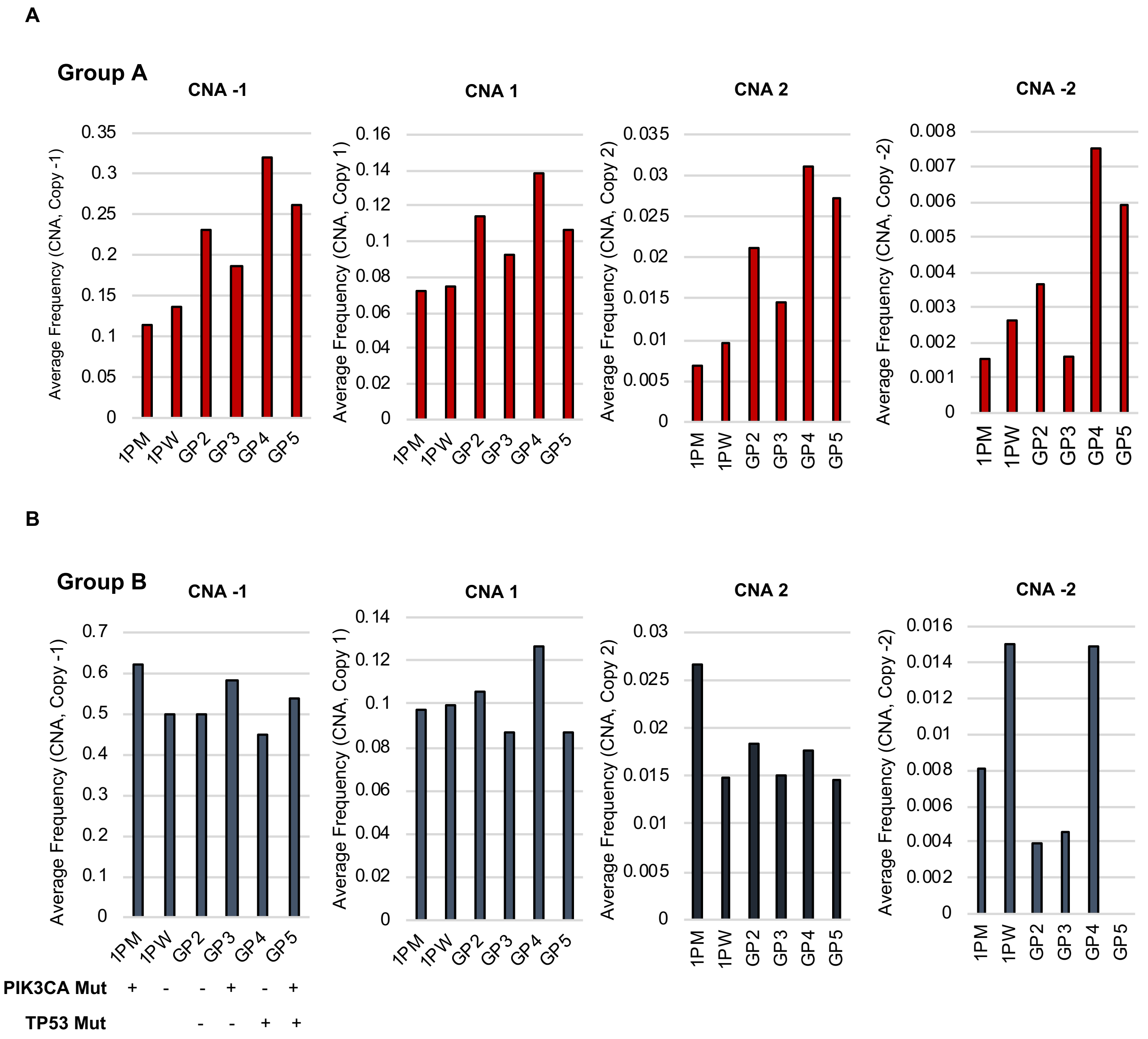


**Supplementary Figure 28 | Copy Number Alteration frequencies across Tier 2 BRCA Subgroups.** Bar plots illustrate the average Copy Number Alteration (CNA) frequencies for **(A)** Group A and **(B)** Group B genes across the Tier 2 subgroups. The low-risk subgroups (1PM, 1PW) exhibit markedly fewer CNAs—particularly copy-number gains (copy 2)—compared with the high-risk subgroups.

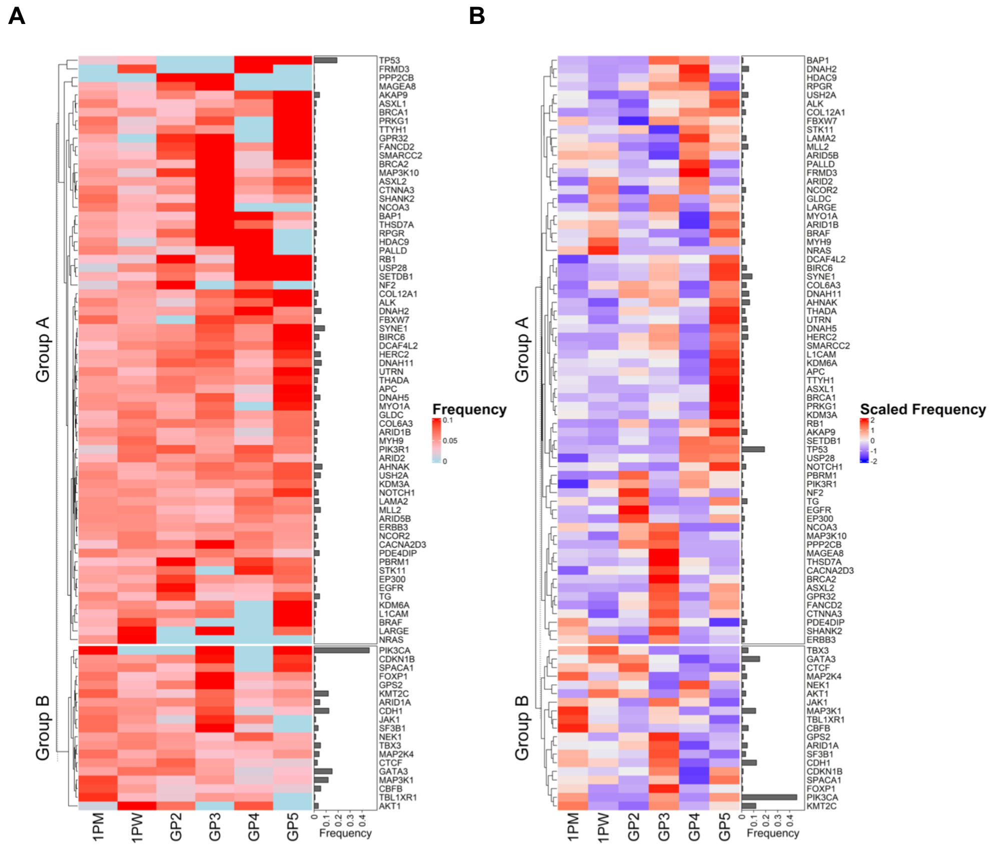


**Supplementary Figure 29 | Group A/Group B genomic mutation frequencies across combined ER+/HER2– METABRIC and TCGA cohorts. (A)** Heatmaps illustrate the frequencies of Group A and Group B genomic mutations in the combined METABRIC and TCGA datasets, after normalising by mutation count for each recurrent gene and by the total sample size for each Tier 2 subgroup. **(B)** The heatmap depicts the row-wise scaled frequencies of Group A and Group B genomic mutations shown in panel A. Adjacent bar charts in both panels display the corresponding mutation frequencies for each gene in ER+/HER2– disease based on combined METABRIC and TCGA datasets.

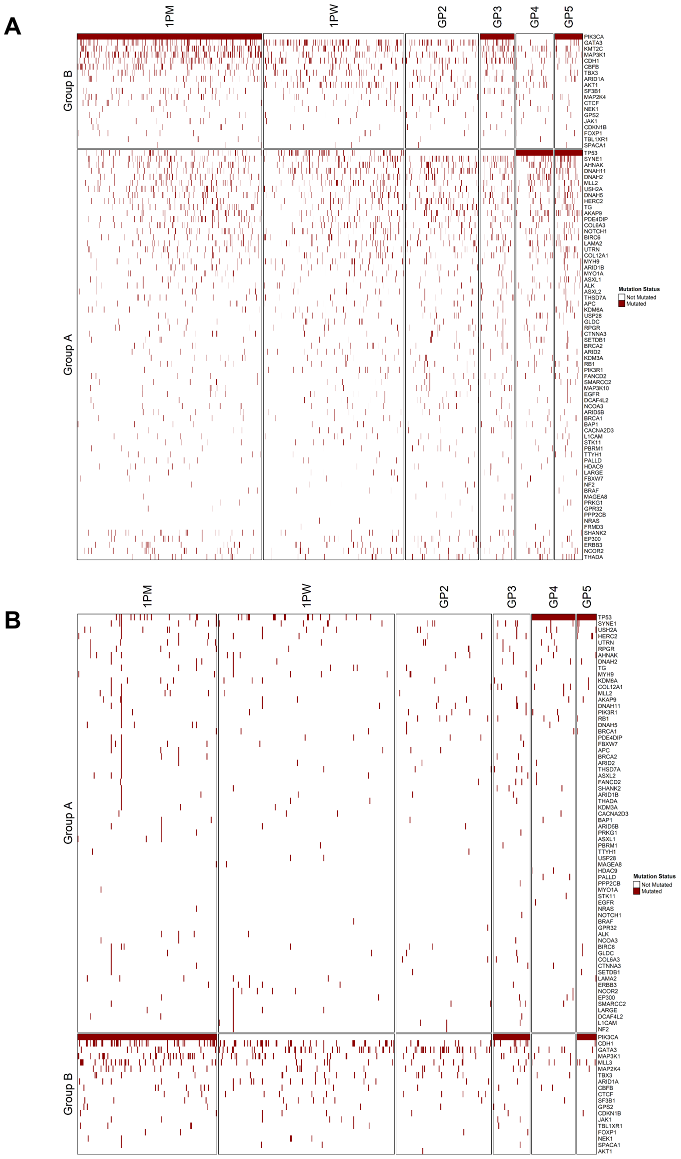


**Supplementary Figure 30 | Tier 2 subgroup-specific mutation frequencies for Group A and Group B genes are visualised as heatmaps for the METABRIC (A) and TCGA (B) cohorts.** In each heatmap, columns represent individual samples, and rows correspond to genes. Group A and Group B genes are separated using row splits, and sample subgroups are annotated through column splits generated using the *ComplexHeatmap* package, with subgroup names shown in the column headers. Overall, the heatmaps illustrate that Group B gene mutations occur more frequently in PIK3CA-mutant subgroups, 1PM and GP3, whereas they are comparatively less pronounced in TP53-mutant subgroups, GP4 and GP5.

### Supplementary Figure 31

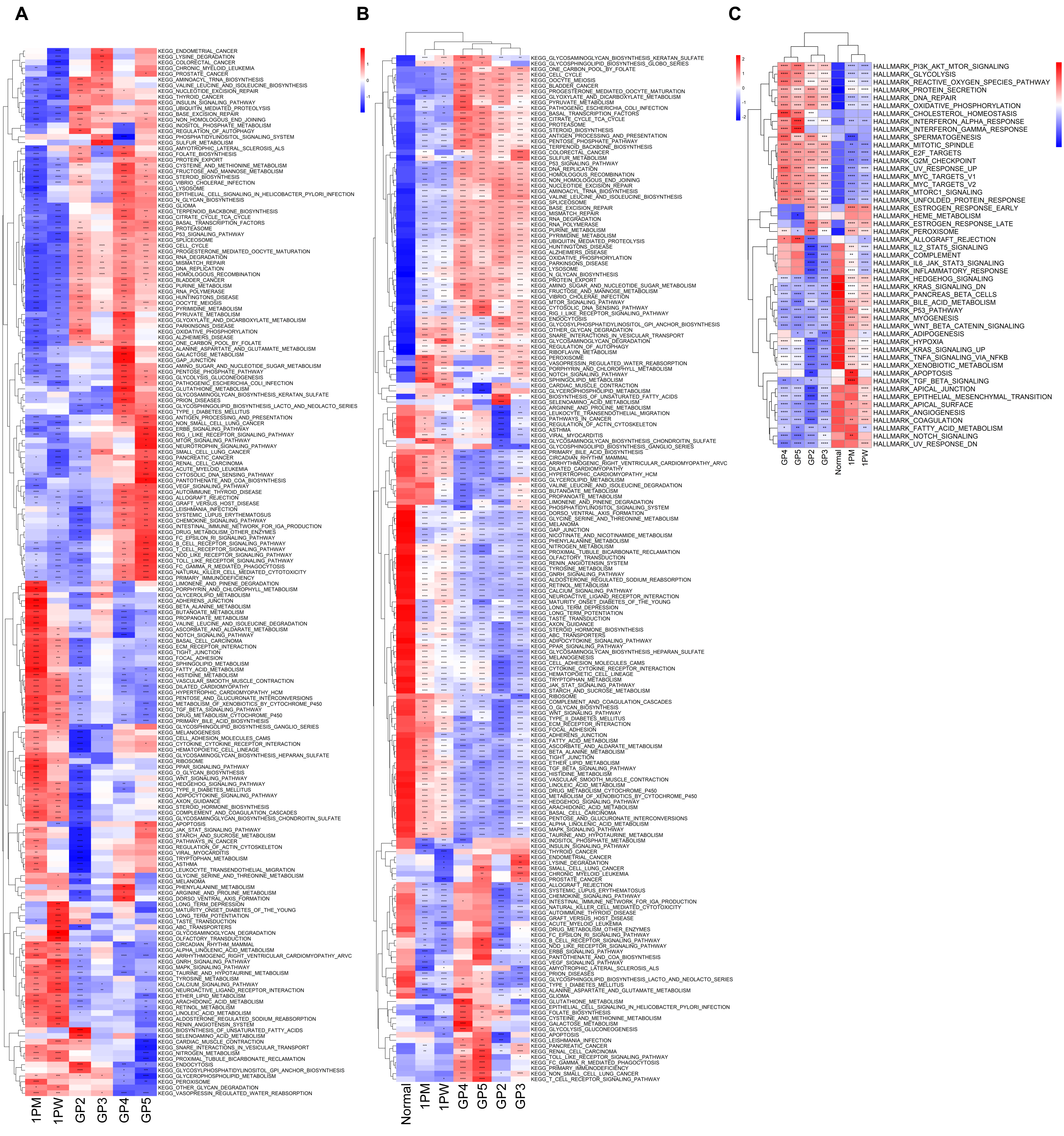


**Supplementary Figure 31 | Distinct GSVA pathway activation patterns across Tier 2 breast cancer subgroups, with and without normal samples.** Heatmaps show the scaled mean GSVA scores of KEGG pathways across Tier 2 subgroups, with normal samples excluded **(A)** and included **(B)**. For heatmaps excluding normal samples, statistical significance was assessed by contrasting each Tier 2 subgroup against all other subgroups using an equal-weighted approach. When normal samples were included, significance was determined by comparing each Tier 2 subgroup with the normal group. Heatmap annotations indicate adjusted p-value thresholds as follows: ***** adj p < 0.0001, *** adj p < 0.001, ** adj p < 0.01, and * adj p < 0.05.* Panel **(C)** presents analogous heatmaps depicting Hallmark pathway activation patterns, including normal samples.

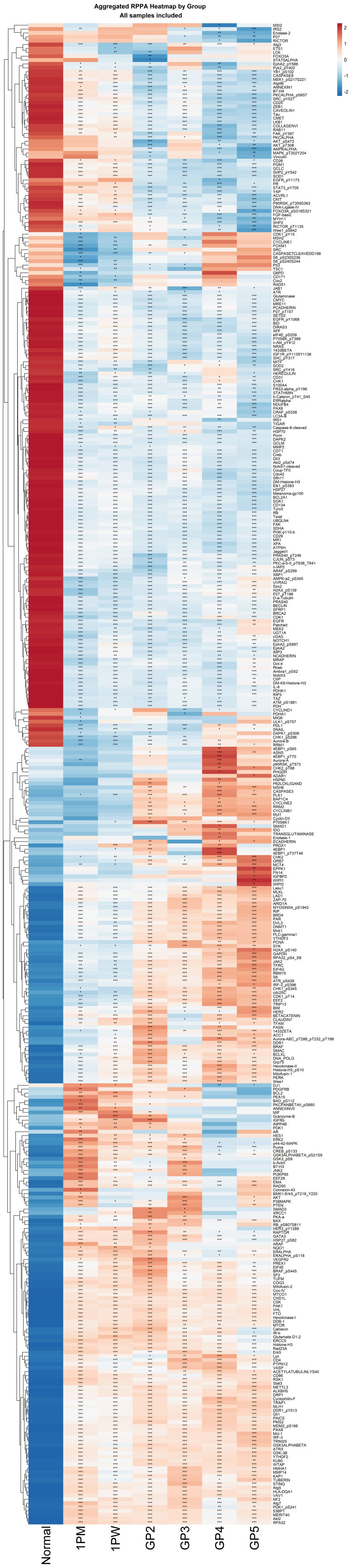


**Supplementary Figure 32 | RPPA profiling reveals that Tier 2 subgroups differ in the levels of total protein abundance and phospho-signalling.** Mean protein/phosphoprotein expression levels across Tier 2 subgroups and normal samples are presented as a heatmap. Statistical significance was assessed by comparing each BRCA subgroup to the normal group. Heatmap annotations denote adjusted p-value thresholds as follows: ***** adj p < 0.0001, *** adj p < 0.001, ** adj p < 0.01, and * adj p < 0.05.*

### Supplementary Figure 33

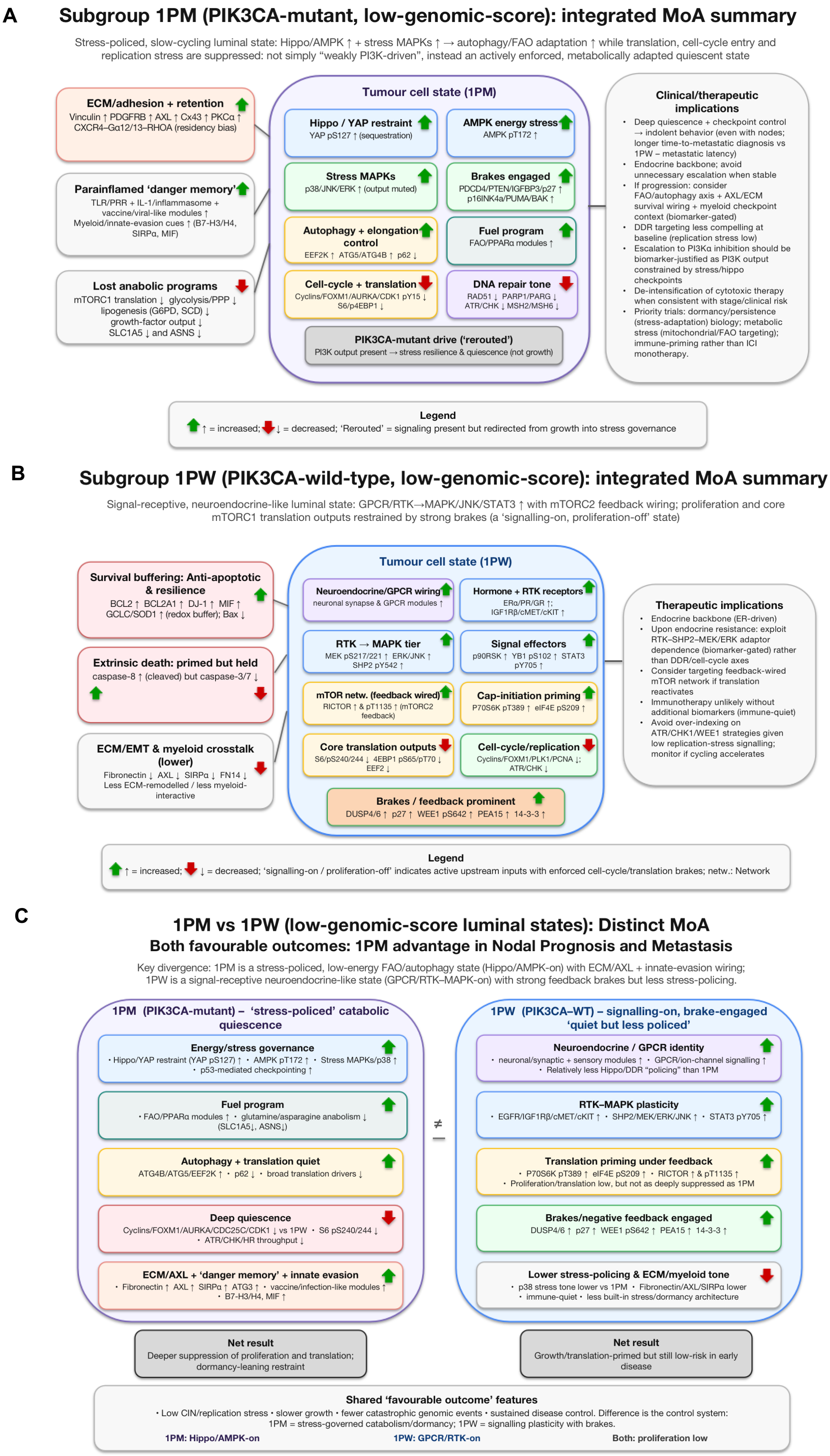


**Supplementary Figure 33 | Integrated mechanism-of-action (MoA) summary of Low Genomic Score subgroups. (A)** 1PM subgroup, **(B)** 1PW subgroup, and **(C)** comparative MoA analysis between 1PM and 1PW.

A

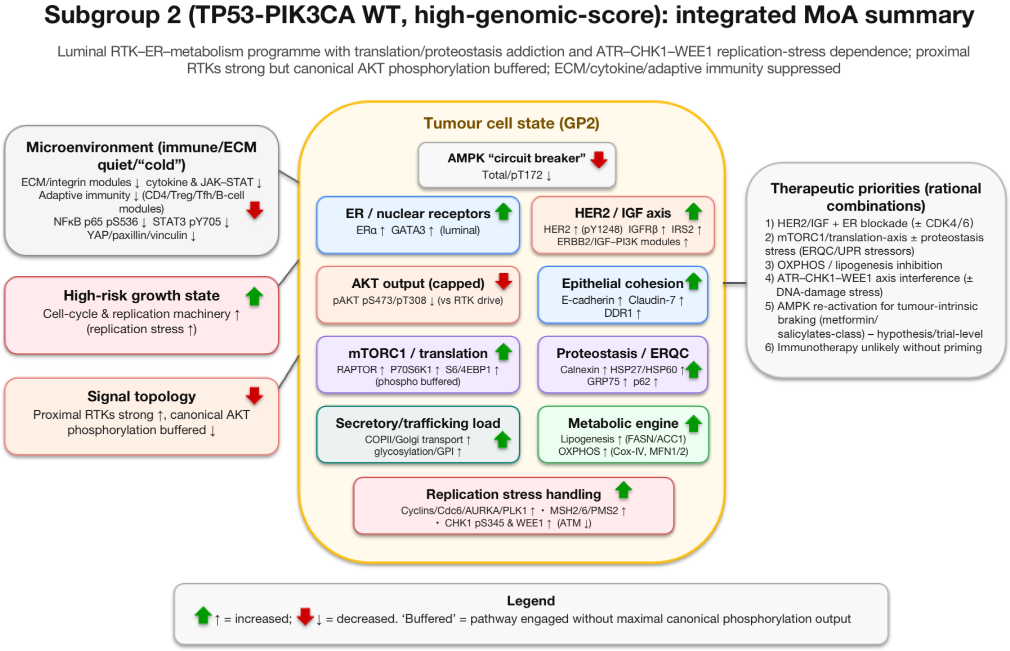
Subgroup 2 (TP53-PIK3CA WT, high-genomic-score): integrated MoA summary
Luminal RTK–ER–metabolism programme with translation/proteostasis addiction and ATR–CHK1–WEE1 replication-stress dependence; proximal RTKs strong but canonical AKT phosphorylation buffered; ECM/cytokine/adaptive immunity suppressed
Microenvironment (immune/ECM quiet/“cold”)
ECM/integrin modules ↓ cytokine & JAK–STAT ↓ Adaptive immunity ↓ (CD4/Treg/Tfh/B-cell modules)
NFκB p65 pS536 ↓ STAT3 pY705 ↓ YAP/paxillin/vinculin ↓
High-risk growth state
Cell-cycle & replication machinery ↑ (replication stress ↑)
Signal topology
Proximal RTKs strong ↑, canonical AKT phosphorylation buffered ↓
Tumour cell state (GP2)
AMPK “circuit breaker”
Total/pT172 ↓
ER / nuclear receptors
ERα ↑ GATA3 ↑ (luminal)
HER2 / IGF axis
HER2 ↑ (pY1248) IGFRβ ↑ IRS2 ↑ ERBB2/IGF–PI3K modules ↑
AKT output (capped)
pAKT pS473/pT308 ↓ (vs RTK drive)
Epithelial cohesion
E-cadherin ↑ Claudin-7 ↑ DDR1 ↑
mTORC1 / translation
RAPTOR ↑ P70S6K1 ↑ S6/4EBP1 ↑ (phospho buffered)
Proteostasis / ERQC
Calnexin ↑ HSP27/HSP60 ↑ GRP75 ↑ p62 ↑
Secretory/trafficking load
COPII/Golgi transport ↑ glycosylation/GPI ↑
Metabolic engine
Lipogenesis ↑ (FASN/ACC1) OXPHOS ↑ (Cox-IV, MFN1/2)
Replication stress handling
Cyclins/Cdc6/AURKA/PLK1 ↑ • MSH2/6/PMS2 ↑ • CHK1 pS345 & WEE1 ↑ (ATM ↓)
Therapeutic priorities (rational combinations)
1) HER2/IGF + ER blockade (± CDK4/6)
2) mTORC1/translation-axis ± proteostasis stress (ERQC/UPR stressors)
3) OXPHOS / lipogenesis inhibition
4) ATR–CHK1–WEE1 axis interference (± DNA-damage stress)
5) AMPK re-activation for tumour-intrinsic braking (metformin/ salicylates-class) – hypothesis/trial-level
6) Immunotherapy unlikely without priming
Legend
↑ = increased; ↓ = decreased. ‘Buffered’ = pathway engaged without maximal canonical phosphorylation output

B

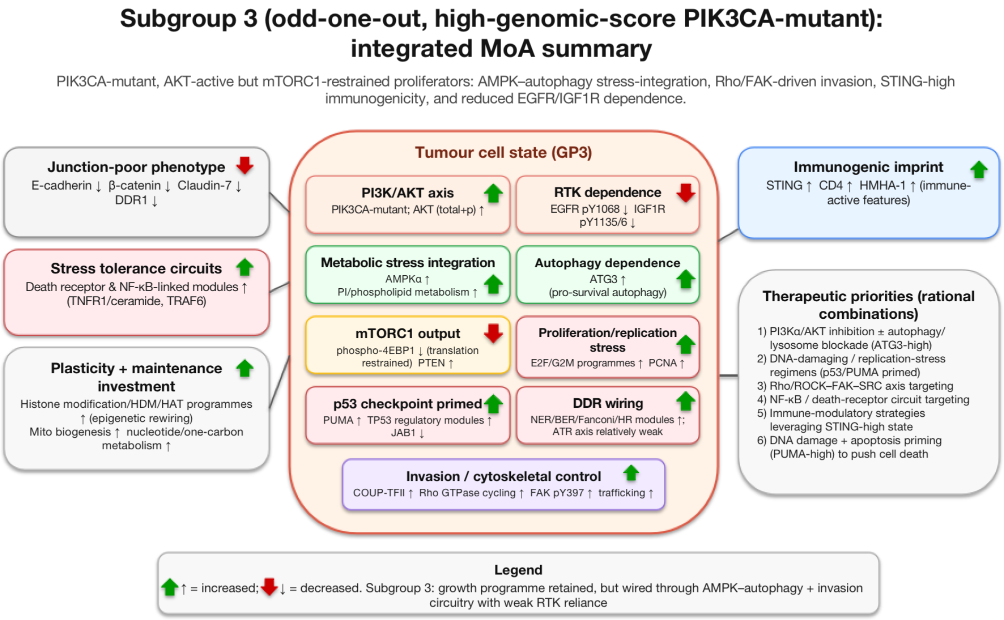
Subgroup 3 (odd-one-out, high-genomic-score PIK3CA-mutant): integrated MoA summary
PIK3CA-mutant, AKT-active but mTORC1-restrained proliferators: AMPK–autophagy stress-integration, Rho/FAK-driven invasion, STING-high immunogenicity, and reduced EGFR/IGF1R dependence.
Junction-poor phenotype
E-cadherin ↓ β-catenin ↓ Claudin-7 ↓ DDR1 ↓
Stress tolerance circuits
Death receptor & NF-κB-linked modules ↑ (TNFR1/ceramide, TRAF6)
Plasticity + maintenance investment
Histone modification/HDM/HAT programmes ↑ (epigenetic rewiring)
Mito biogenesis ↑ nucleotide/one-carbon metabolism ↑
Tumour cell state (GP3)
PI3K/AKT axis
PIK3CA-mutant; AKT (total+p) ↑
RTK dependence
EGFR pY1068 ↓ IGF1R pY1135/6 ↓
Metabolic stress integration
AMPKα ↑ PI/phospholipid metabolism ↑
Autophagy dependence
ATG3 ↑ (pro-survival autophagy)
mTORC1 output
phospho-4EBP1 ↓ (translation restrained) PTEN ↑
Proliferation/replication stress
E2F/G2M programmes ↑ PCNA ↑
p53 checkpoint primed
PUMA ↑ TP53 regulatory modules ↑ JAB1 ↓
DDR wiring
NER/BER/Fanconi/HR modules ↑; ATR axis relatively weak
Invasion / cytoskeletal control
COUP-TFII ↑ Rho GTPase cycling ↑ FAK pY397 ↑ trafficking ↑
Immunogenic imprint
STING ↑ CD4 ↑ HMHA-1 ↑ (immune-active features)
Therapeutic priorities (rational combinations)
1) PI3Kα/AKT inhibition ± autophagy/ lysosome blockade (ATG3-high)
2) DNA-damaging / replication-stress regimens (p53/PUMA primed)
3) Rho/ROCK–FAK–SRC axis targeting
4) NF-κB / death-receptor circuit targeting
5) Immune-modulatory strategies leveraging STING-high state
6) DNA damage + apoptosis priming (PUMA-high) to push cell death
Legend
↑ = increased; ↓ = decreased. Subgroup 3: growth programme retained, but wired through AMPK–autophagy + invasion circuitry with weak RTK reliance

**C**

#### Subgroup 4 (TP53-mutant/PIK3CA-WT, high-genomic-score group): integrated MoA summary

Translation-addicted + replication-stressed state with metabolic rewiring and antioxidant buffering. ERK input remains intact but is not the dominant quantitative driver; differentiation/homeostatic circuits suppressed; immune-evasion via PD-L1/IDO.

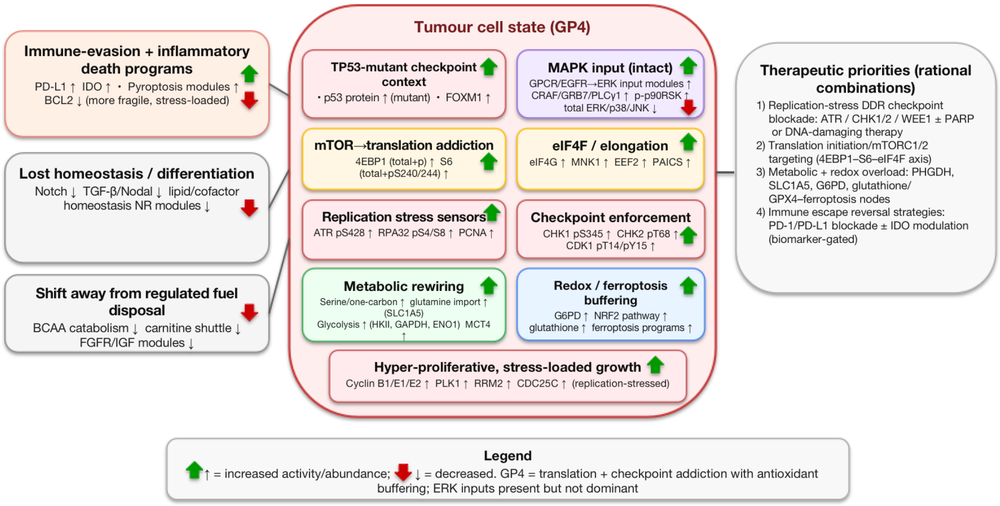


**D**

#### Subgroup 5 (TP53-PIK3CA double mutant, high-genomic-risk score): integrated MoA summary

Checkpoint-primed proliferators with chronic Innate immune/Type-I IFN–wiring, PI3K-linked immune-receptor signals, and preserved translation/DDR, but redox detoxification compromised (oxidative-overload vulnerability) and relatively low ERK

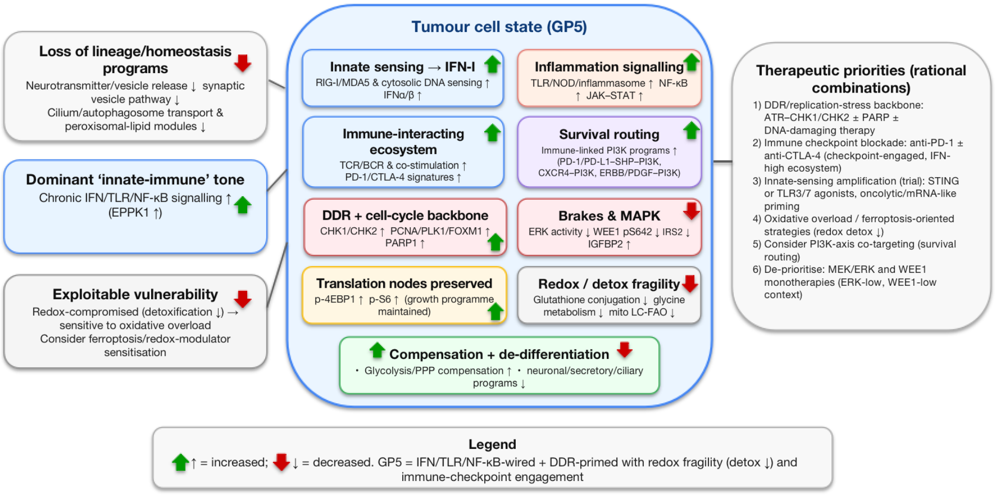


**Supplementary Figure 34 | Integrated mechanism-of-action (MoA) summary of High Genomic Score subgroups.** **(A)** GP2, **(B)** GP3, **(C)** GP4 and **(D)** GP5.

### Supplementary Figure 35

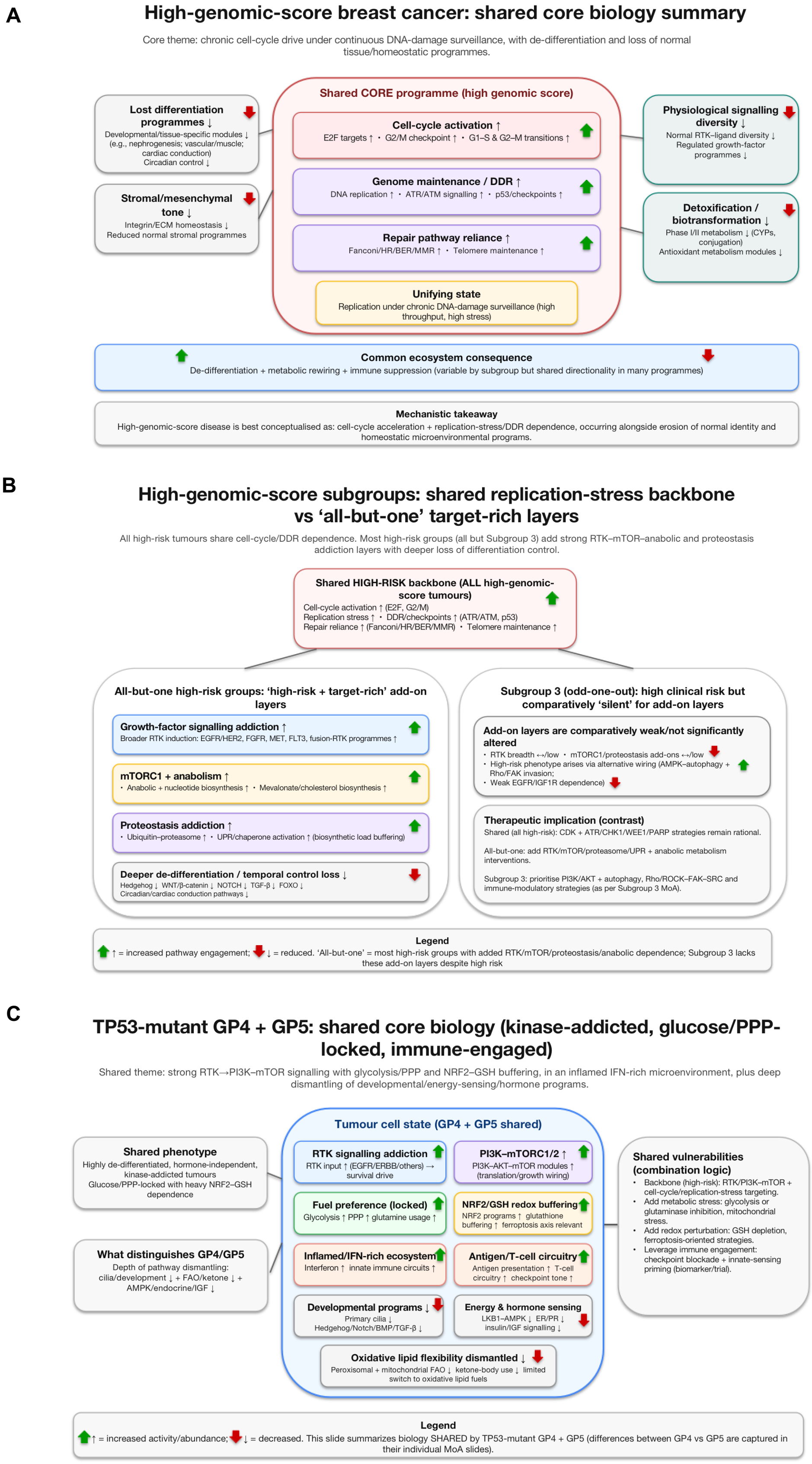


**Supplementary Figure 35 | Shared dominant programmes between High Genomic Score subgroups. (A)** Shared core biology between all high Genomic Score tumours. **(B)** Shared core biology across all but one high genomic score subgroups, **(C)** Shared core biology between GP4 and GP5 subgroups.

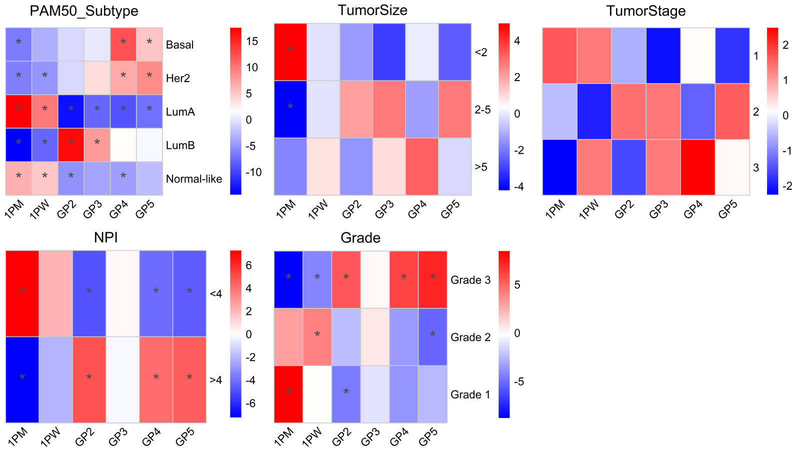


**Supplementary Figure 36 | Residual heatmaps showing clinicopathological enrichments across Tier 2 subgroups.** Associations between Tier 2 subgroups and clinicopathological variables (PAM50 subtype, tumour size, tumour stage, Nottingham Prognostic Index, and histological grade) were assessed using chi-square contingency tests. To identify which subgroup–category combinations most strongly drove each association, standardised Pearson residuals were computed for each cell of the contingency tables and visualised as a heatmap. Positive residuals (red) indicate categories observed more frequently than expected under independence (over-representation), whereas negative residuals (blue) indicate categories observed less frequently than expected (under-representation). Colour intensity reflects the absolute magnitude of the residual. Asterisks denote cells with absolute standardised residual > 3.

### Supplementary Figure 37

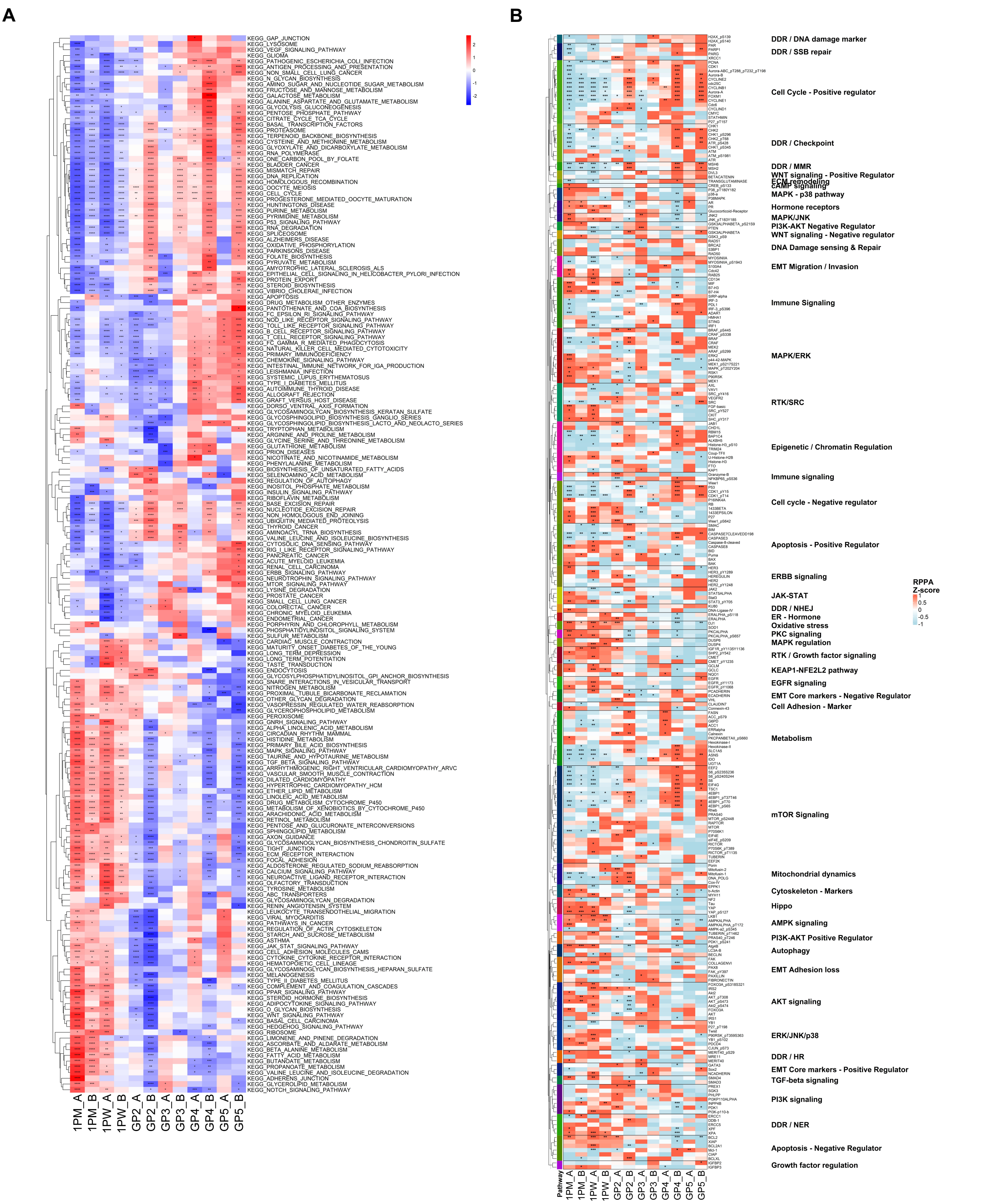


**Supplementary Figure 37 | Tier 3 stratification reveals the gradient nature of tumour biology as seen through pathway and RPPA profiling. (A)** Heatmap showing the scaled mean GSVA scores of KEGG pathways across Tier 3 breast cancer subgroups. **(B)** Mean RPPA protein/phosphoprotein levels from TCGA-RPPA data across Tier 3 subgroups are shown in a heatmap. Statistical significance was assessed by contrasting each BRCA subgroup against all other subgroups using an equal-weighted method. Significance is annotated as: *adj p < 0.0001 (****), adj p < 0.001 (***), adj p < 0.01 (**), adj p < 0.05 (*)*. Suffix A denotes a below-median score; B denotes an above-median score. Proteins were annotated using curated pathway mappings, grouped by pathway, and classified by their expected functional effect in breast cancer (positive vs negative regulators); these annotations are displayed alongside the heatmap.

### Supplementary Figure 38

A

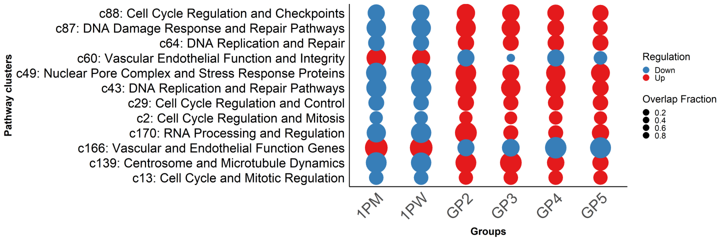


B

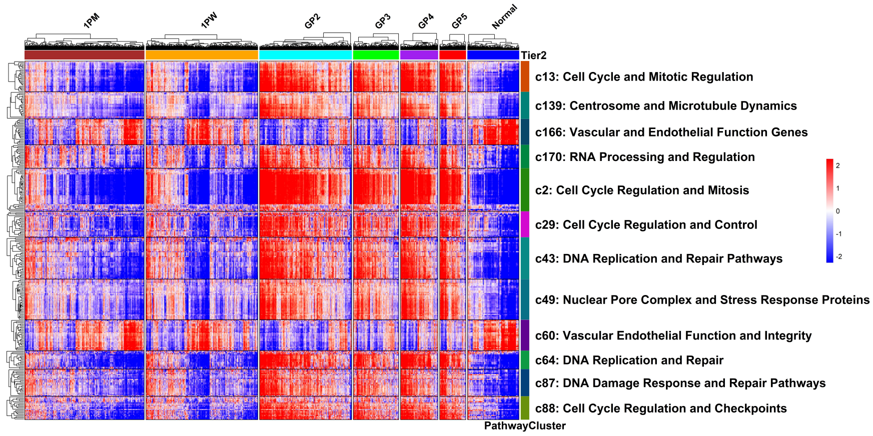


C

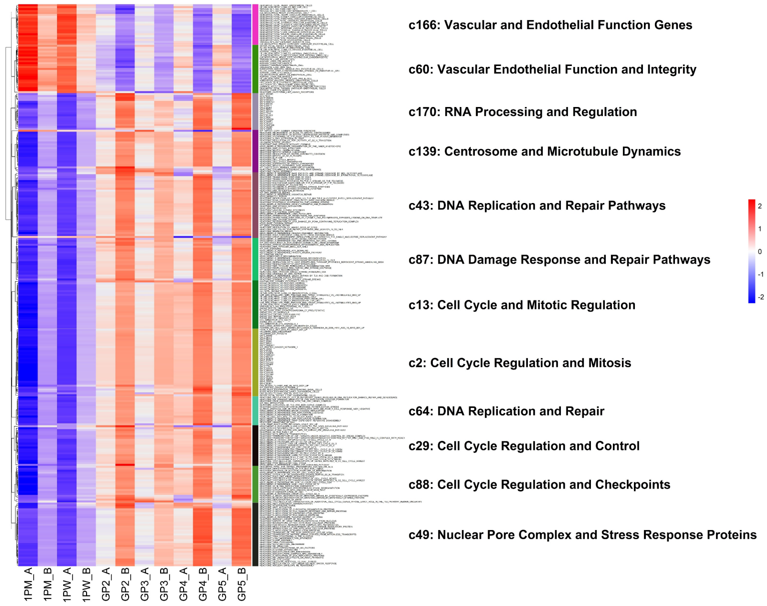


**Supplementary Figure 38 | Core proliferative and replicative programmes across Tier 3 strata. (A)** Dot plots summarise the pathway clusters that are significantly enriched (*adjusted p < 0.05*) in each Tier 2 subgroup compared with all other subgroups. Each column represents a subgroup contrast and each row an enriched pathway cluster; dot colour indicates directionality (red: enrichment/activation in the subgroup; blue: depletion/repression relative to other subgroups). Vascular/endothelial clusters are preferentially enriched in lower genomic-risk strata, whereas cell-cycle and DNA replication clusters are preferentially enriched in higher genomic-risk strata. **(B)** Sample-level heatmaps show pathway activity scores across individual tumours, with columns grouped by Tier 2 subgroup. Rows correspond to pathway gene sets annotated by their associated cluster terms, and top annotations indicate subgroup membership. These heatmaps recapitulate the subgroup-specific activation and repression patterns observed in (A). **(C)** Heatmap related to *Figure 9B* showing mean pathway activity scores for the indicated cluster pathways across Tier 3 subgroups. This highlight attenuated activation of cell-cycle–associated programmes in a subset of tumours representing a biologically “quieter” state within nominally high-risk Tier 2 classes (GP2A, GP3A, GP4A, GP5A). Conversely, within nominally low-genomic-risk Tier 2 groups, a subset showed higher-than-expected activity (1PMB, 1PWB).

### Supplementary Figure 39

A

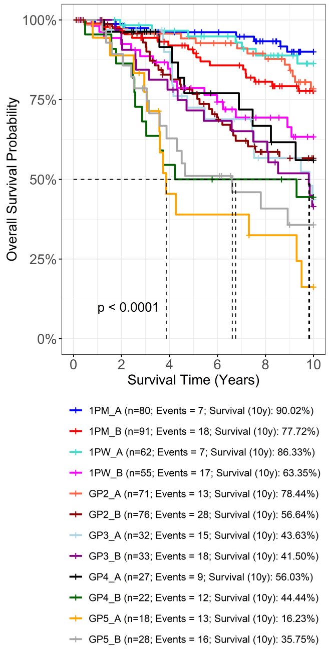


B

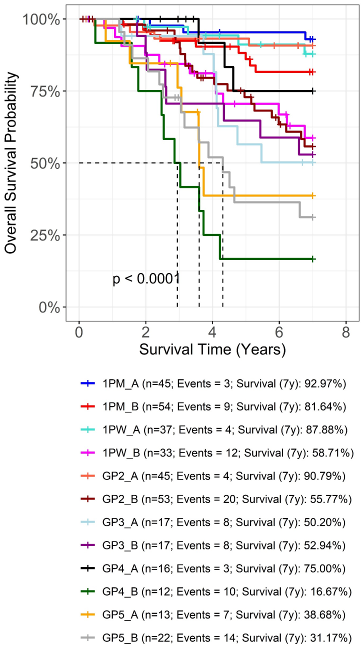


C

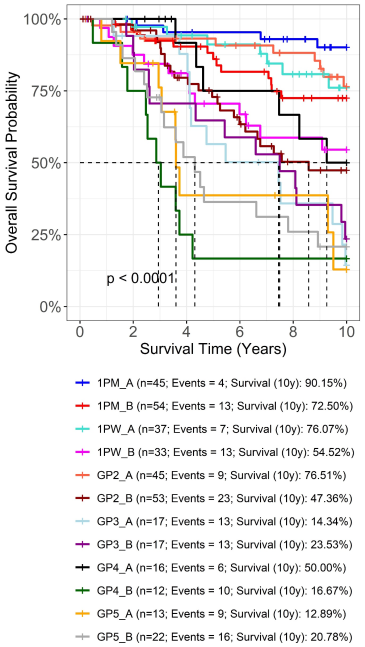


| | Coefficient | HR [CI Lower - CI Upper] | p_value |
|---|---|---|---|
| 1PM_B vs 1PM_A | 0.90 | 2.47 [1.03 - 5.92] | 0.04 |
| 1PW_A vs 1PM_A | 0.31 | 1.37 [0.48 - 3.90] | 0.55 |
| 1PW_B vs 1PM_A | 1.56 | 4.76 [1.97 - 11.49] | 0.0005 |
| GP2_A vs 1PM_A | 0.76 | 2.14 [0.86 - 5.37] | 0.10 |
| GP2_B vs 1PM_A | 1.75 | 5.75 [2.51 - 13.17] | <0.0001 |
| GP3_A vs 1PM_A | 1.93 | 6.89 [2.81 - 16.91] | <0.0001 |
| GP3_B vs 1PM_A | 2.05 | 7.80 [3.26 - 18.67] | <0.0001 |
| GP4_A vs 1PM_A | 1.63 | 5.12 [1.91 - 13.76] | 0.001 |
| GP4_B vs 1PM_A | 2.25 | 9.49 [3.74 - 24.12] | <0.0001 |
| GP5_A vs 1PM_A | 2.79 | 16.22 [6.46 - 40.77] | <0.0001 |
| GP5_B vs 1PM_A | 2.41 | 11.16 [4.58 - 27.17] | <0.0001 |

| | Coefficient | HR [CI Lower - CI Upper] | *p_value* |
|---|---|---|---|
| *1PM_B vs 1PM_A* | 1.03 | 2.81 [0.76 - 10.39] | 0.121 |
| *1PW_A vs 1PM_A* | 0.56 | 1.75 [0.39 - 7.81] | 0.47 |
| *1PW_B vs 1PM_A* | 1.98 | 7.25 [2.04 - 25.69] | 0.002 |
| *GP2_A vs 1PM_A* | 0.33 | 1.40 [0.31 - 6.24] | 0.662 |
| *GP2_B vs 1PM_A* | 1.99 | 7.34 [2.18 - 24.72] | 0.001 |
| *GP3_A vs 1PM_A* | 2.2 | 8.99 [2.38 - 33.93] | 0.001 |
| *GP3_B vs 1PM_A* | 2.2 | 8.98 [2.38 - 33.88] | 0.001 |
| *GP4_A vs 1PM_A* | 1.24 | 3.46 [0.70 - 17.14] | 0.13 |
| *GP4_B vs 1PM_A* | 3.38 | 29.27 [8.01 - 106.96] | <0.0001 |
| *GP5_A vs 1PM_A* | 2.67 | 14.37 [3.71 - 55.74] | <0.0001 |
| *GP5_B vs 1PM_A* | 2.8 | 16.39 [4.70 - 57.18] | <0.0001 |

| | Coefficient | HR [CI Lower - CI Upper] | *p_value* |
|---|---|---|---|
| *1PM_B vs 1PM_A* | 1.17 | 3.22 [1.05 - 9.87] | 0.04 |
| *1PW_A vs 1PM_A* | 0.91 | 2.49 [0.73 - 8.52] | 0.14 |
| *1PW_B vs 1PM_A* | 1.88 | 6.57 [2.14 - 20.17] | 0.001 |
| *GP2_A vs 1PM_A* | 0.87 | 2.40 [0.74 - 7.78] | 0.14 |
| *GP2_B vs 1PM_A* | 2.01 | 7.43 [2.57 - 21.52] | 0.0002 |
| *GP3_A vs 1PM_A* | 2.57 | 13.08 [4.26 - 40.19] | <0.0001 |
| *GP3_B vs 1PM_A* | 2.5 | 12.19 [3.97 - 37.42] | <0.0001 |
| *GP4_A vs 1PM_A* | 1.73 | 5.62 [1.59 - 19.93] | 0.007 |
| *GP4_B vs 1PM_A* | 3.25 | 25.92 [8.09 - 83.00] | <0.0001 |
| *GP5_A vs 1PM_A* | 2.81 | 16.65 [5.11 - 54.22] | <0.0001 |

**Supplementary Figure 39 | Tier 3 stratification improves risk discrimination in lymph-node–positive disease. (A-C)** Kaplan–Meier analysis shows breast cancer-specific overall survival of **(A)** lymph-node–positive ER+/HER2− patients (≥1 positive node) in the METABRIC cohort (n=595) at 10-years post-diagnosis. The y-axis shows the survival probability; patients with follow-up beyond 10 years were censored. **(B)** METABRIC lymph-node–positive ER+/HER2− patients with ≥2 positive nodes, overall survival censored at 10 years. **(C)** METABRIC lymph-node–positive ER+/HER2− patients with ≥2 positive nodes, overall survival censored at 7 years. The accompanying tables below each survival plot report pairwise comparisons versus the indicated reference group (regression coefficient, HR with 95% CI, and p-value). Plot annotations indicate the number of patients per group, the number of events, and the survival estimate. The log-rank p-value from the Cox proportional hazards analysis is shown in the plot.

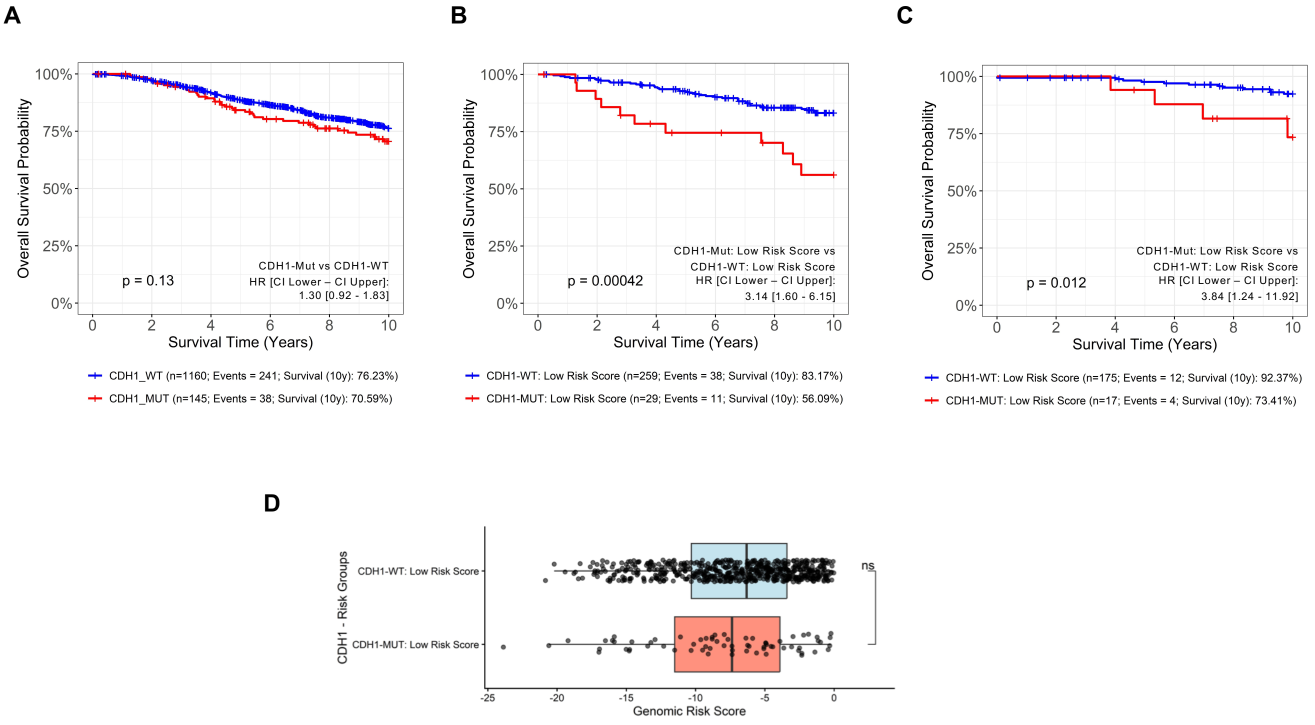


**Supplementary Figure 40 | Subgroup-specific prognostic associations of CDH1 mutations in ER+/HER2− breast cancer in METABRIC. (A-C)** Kaplan–Meier (KM) curves show breast cancer–specific overall survival for: **(A)** CDH1-mutant vs wild-type patients in the full cohort without molecular subtyping (population-level comparison); **(B)** CDH1-mutant tumours within lymph node–positive, low–genomic score ER+/HER2− patients; **(C)** CDH1-mutant tumours within lymph node-negative systemic therapy untreated patients (patients who have not received chemotherapy and endocrine therapy). Survival probability at 10 years is plotted on the y-axis; patients with follow-up beyond the displayed time horizon were censored. For each group, the number of patients (n), number of events, and estimated survival probability at the specified time point (accounting for censoring) are shown in the plot legend. Tables accompanying each panel report comparisons versus the indicated reference group, including hazard ratios (HRs) with 95% confidence intervals (CIs). The log-rank p-value from the Cox proportional hazards model is displayed on each plot. **(D)** Distribution of composite genomic scores in CDH1-mutant and CDH1–wild-type low-risk subgroups, shown as boxplots. Statistical comparison using the Wilcoxon rank-sum test revealed no significant difference (ns).

### Supplementary Figure 41

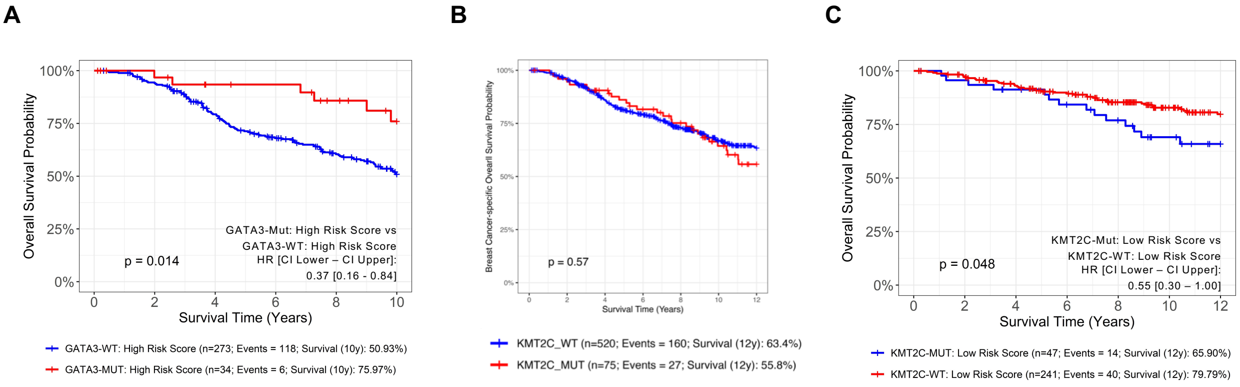


**Supplementary Figure 41 | Subgroup-specific prognostic associations of Tier 4 modifier mutations (GATA3 and KMT2C) in ER+/HER2− breast cancer in METABRIC. (A–C)** Kaplan–Meier (KM) curves show breast cancer–specific overall survival for: **(A)** GATA3-mutant tumours within lymph node–positive, high–genomic score ER+/HER2− patients (10-year horizon); **(B)** KMT2C-mutant vs wild-type patients in the full cohort without molecular subtyping (population-level comparison); and **(C)** KMT2C-mutant tumours within lymph node–positive, low–genomic score ER+/HER2− patients (12-year horizon). Survival probability is plotted on the y-axis; patients with follow-up beyond the displayed time horizon were censored. For each group, the number of patients (n), number of events, and estimated survival probability at the specified time point (accounting for censoring) are shown in the plot legend. Tables accompanying each panel report comparisons versus the indicated reference group, including hazard ratios (HRs) with 95% confidence intervals (CIs). The log-rank p-value from the Cox proportional hazards model is displayed on each plot.

### Supplementary Figure 42

A

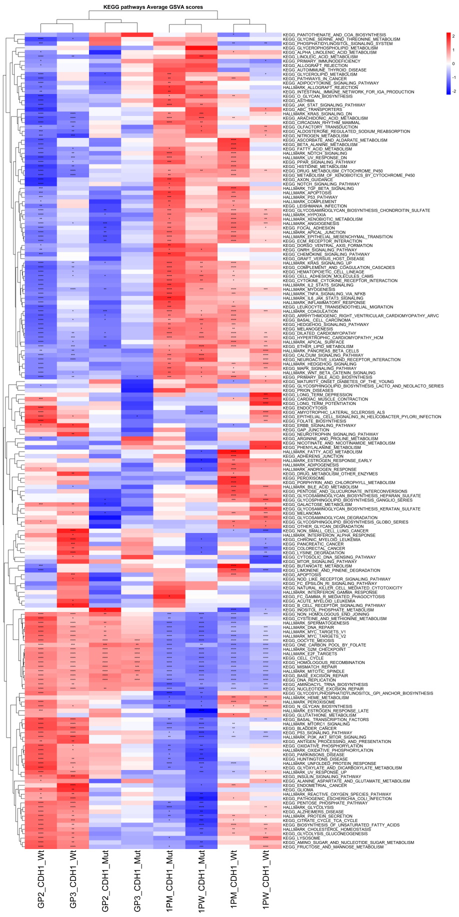


B

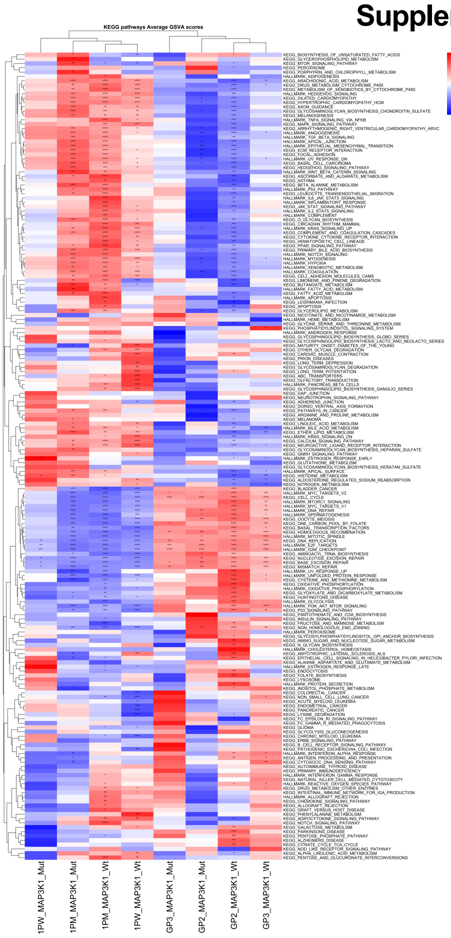


C

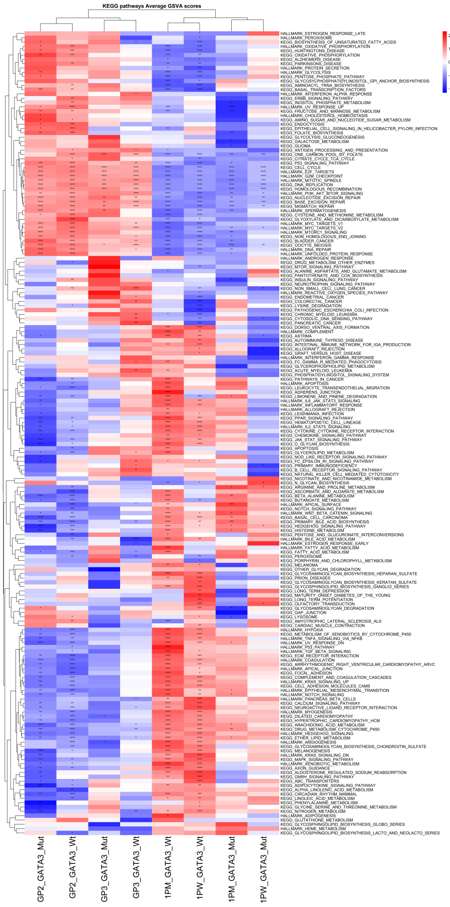


**Supplementary Figure 42 | Tier 4 modifier mutation overlays refine tumour biology.** Heatmaps of scaled mean GSVA Hallmark pathway scores across Tier 2 subgroups, further stratified by Tier 4 mutation overlays: CDH1 **(A)**, MAP3K1 **(B)**, and GATA3 **(C)**. Columns represent Tier 4 mutation status within each Tier 2 subgroup; rows represent Hallmark pathways. Statistical significance is annotated as: *adj p < 0.0001 (****), adj p < 0.001 (***), adj p < 0.01 (**), adj p < 0.05 (*)*. Column-wise hierarchical clustering shows that Tier 4 mutation overlays remain transcriptionally aligned with their parent Tier 2 subgroup, consistent with modifier effects that refine phenotype without redefining core identity.

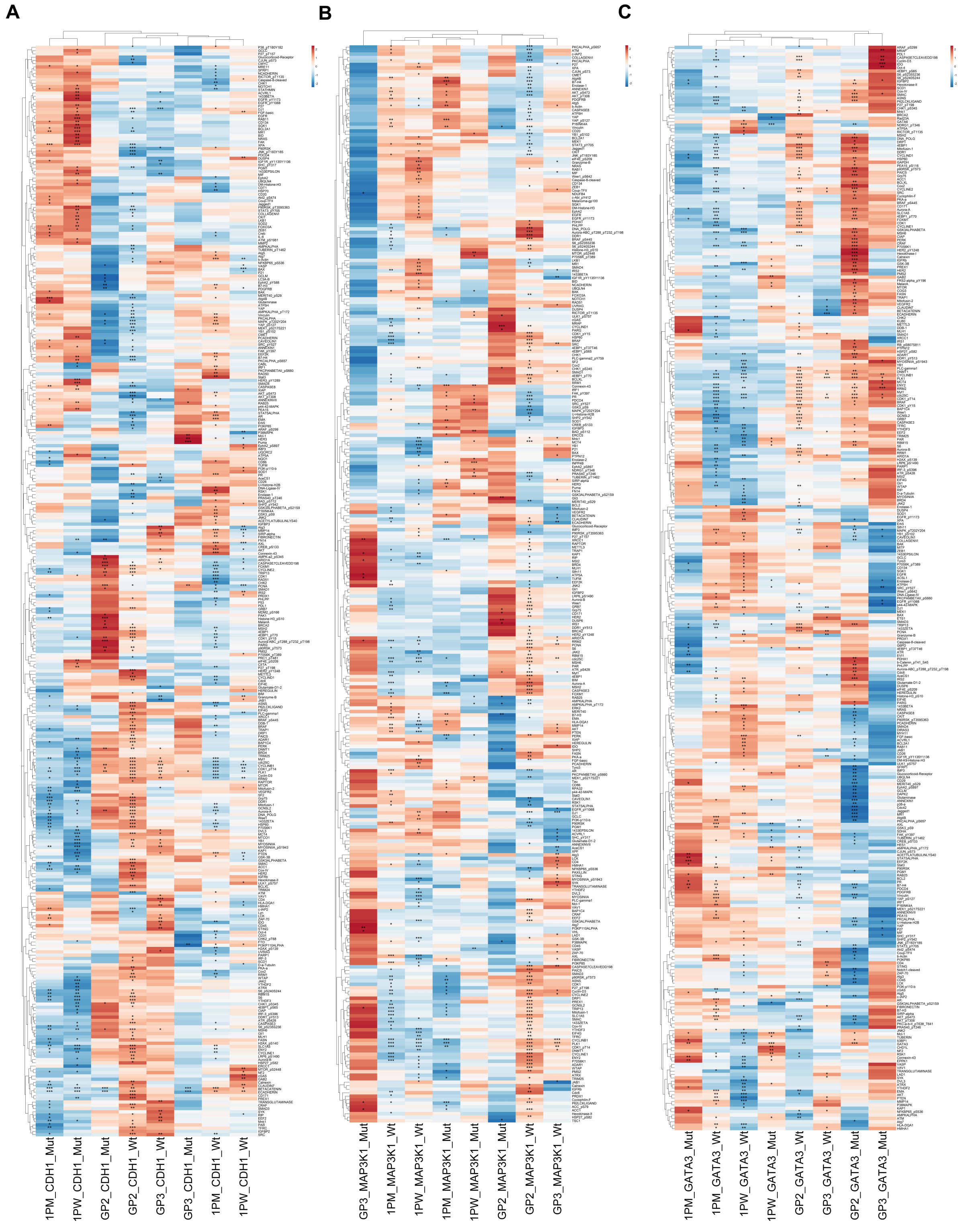


**Supplementary Figure 43 | Tier 4 modifier mutation overlays and RPPA profiling.** Heatmaps show scaled mean RPPA protein/phosphoprotein expression levels across Tier 2 breast cancer subgroups stratified by key mutations, including CDH1 **(A)**, MAP3K1 **(B)**, and GATA3 **(C)**. Statistical significance was assessed by comparing each Tier 4 subgroup with all other subgroups using an equal-weighted approach, and results are annotated as follows: **** *p-value < 0.0001*, *** *p-value < 0.001*, ** *p-value < 0.01*, and * *p-value < 0.05*.

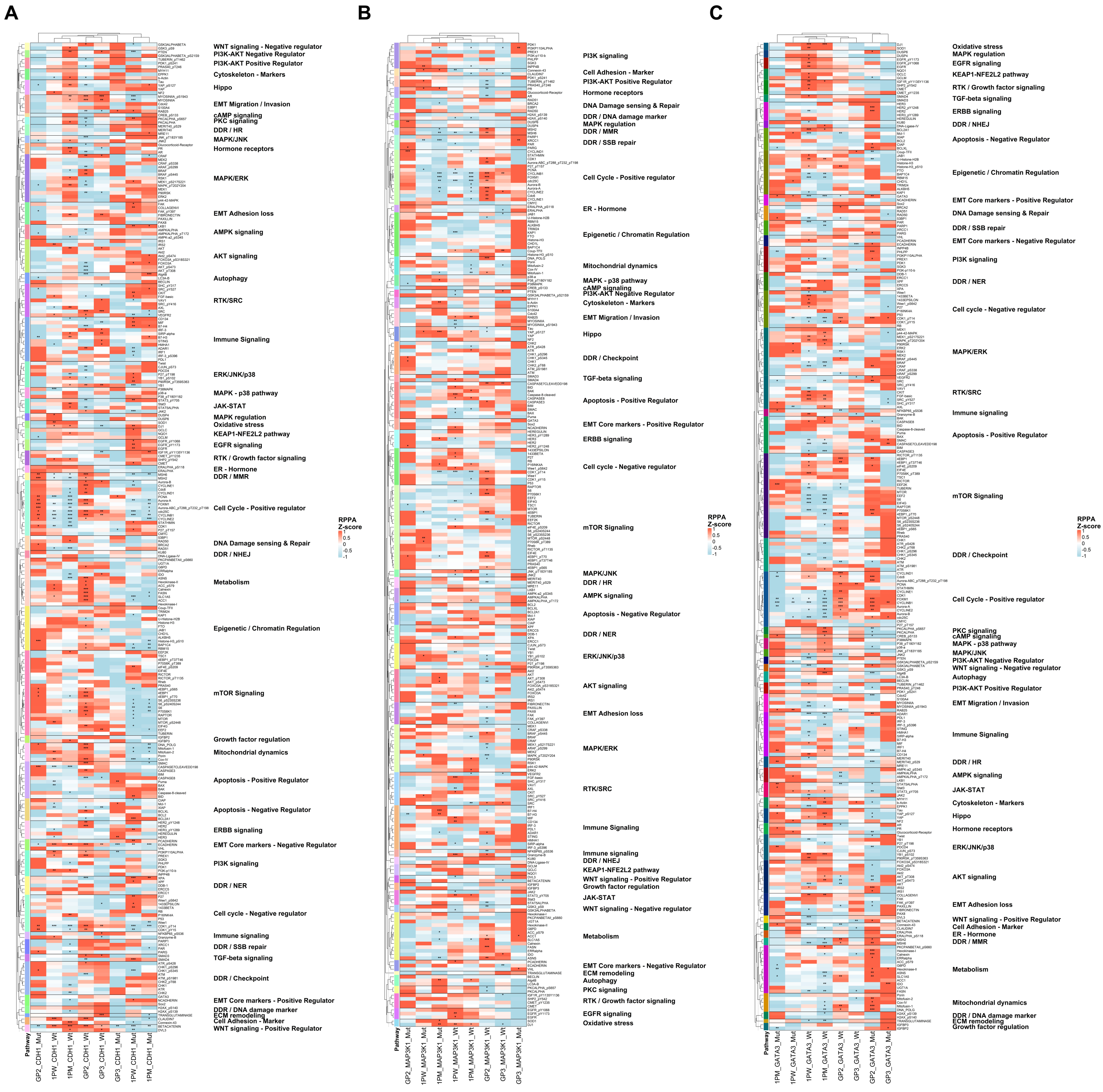


**Supplementary Figure 44 | Tier 4 modifier mutation overlays and annotated RPPA heatmaps.** Heatmaps show scaled mean RPPA protein/phosphoprotein levels across Tier 2 breast cancer subgroups, stratified by Tier 4 modifier mutation status for CDH1 **(A)**, MAP3K1 **(B)**, and GATA3 **(C)**. For each Tier 4–defined subgroup, statistical significance was assessed using an equal-weighted comparison against all other subgroups, with significance annotated as **** $p < 0.0001$, *** $p < 0.001$, ** $p < 0.01$, and * $p < 0.05$. Proteins were annotated using curated pathway mappings, grouped by pathway, and classified by expected functional effect in breast cancer (positive vs negative regulators); these annotations are shown alongside the heatmaps.

### Supplementary Figure 45

**A**

#### CDH1-mutant tumour subgroup: integrated MoA summary

Junction-disrupted, plastic epithelial state with inflammatory lipid conditioning, RTK–STAT3 survival wiring, FOXO/SGK stress adaptation, and suppressed mTORC1–glycolysis–anabolism, associated with poorer outcomes

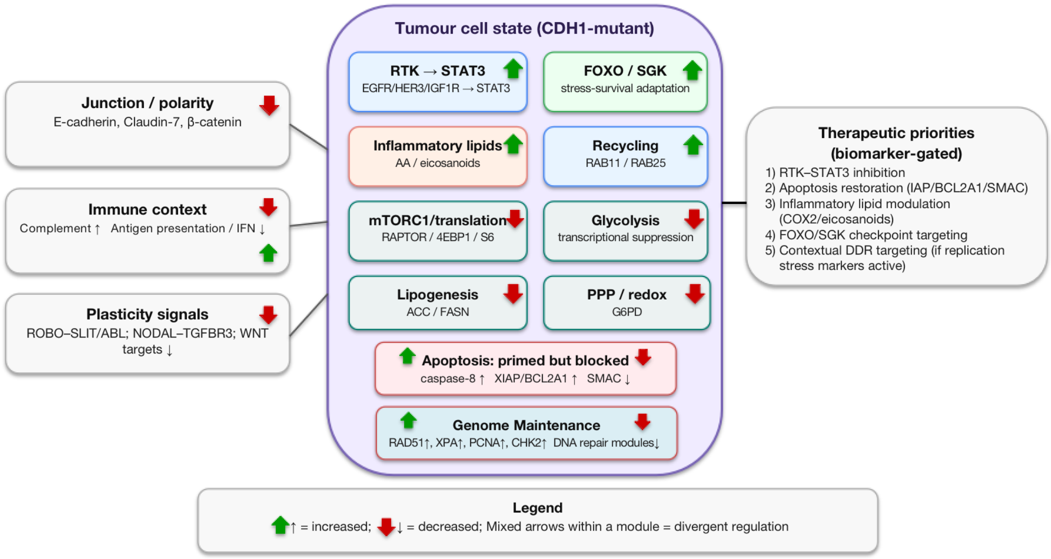


**B**

#### MAP3K1-mutant tumour subgroup: integrated MoA summary

Oxidative, metabolically fit luminal state associated with favourable outcome: FAO/BCAA/urea + glutathione/redox ↑; tonic ERK under feedback ↑ with RTK/RAS inputs ↓; mitotic/replication-stress machinery ↓; immune-quiet chemokine/GPCR tone ↓

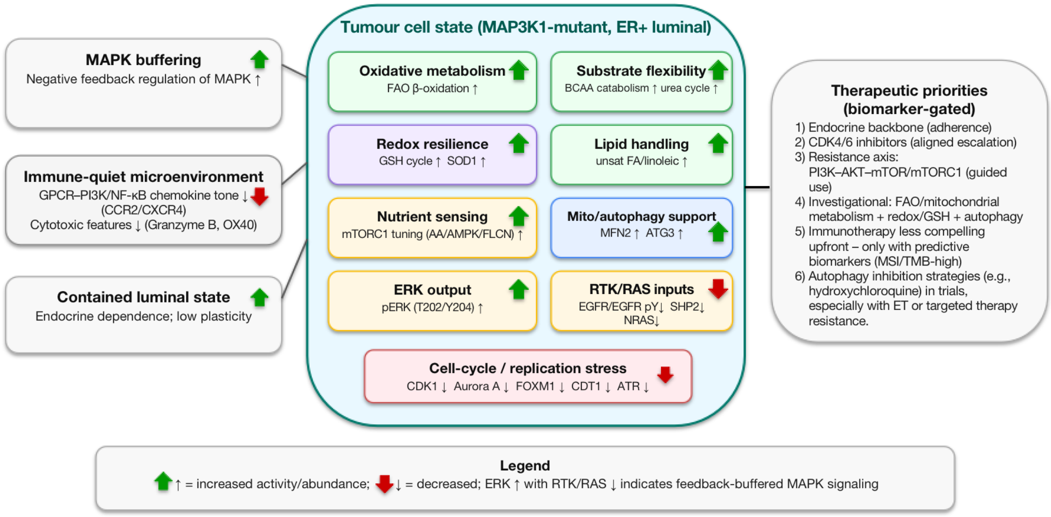

**C**

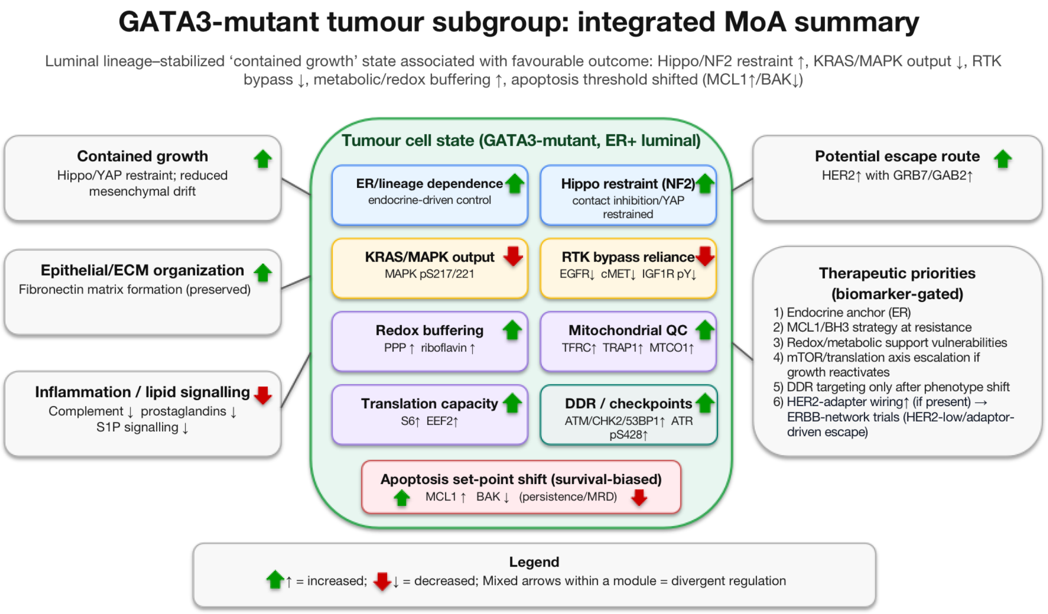


**Supplementary Figure 45 | Integrated mechanism-of-action (MoA) summary of Tier 4 modifier mutations. (A)** CDH1-mutant tumor subgroup, **(B)** MAP3K1-mutant tumor subgroup and **(C)** GATA3-mutant tumor subgroup.

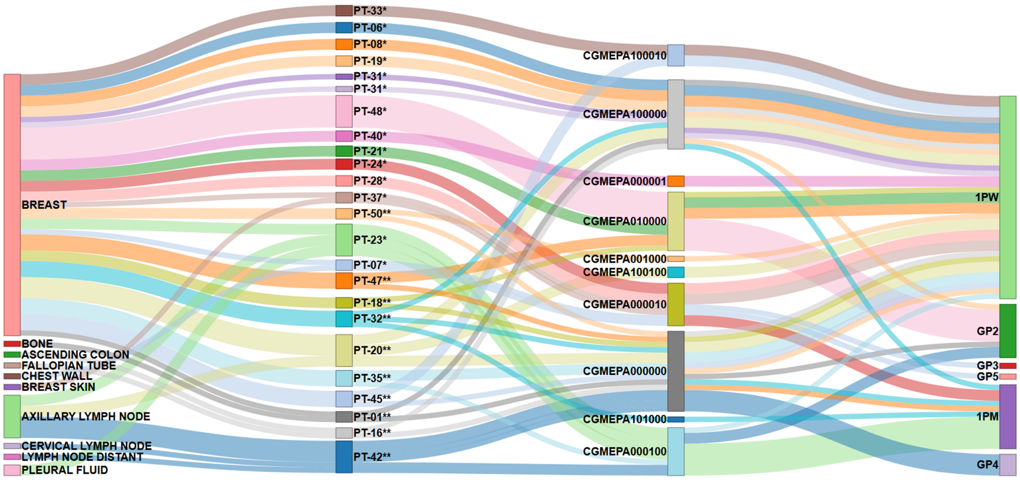


**Supplementary Figure 46 | Patient-level alluvial plot reveals two non-overlapping Tier 4 mutation patterns across multi-site sampling in metastatic breast cancer.** Alluvial plots track breast cancer specimens from tissue site through patient identity and T-OMICS classifications, including Tier 4 mutation status and Tier 2 subgroup assignment. Each colour denotes an individual patient. Two distinct Tier 4 patterns are observed: (i) Stable, truncal Tier 4 alterations (marked with an asterisk *)—26% of patients harboured a Tier 4 mutation in at least one of the queried genes that was conserved across all additional specimens from the same patient, with no new Tier 4 events detected upon expanded sampling; and (ii) Variable/acquired Tier 4 alterations (marked with a double asterisk **)—20% of patients showed lesion-to-lesion differences, where at least one specimen contained additional Tier 4 events absent in another specimen, consistent with inter-lesional divergence and/or therapy-associated adaptation in a subset.

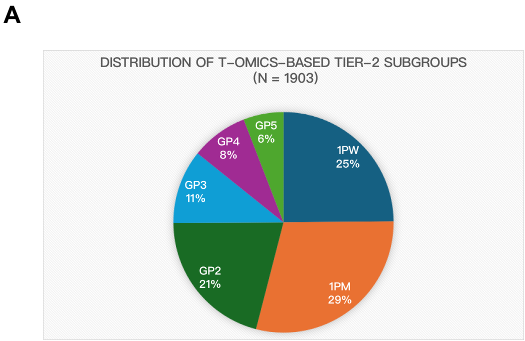


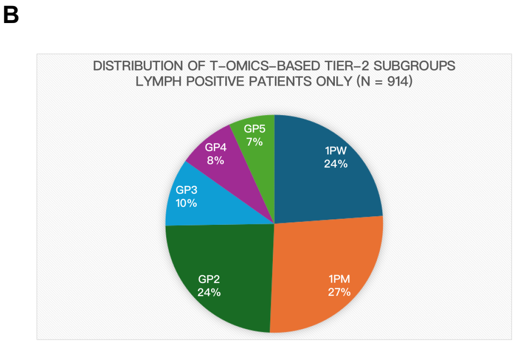


**Supplementary Figure 47 | Tier 2 subgroup composition of the analysed cohorts.** Pie charts show the proportion of samples assigned to each predefined subgroup in the full analysed cohorts **(A)** and in the subset restricted to lymph node–positive cases **(B)**. Percentages indicate the fraction of samples in each subgroup within the corresponding cohort/subset.

A

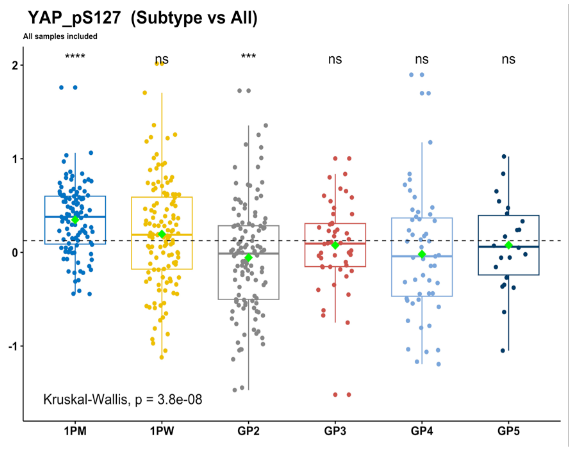


B

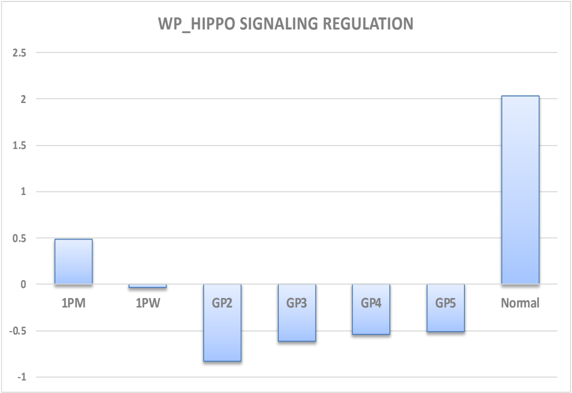


**Supplementary Figure 48 | Active Hippo signalling in 1PM.** (**A)** Boxplot shows RPPA-based phosphoprotein levels of YAP1 pS127. **(B)** Barchart shows levels of WP_Hippo Signalling Regulation module in Tier 2 subgroups. Proteomically, 1PM shows elevated phosphorylation of YAP at S127 (suggesting cytoplasmic sequestration via Hippo-mediated inhibition), corroborated by transcriptomic enrichment of multiple Hippo/YAP-restraining modules. The centre line indicates the median. Overall differences were assessed by Kruskal–Wallis test (p-value shown), followed by pairwise Wilcoxon rank-sum tests with significance annotated as *p < 0.05 (*), <0.01 (**), <0.001 (***), and <0.0001 (****)*. A dashed horizontal line in (A) indicates the mean YAP1 phosphorylation level across all samples.
